# Novel dPCR-Based Approach for SARS-CoV-2 Variant Detection and Monitoring in Wastewater: A Multi-State Comparison with Clinical Genotyping and GISAID Sequencing Data

**DOI:** 10.1101/2024.08.07.24311627

**Authors:** Patrick Acer, Patrick Andersen, Robbie Barbero, Stephanie Barksdale, Sophia Bellakbira, Dalton Bunde, Ross Dunlap, James Erickson, Daniel Goldfarb, Tara Jones-Roe, Michael Kilroy, Hien Le, Ben Lepene, Emily Milich, Ayan Mohamed, Sayed Mosavi, Denton Munns, Jared Obermeyer, Anurag Patnaik, Ganit Pricer, Marion Reven, Dalaun Richardson, Chamodya Ruhunusiri, Saswata Sahoo, Lauren P. Saunders, Olivia Swahn, Kalpita Vengurlekar, David White, Jean Lozach, Aouda Patricia Flores-Baffi, Fletcher Easton, Maya Dahlke, Andrea Fang, David Cibin, Tim Wesselman, Orlando Sablon, Marlene Wolfe, Pengbo Liu, Stephen Hilton, Yuke Wang, Christine L. Moe, Siya Kashwala, Erica Camarato, Shreya Shrestha, Regan Wied, Adelaide Roguet, Kayley Janssen, Dagmara Antkiewicz, Ian Bradley, Sydney Gallo, Yinyin Ye, Sarah Kane, Jim Huang, Johannah Gillespie, Andrew Jones, Mayumi E. Pascual, Dolores Sanchez Gonzalez, Michael Secreto, Rachel Poretsky, Sarah Owens, Stephanie M. Greenwald, Rose Wilton, Elizabeth Donahue

**Affiliations:** Ceres Nanosciences, Inc.; Rosalind, Inc; Emory University; University of Wisconsin Madison; University at Buffalo-SUNY; GT Molecular; University of Illinois Chicago; Argonne National Laboratory

**Keywords:** Wastewater, COVID-19, genotyping, mutations, Omicron, Polymerase Chain Reaction (PCR), SARS-CoV-2, Variants, Next-Generation Sequencing (NGS), Wastewater-based Epidemiology (WBE), Digital PCR (dPCR)

## Abstract

Wastewater testing has emerged as an effective tool for monitoring levels of SARS-CoV-2 infection in sewered communities. As of July 2024, PCR-based methods continue to be the most widely used methods in wastewater surveillance (1–3). Data from PCR-based wastewater testing is usually available to public health authorities in near real time, typically within 5 to 7 days after waste enters the sewer (4,5). Unfortunately, while these methods can accurately detect and quantify SARS-CoV-2, they are not usually used to differentiate between the multitude of variants, including variants that are classified as Variants of High Consequence (VOHC) and Variants of Concern (VOC) (6). Currently, to identify these variants, the extracted nucleic acids must be analyzed using resource-intensive sequencing-based methods. Moreover, not every lab has access to sequencing technology, so availability of equipment and expertise is also a roadblock besides These costly and time-consuming sequencing methods, while informative, diminish some of the early warning benefits provided by wastewater surveillance. Moreover, not every lab has access to sequencing technology, creating additional barriers due to the availability of equipment and expertise.

In response to these analytical shortcomings, we developed and assessed an alternative approach for variant monitoring in wastewater using customizable dPCR-based genotyping assays. This approach is an expansion from a previously described method for analyzing clinical samples utilizing customizable qPCR-based genotyping. Relative to sequencing, this approach is cost-effective, fast, and easily implemented.

We combined the dPCR-based wastewater genotyping approach along with the well-established Nanotrap^Ⓡ^ Particles virus concentration method as part of a wastewater processing protocol to perform SARS-CoV-2 genotyping in five wastewater testing labs across multiple regions in the United States. The results for the wastewater genotyping approach are displayed on a public-facing dashboard alongside clinical genotyping results and GISAID data (see https://tracker.rosalind.bio).

Despite conducting genotyping on fewer wastewater samples than clinical samples, our approach effectively detected signals of emerging variants and trends in SARS-CoV-2 variants within the community, similar to clinical analyses. For instance, in Georgia, the rapid rise and dominance of the Unknown and BA.2.86*/JN* variants in early 2024 were consistently observed in wastewater samples and closely matched trends in the GISAID clinical sequencing database. Similarly, the EG.5* and FL* variants showed elevated signals in wastewater before clinical detection, highlighting the early warning potential of wastewater testing. Detailed analysis of multiple datasets from various states revealed consistency in the rise and fall of variants across wastewater genotyping, clinical genotyping, and GISAID data. This consistency demonstrates that the prevalence of variants in wastewater closely matches that in clinical settings, underscoring the capability of wastewater-based surveillance to provide extended monitoring of circulating variants, often preceding clinical detections by several weeks.

We further assessed the wastewater genotyping approach by calculating positive percent agreement for detection of four variants (JN, EG.5, FL, and XBB) between the genotyping results and whole genome sequencing results for a set of 129 matched samples that were analyzed using both methods. The agreement ranged between 54% agreement for FL to 97% agreement for JN, with an average of 76% agreement across all samples for all four variants.

Additionally, we estimate that collecting and analyzing data using the dPCR genotyping method is significantly less expensive and time-consuming compared to next-generation sequencing. Labs that outsource next-generation sequencing face much higher costs and longer delays. Transitioning to multiplex dPCR for variant detection could further reduce both cost and turnaround time.

Finally, we discuss the challenges and lessons learned in the development, validation, and implementation of dPCR-based wastewater genotyping. These findings support the use of wastewater-based surveillance as a complementary approach to clinical surveillance, offering a broader and more inclusive picture of variant prevalence and transmission in the community.

## Introduction

Public health surveillance of infectious diseases plays a critical role in public health safety by informing prevention and control measures and the mitigation of outbreaks (7,8). One innovative approach that has garnered significant attention is wastewater surveillance, a method that involves monitoring pathogens in sewage to provide an early warning system for the presence and spread of diseases within communities (9–12). Studies show that microbial and viral pathogens, as well as other biomarkers, can be detected in human excreta and subsequently enter the wastewater system (13–15). Since both symptomatic and asymptomatic infected individuals, along with those lacking access to healthcare, contribute to the wastewater system, wastewater-based epidemiology (WBE) has become a valuable tool for unbiased community health monitoring and early outbreak detection (12,16,17). This technique has been particularly valuable in the context of the SARS-CoV-2 pandemic, enabling public health officials to track the virus’s spread and infection rates in a non-invasive manner (18,19). By analyzing and quantifying the genetic material of the virus shed in untreated wastewater, researchers can gain insights into spatial and temporal patterns of the virus infection, even in asymptomatic populations, thus offering a comprehensive view of the public health landscape (18,20,21).

As SARS-CoV-2 continues to mutate, giving rise to variants with differing levels of transmissibility and virulence, the monitoring of the appearance and abundance of these variants has become a pivotal aspect of both clinical and wastewater sample surveillance (22,23). Variant monitoring typically involves the genetic sequencing and analysis of virus samples to identify mutations and categorize them into specific variants (24). This process is crucial for tracking the emergence and distribution of new variants, assessing their potential impact on public health, and guiding vaccination and public health response strategies. Wastewater testing can detect the presence of emerging SARS-CoV-2 variants before they are identified in clinical settings, providing a crucial head start in the race to contain potential outbreaks in a community (9). By offering a timely and accurate picture of circulating variants, this approach informs clinical and public health strategies, enabling the optimization of diagnostic tests, the adjustment of treatment protocols, and the deployment of vaccines that are effective against current variants.

Next-generation sequencing (NGS) is the primary method for monitoring SARS-CoV-2 genetic lineages in the U.S., analyzing around 5% of positive clinical samples (25). NGS laboratory methods are complex, time-consuming, and costly. While NGS testing methods can have lower sensitivity compared to PCR methods in certain instances, many testing labs do not possess their own sequencing instruments or expertise required to perform NGS (25–27).

Wastewater viral sequencing is not always a practical solution for variant monitoring in testing labs. However, SNP genotyping assays can be developed for rapid screening of wastewater samples by PCR (25,28,29). Once novel mutation sites are identified, specific primers for these sites can be developed for routine screening by sensitive techniques such as dPCR and qPCR.

In this study, we developed and assessed a novel method to provide a fast and cost-effective solution for SARS-CoV-2 variant detection in wastewater. We combined the widely utilized Nanotrap Particle wastewater processing method with a PCR-based genotyping approach that previously had been demonstrated and deployed for clinical sample genotyping (30). We assessed the viability of utilizing qPCR-based variant genotyping panels for identifying variant percentages in wastewater samples and then tested their applicability with a dPCR system commonly used in U.S. wastewater testing laboratories. Results uploaded to the [ROSALIND tracker](https://tracker.rosalind.bio/tracker/dashboard/?start-date=2023-04-01&end-date=2024-05-31&selected-date=2024-08-26&graph-display=percentage&metric=cases&graph-controls=cases&graph-controls=hospitalizations&graph-controls=deaths&graph-controls=positivity&graph-daily-weekly=daily&selected-lab=all-labs) for real-time SARS-CoV-2 genotyping. The ROSALIND tracker calculates variant percentages and presents the results in a public dashboard alongside clinical genotyping and GISAID sequencing data.

We then describe how we used this approach to test wastewater samples over a 13-month period using dPCR panels of two to four mutations to identify SARS-CoV-2 variants in six different states. We compare the results of the wastewater genotyping to the clinical genotyping and clinical sequencing results for multiple geographic areas where the wastewater testing was conducted. Some of the wastewater samples that were genotyped were also processed for next-generation sequencing in parallel, and the results of those sequencing analyses are compared to the genotyping. Our results show that wastewater-based genotyping closely resembles the clinical variant profile in each geographic area. While each clinical specimen typically has a single source and represents only one SARS-CoV-2 variant (apart from rare events such as co-infection or mutagenesis within the host), wastewater-based genotyping can detect early signals and indicate extended periods of variant circulation in the community without any clinical implications.

At a national level, the wastewater genotyping method clearly shows the rise of the JN variant, with the first detection happening in November 2023 and rapidly becoming the dominant variant in the wastewater by the end of January 2024. These results closely mirror the results for JN in the GISAID data (31,32).

## Methods

The wastewater processing and digital PCR (dPCR) genotyping workflow involves four key steps: sample collection, automated viral concentration and nucleic acid extraction, digital PCR analysis, and data visualization (Figure 1). Wastewater samples are initially collected from influent lines at treatment facilities. These samples are processed using Nanotrap Enhancement Reagent 1 (ER1) and Nanotrap Microbiome A Particles coupled with MagMAX™ Wastewater Ultra Nucleic Acid Isolation Kit, which are added to an automated machine (KingFisher Apex) to concentrate, purify, and extract RNA. The RNA samples undergo digital RT-PCR analysis to detect and quantify SARS-CoV-2 RNA. The resulting data is visualized to show the composition of different SARS-CoV-2 variants over time and their location, with a color-coded bar chart representing daily variant composition and a prevalence summary detailing the percent abundance of various identified lineages and markers. This workflow facilitates the monitoring and analysis of SARS-CoV-2 variants in the community through wastewater surveillance.

**Figure 1:**
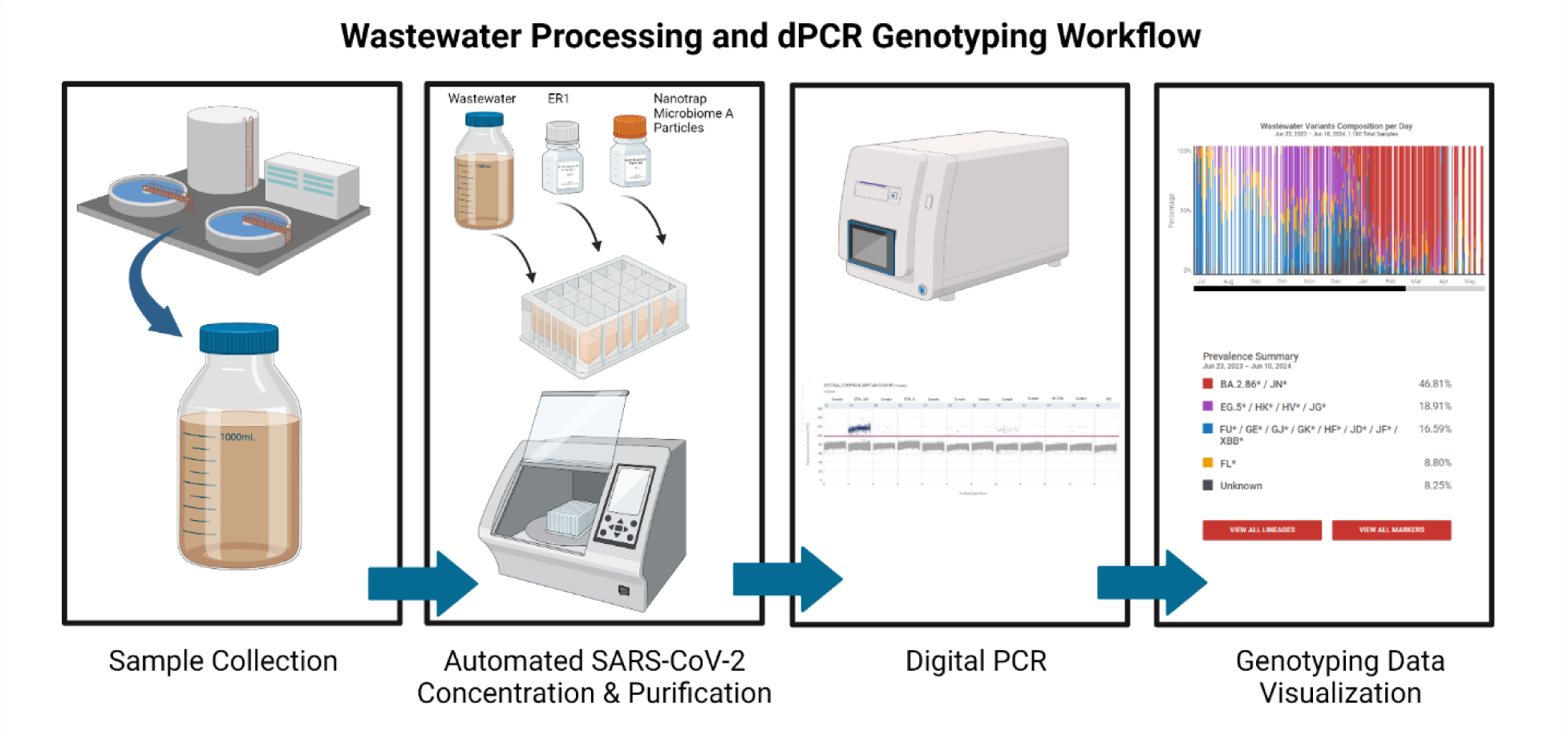
Workflow for wastewater processing and digital PCR genotyping. (1) Sample Collection: Untreated wastewater is collected from treatment facilities. (2) Automated SARS-CoV-2 Concentration & RNA extraction: Samples are concentrated using Nanotrap Microbiome A particles and extracted using MagMAX kit in a KingFisher Apex system. (3) Digital PCR: The samples are then analyzed to detect and quantify SARS-CoV-2 variants. (4) Genotyping Data Visualization: Data visualization shows the composition and prevalence of various SARS-CoV-2 variants over time.

### Wastewater samples

This study processed and analyzed 1416 wastewater samples from six states from April 2023 to May 2024. Table 1 lists the number of samples tested from each state and the collection period. Detailed information on the collection sites and methods is described in the following paragraphs.

**Table 1:** Summary of Wastewater Samples Collected Across Six States. Overview of the total number of wastewater samples collected, along with the start and end dates of sample collection, from each state. The data spans April 11, 2023, to May 16, 2024, across California, Georgia, Illinois, Louisiana, New York, and Wisconsin.

| State | Total wastewater samples | Start | End |
| --- | --- | --- | --- |
| California | 155 | 10/31/2023 | 04/24/2024 |
| Georgia | 528 | 04/11/2023 | 04/05/2024 |
| Illinois | 322 | 10/31/2023 | 05/16/2024 |
| Louisiana | 101 | 02/27/2024 | 04/25/2024 |
| New York | 152 | 01/04/2024 | 04/25/2024 |
| Wisconsin | 158 | 11/22/2023 | 02/01/2024 |

#### Georgia

The Center for Global Safe Water, Sanitation, and Hygiene (CGSW) laboratory at Emory University was the first wastewater testing laboratory to use the dPCR-based wastewater genotyping method. Between April 2023 to April 2024, CGSW analyzed over 500 wastewater samples from Georgia, spanning three distinct genotyping panels referred to as “Panel 1”, “Panel 2”, and “Panel 3”.

During the feasibility phase, CGSW tested a combination of grab, Moore swabs, and composite samples from various locations such as public schools, influent lines at wastewater treatment plants (WWTPs), a county jail, and an airport. This breadth of sample types was utilized to assess the feasibility of dPCR-based wastewater genotyping across diverse locations to evaluate the method’s reliability and robustness.

After the feasibility phase, CGSW transitioned from testing multiple community sites to sampling selected WWTPs in metro Atlanta, as well as selected WWTPs throughout the state participating in the GA National Wastewater Surveillance System (NWSS) program. Testing was exclusively conducted on untreated composite wastewater samples sent to the Georgia Department of Public Health (GDPH) weekly from April 2023 to April 2024 and wastewater samples from metro Atlanta WWTPs collected by the CGSW collection team. Wastewater samples were stored at 4°C before concentration and extraction of SARS-CoV-2, as described in the wastewater processing method section. Extracted RNA samples were preserved at -20°C until digital PCR analysis was performed for SARS-CoV-2 variant detection.

Figure 2 depicts the location and population size served by the wastewater collection sites and the total number of wastewater samples tested for Panel 1, Panel 2, and Panel 3 from April 2023 to April 2024. A total of 528 wastewater samples were tested from these WWTP sites in this period. The wastewater collection sites cover more than two million Georgia residents or approximately 18% of the state’s population. Most of the collection sites were in the Atlanta metropolitan area, reflecting the dense population and urbanization in Fulton, Gwinnett, and Cobb counties. Conversely, rural regions like Dougherty, Laurens, and Treutlen counties feature smaller facilities, corresponding to their sparse populations. The dispersed collection sites throughout various geographical locations ensured extensive coverage of the state of Georgia.

**Figure 2:**
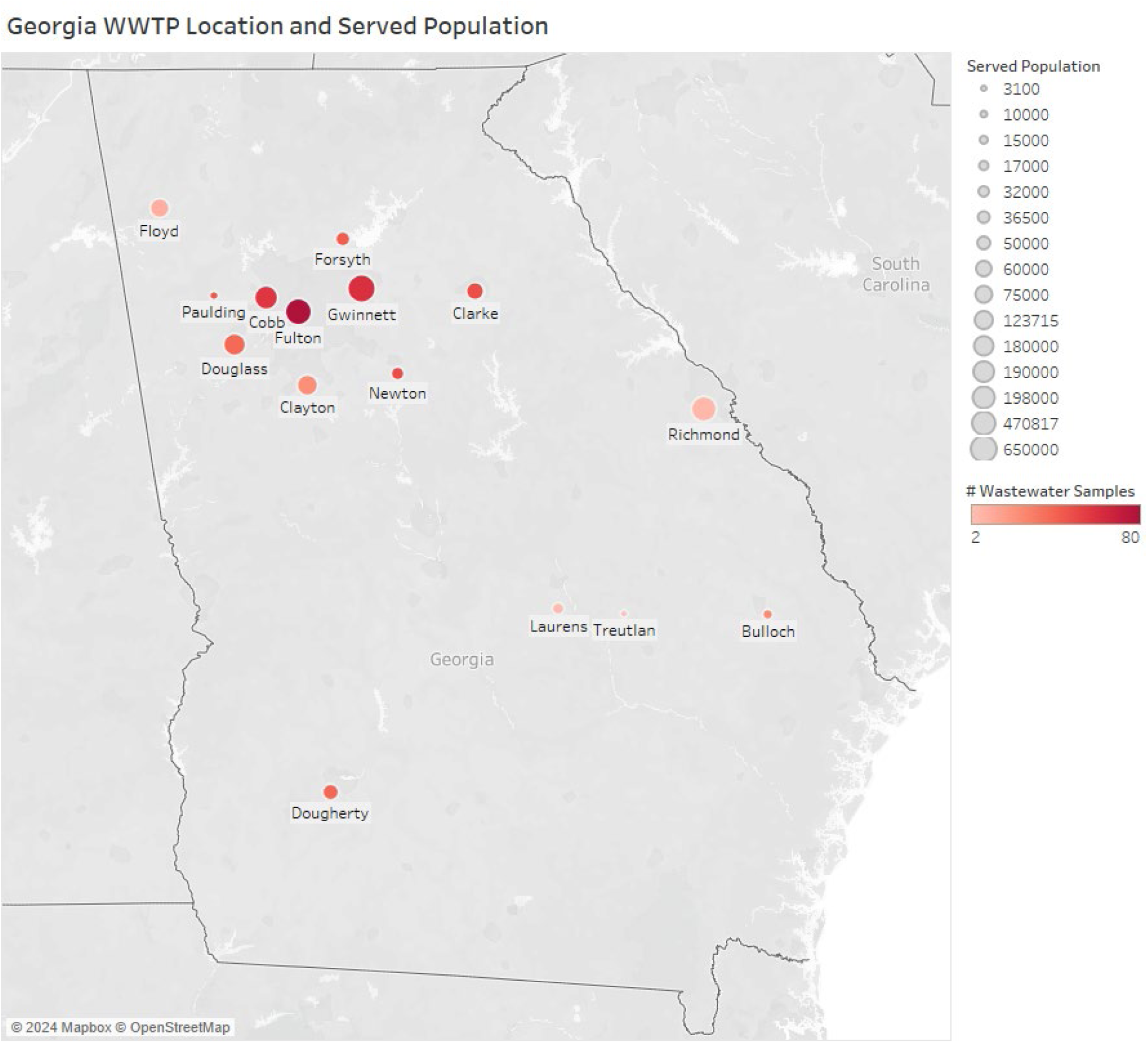
Locations and Population Served by Wastewater Treatment Plants (WWTP) in Georgia. This map illustrates the distribution of 17 Wastewater Treatment Plants (WWTP) across Georgia counties (3 WWTPs in Fulton county) and the populations they serve. The size of each circle represents the served population, with larger circles indicating larger populations. The color intensity, ranging from light to dark red, denotes the number of wastewater samples tested at each location, with darker shades representing higher sample counts. Key areas with significant populations and multiple facilities include Cobb, Fulton, and Gwinnett counties.

#### Wisconsin

For this study, the Wisconsin State Lab of Hygiene (WSLH) collected 158 wastewater samples as part of its regular Wastewater Monitoring Program. Samples were taken from WWTPs across Wisconsin using automated samplers to gather untreated 24-h flow-proportional composite influent samples, with an exception of one site that collected time-weighted 24-h composite samples. Influent composites were sub-sampled into 250 mL bottles and refrigerated prior to overnight shipping on ice. They were processed by WSLH immediately upon arrival or refrigerated at 2-8 ℃ for less than a week before being analyzed. Extracted samples were quantified immediately post-extraction and subsequently stored in a freezer at -80 ℃, except for JN.1 variant analysis which was done retrospectively on previously frozen RNA extracts.

Samples were analyzed from 20 WWTPs (Figure 3), representing more than 1.2 million people, or about 20% of the state’s population. High-density areas, particularly in the southeastern region, including counties like Kenosha, Waukesha, and Jefferson were well represented (Figure 3). However, Wisconsin has a clear regional variation, with northern regions having fewer and smaller WWTPs compared to the more urbanized and densely populated southern areas. The selection of the 20 sites was aimed at comprehensive statewide coverage, while also balancing wastewater treatment needs, support of public health, and environmental sustainability.

**Figure 3:**
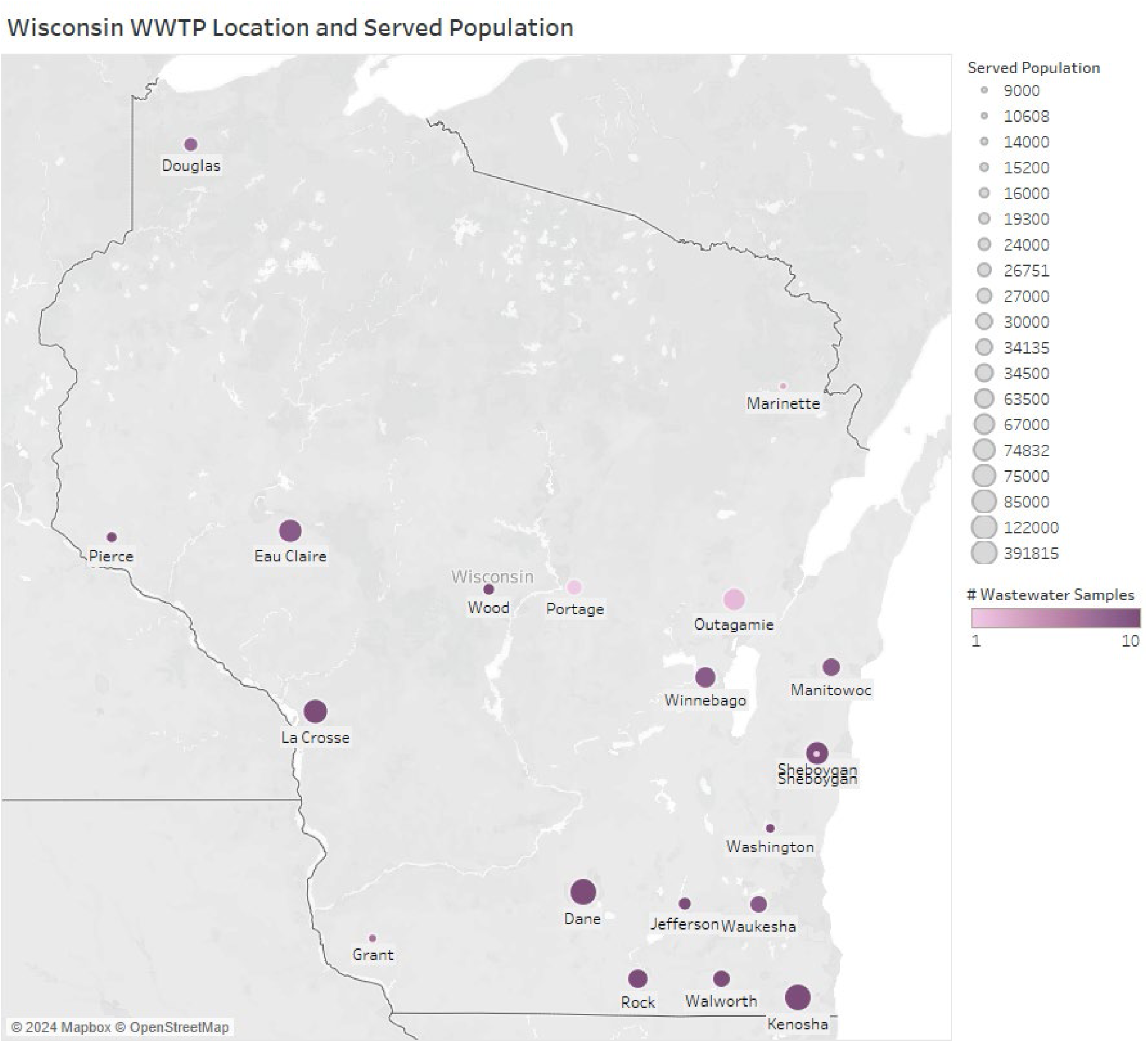
Locations and Population Served by Wastewater Treatment Plants (WWTP) in Wisconsin. This map displays the distribution of Wastewater Treatment Plants (WWTP) across Wisconsin and the populations they serve. The size of each circle represents the served population, with larger circles indicating larger populations. The color intensity, ranging from light to dark purple, denotes the number of wastewater samples tested at each location, with darker shades representing higher sample counts. Key areas with significant populations and multiple facilities include La Crosse, Sheboygan, and Eau Claire counties.

### Wastewater processing - Nanotrap particle method details

Automated target concentration was accomplished using Nanotrap® Microbiome A Particles and Enhancement Reagent 1 (ER1) on a Thermo Scientific™ KingFisher Apex System. In brief, 75 μl of Nanotrap Microbiome A particles and 50 μl of Nanotrap® Enhancement Reagent 1 were mixed with ∼5 ml of wastewater in two replicate wells with a total of 10 ml of wastewater for each sample. The automated script can process up to 24 samples at once in 1 hour. Concentrated viruses were lysed in 500 μL Microbiome Lysis Buffer at 56 ℃.

After viral concentration, the samples were processed for RNA extraction using the Applied Biosystems MagMAX™ Wastewater Ultra Nucleic Acid Isolation Kit on the KingFisher Apex System. Briefly, 400 μL of lysate were mixed with 550 μL of MagMAX binding mix. 10 μL ProK was added to each sample prior to running on the KingFisher Apex. Wash 1 and wash 2 were prepared by adding 1 mL of MagMAX wash solution and 80% ethanol respectively. RNA was eluted in 100 μL Microbiome Elution Solution.

### Genotyping assays and methods

SARS-CoV-2 genotyping was accomplished using the Applied Biosystems™ TaqMan™ SARS-CoV-2 Mutation Panel (Cat# A49785). Thermo Fisher Scientific provides users the option of ordering different variant detection assays. Each assay utilizes TaqMan™ 5’ nuclease chemistry to amplify and detect specific variants within purified genomic RNA samples. At the core of each assay are sequence-specific forward and reverse primers. Key to the panel’s specificity are two TaqMan™ probes equipped with minor groove binders (MGBs), nonfluorescent quenchers (NFQs), and distinct 5’ reporter dyes: one labeled with VIC™ dye for the reference sequence, and another with FAM™ dye for variant detection. The reference allele is based on the WHO-classified Wuhan strain, and the mutant allele is a target single nucleotide polymorphism (SNP) mutation. This dual-probe approach allows for precise differentiation between the reference and mutant alleles. Table 2 summarizes the assays and targeted mutation sites used in this study:

**Table 2:** Assay and mutation sites for SARS-CoV-2 genotyping. This table lists the assays used for detecting specific SARS-CoV-2 variants based on their mutations. Each row includes the ThermoFisher Assay ID, the specific mutation it targets, and the variants and sublineages detected. For example, assay ANCFPZX targets the G8393A mutation and is used for identifying the JN variants. Panels of 4 assays are used in combination in order to monitor the prevalence of different SARS-CoV-2 variants and. The genotyping panels evolve in time to follow the virus mutations.

| Assay ID | Mutation | Variants | Period |
| --- | --- | --- | --- |
| CV9HHWW | G28681T | BQ* | Jan 2023 - Aug 2023 |
| CV32Z67 | A19326G | XBB.* | Jan 2023 - now |
| CVXGPWV | C28928T | EG.1* | Aug 2023 - Dec 2023 |
| CVU62R2 | C29625T | EG.5* | Jan 2024 - now |
| CVEPRZU | G5720A | FL* and EG* | Aug 2023 - now |
| ANCFPZX | G8393A | JN*, KP*, LB* | Jan 2024 - now |

Figure 4 illustrates the evolutionary relationships of SARS-CoV-2 variants as defined by Nextstrain clades. Starting from the 20B (B.1.1) lineage, it traces the progression and diversification of variants, including significant nodes like 21M (Omicron, B.1.1.529), which branches into sub-lineages such as 21K (Omicron, BA.1) and 21L (Omicron, BA.2). The genotyping assays in Table 2 targeted six variants in the 21L (Omicron, BA.2) clade. The tree uses color coding to differentiate between major variants and sub-variants.

**Figure 4:**
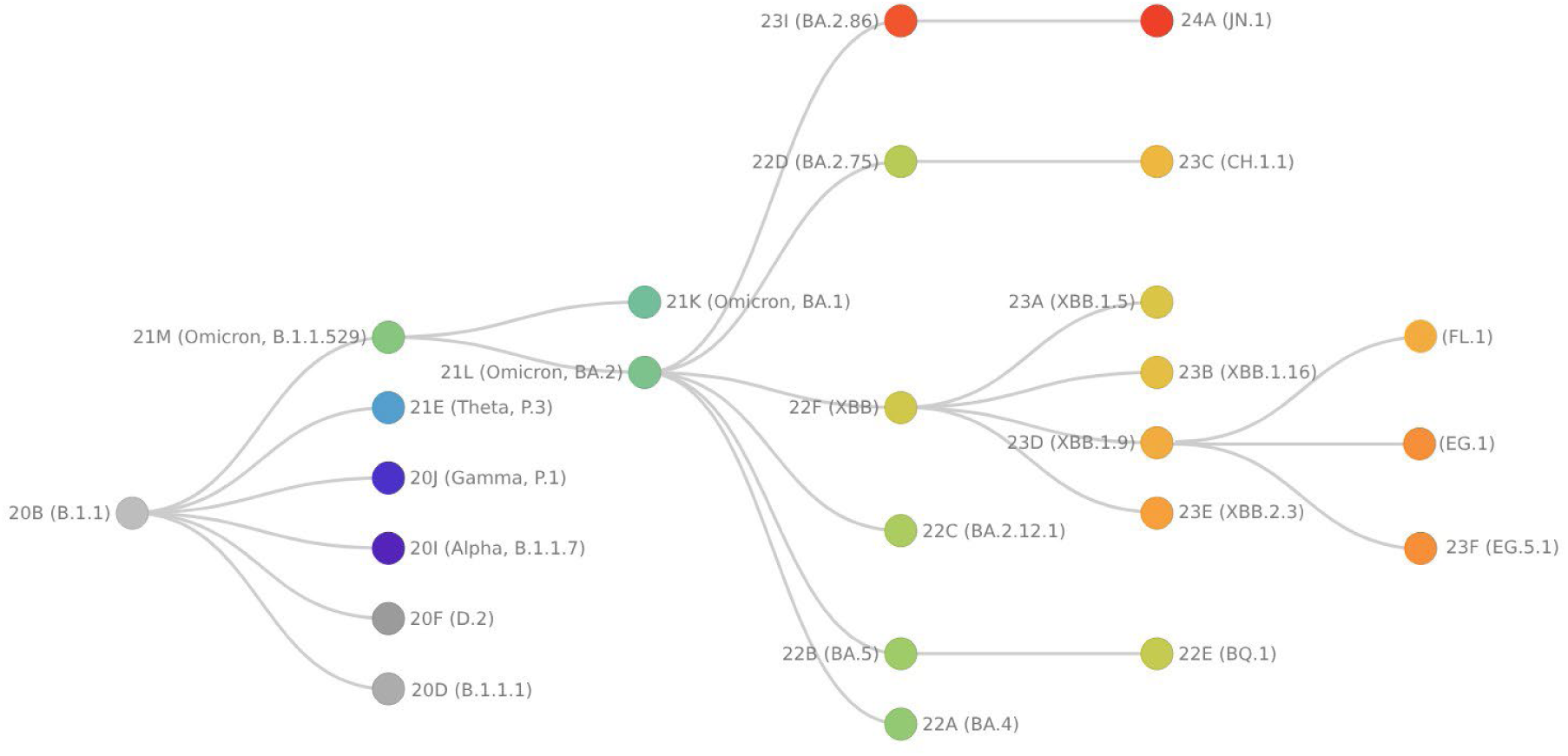
Phylogenetic Tree of SARS-CoV-2 Variants. This figure illustrates the evolutionary relationships between SARS-CoV-2 variants as defined by Nextstrain clades. Genotyping assays in Table 2 target six variants in the 21L clade. The phylogenetic tree was adapted from Nextstrain’s ncov-clades-schema with some modifications (33,34).

### Marker selection

The marker selection for SARS-CoV-2 variant detection and lineage assignment methods used in this study have been previously described (25).

Sequences for positive controls (mutation and wild-type) were designed in-silico by ROSALIND Bio and were constructed and manufactured using the gBlocks service by IDT or the GeneArt service by ThermoFisher Scientific. Assays and controls were validated by Emory University and Ceres Nanosciences, Inc. using the QIAcuity dPCR system.

### Variant detection panels

In this one-year study, three variant genotyping panels were utilized, with new panels deployed as the relative prevalence of different SARS-CoV-2 variants changed in the United States.

#### **i-** Panel 1

Table 3 shows the variants of concern (VOC) and associated SNP sites in Panel 1:

**Table 3:**
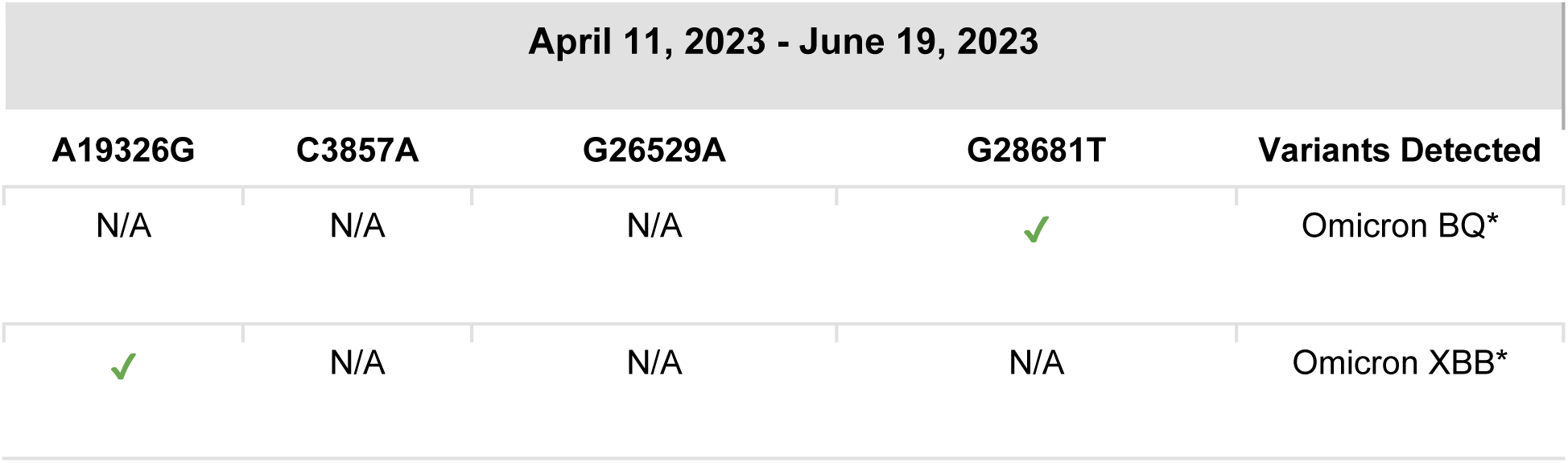
Panel 1 SARS-CoV-2 Variant Classification Based on Mutations (April 11, 2023 - June 19, 2023). Presence of G28681T mutation identifies Omicron BQ* variant, while A19326G mutation identifies Omicron XBB* variant. N/A indicates mutations not applicable.

| April 11, 2023 - June 19, 2023 |  |  |  |  |
| --- | --- | --- | --- | --- |
| A19326G | C3857A | G26529A | G28681T | Variants Detected |
| N/A | N/A | N/A | ✓ | Omicron BQ* |
| ✓ | N/A | N/A | N/A | Omicron XBB* |

Emory University analyzed 86 wastewater samples from multiple sites in Georgia in this period. The assays were run on a QIAcuity dPCR platform, and the data were uploaded to the ROSALIND TRACKER.

#### **ii-** Panel 2

Table 4 shows the VOCs and SNP sites in Panel 2:

**Table 4:** Panel 2 classification of SARS-CoV-2 Variants by Mutations (June 21, 2023 - February 18, 2024). The table classifies SARS-CoV-2 variants based on the presence (✓) or absence (✗) of specific mutations. Variants are classified as EG.1* with mutations A19326G, C28928T, and G5720A, FL* with mutations A19326G and G5720A, XBB*/JD*/JF*/GK*/GJ*/FU*/HF*/GE* with mutation A19326G, and EG.5*/JG*/HK*/HV* with mutations A19326G, C29625T, and G5720A.

| June 21, 2023 - February 18, 2024 |  |  |  |  |
| --- | --- | --- | --- | --- |
| A19326G | C29625T | C28928T | G5720A | Variants Detected |
| ✓ | X | ✓ | ✓ | EG.1* |
| ✓ | X | X | ✓ | FL* |
| ✓ | X | X | X | XBB*/JD*/JF*/GK*/GJ*/FU*/HF*/GE* |
| ✓ | ✓ | X | ✓ | EG.5*/JG*/HK*/HV* |

Initially, Only samples for Georgia were tested in this panel. In November 2023, the testing capability was expanded to include five more states (New York, Wisconsin, Illinois, California, and Louisiana).

#### **iii-** Panel 3

Panel 3 was designed in response to the rise of JN variants in wastewater and clinical samples. Analysis of Georgia wastewater genotyping data between panel 2 period showed that prevalence of the EG1 variant had dropped to <1%. Therefore, the assay to detect EG.1 was retired from the genotyping panel and replaced by the JN variant assay. Table 5 shows the VOCs and SNP sites in the variant genotyping Panel 3:

**Table 5:** Classification of SARS-CoV-2 Variants by Mutations on Panel 3 (February 19, 2024 - Present). BA.2.86*/JN* identified by G8393A; FL* by A19326G and G5720A; XBB*/JD*/JF*/GK*/GJ*/FU*/HF*/GE* by A19326G; EG.5*/JG*/HK*/HV* by A19326G, C29625T, and G5720A.

| February 19, 2024 - Present |  |  |  |  |
| --- | --- | --- | --- | --- |
| A19326G | C29625T | G8393A | G5720A | Variants Detected |
| X | X | ✓ | X | BA.2.86*/JN*/KP*/KS*/KV* / LB* |
| ✓ | X | X | ✓ | FL* |
| ✓ | X | X | X | XBB*/JD*/JF*/GK*/GJ*/FU*/HF*/GE* |
| ✓ | ✓ | X | ✓ | EG.5*/JG*/HK*/HV* |

### Digital PCR

The mutation detection assay was performed on the QIAGEN QIAcuity Digital PCR system. In brief, the reaction mix was made by mixing 10 μL OneStep Advanced Probe Master Mix and 0.4 μL OneStep Advanced RT Mix. The volume brought up to 30 μL by adding RNase-free water. Ten μL of RNA template was added to the reaction mix, then the entire volume of 40 μl was transferred into 26K 24-well Qiagen Nanoplate. The QIAGEN QIAcuity Digital PCR system was used to amplify and detect the signals. Amplification was accomplished according to the following steps: 1-One cycle at 50°C for 40 minutes, 2-One cycle, at 95°C for 2 minutes, 3-Forty-five cycles, at 95°C, for 3 seconds, then 60°C for 30 seconds. Only for the FL assay, step 3 was modified to forty-five cycles, at 95°C, for 30 seconds, then 57°C for 1 minute. Signal detection was obtained using default settings for exposure duration and gain in each channel. QIAcuity Software Suite (ver 2.2) was used to analyze the data. A common threshold was applied across the samples to clearly separate negative partitions from positive partitions. Mutation detection results were exported in CSV format.

### Uploading data to ROSALIND Dashboard

To convert the dPCR assay data into variant genotyping results, the data was uploaded to the ROSALIND Tracker platform (https://app.rosalindai.bio) for analysis. This involved uploading two types of files: dPCR CSV files, which were exported from the QIAcuity instrument, and metadata CSV files, which provide information about each sample, such as its origin and collection date. Once both the PCR data and metadata were uploaded, the ROSALIND Tracker calculated variant percentages for each sample and presented the results in a publicly-available dashboard on the [ROSALIND Tracker website](https://tracker.rosalind.bio/tracker/dashboard/?date-range=Last+90+Days&selected-date=2024-07-19&graph-display=percentage&metric=cases&graph-controls=cases&graph-controls=hospitalizations&graph-controls=deaths&graph-controls=positivity&graph-daily-weekly=daily&selected-lab=all-labs).

### Calculating variant percentages

Each genotyping assay was designed to detect a specific single nucleotide mutation and the corresponding wild type sequence in the SARS-CoV-2 genome. Using digital PCR, the mutation fraction was calculated using the formula below:

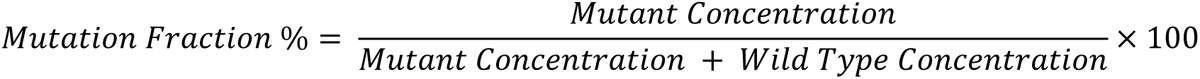

As SARS-CoV-2 evolved, combinations of mutations led to the emergence of different lineages and variants. For instance, the XBB variant isa recombinant lineage derived from the BA.2.10.1 and BA.2.75 sublineages of Omicron and includes the A19326G mutation (35,36). Another example is the EG.5 variant, which has acquired a set of mutations, including A19326G from XBB and C29625T, illustrating how SARS-CoV-2 can accumulate advantageous mutations from various lineages to potentially enhance its survival and spread (37).

To monitor the dynamics of SARS-CoV-2 variants in wastewater, we tracked specific mutations using a combination of genotyping assays. This strategy enables the detection of unique mutations associated with specific lineages through direct detection. If a mutation is shared among multiple variants, the mutation fraction was determined by subtracting the fractions of other mutations. These adjustments were made in a pre-defined order to estimate the fraction of each variant detected in the wastewater samples. The sum of variant prevalence should be close to 100% in each sample. To account for assay performance differences, we applied a normalization step if the total value was at or above 95%. For example, if the total prevalence was 97%, each variant prevalence was normalized by dividing by 0.97. Additionally, if there were undetected variants in the sample, the total prevalence will be below 95%. The percentage of undetected variants was obtained by subtracting the total mutation fraction in the panel from 100%.

Table 6 illustrates the SNP assays and associated lineages in panel 2. Assay 4 and assay 3 uniquely target the EG.5 and EG.1 lineages, respectively. Assay 2 detects the G5720A mutation, common to the FL, EG.1, and EG.5 lineages within this panel. To calculate the mutation fraction for the FL variant, we subtracted the EG.1 and EG.5 mutation fractions from the result of assay 2. Similarly, assay 1 targets mutation A19326G, which is shared by all four variants in this panel. By estimating the mutation fractions of EG.5, EG.1, and FL variants from the outcomes of assays 4, 3, and 2 respectively, we could then determine the XBB mutation fraction by subtracting these fractions from the result of assay 1. Table 6 shows an example of mutant fraction and variant fraction calculated using a mock wastewater sample in panel 2:

**Table 6:** Mutation Detection and Fraction Analysis for SARS-CoV-2 Variants. This table shows the detection of specific mutations in various SARS-CoV-2 lineages using different assays in Panel 2. Assay 3 (C28928T) and Assay 4 (C29625T) are specific to EG.1 and EG.5, respectively. The first step is to calculate the FL variant fraction by subtracting the results of Assays 3 and 4 from Assay 2. Following this first step, the XBB variant fraction is calculated by subtracting the results of Assays 2 (now corrected from previous step), 3, and 4 from Assay 1. For example, the Assay 1 (A19326G) has a mutant concentration of 70 Cp/μL and a wild-type concentration of 100 Cp/μL, resulting in a 41% mutant fraction and a 3% adjusted variant fraction after subtracting the contributions of Assays 2, 3, and 4 (38%). The unknown variant fraction is calculated by subtracting the combined known fractions from 100%. In this example, the unknown variant(s) show a notably high adjusted variant fraction of 59%.

| Panel 2 | Example of Genotyping Assay Result |  |  | Assigned Lineage/Variant |  |  |  |  |
| --- | --- | --- | --- | --- | --- | --- | --- | --- |
|  | Mut (Cp/μL) | WT (Cp/μL) | Mutant Fraction | XBB | FL | EG.1 | EG.5 | Unknown |
| Assay 1 (A19326G) | 70 | 100 | 41% | ✓ | ✓ | ✓ | ✓ | NA |
| Assay 2 (G5720A) | 60 | 100 | 38% | × | ✓ | ✓ | ✓ | NA |
| Assay 3 (C28928T) | 20 | 100 | 17% | × | × | ✓ | × | NA |
| Assay 4 (C29625T) | 10 | 100 | 9% | × | × | × | ✓ | NA |
| Variant Fraction |  |  |  | 3% | 12% | 17% | 9% | 59% |

A classification algorithm was developed by [ROSALIND](https://www.rosalind.bio/) to automate the mutation fraction analysis in each sample based on the input from digital PCR CSV files and assigned panels. A dedicated system was established to host the classification algorithm and to provide a web application with an application programming interface for standardized data submission and processing. This system was deployed on a secure virtual private cloud instance on Google Cloud Platform, enabling the processing of thousands of specimens per minute.

### Presenting genotyping results on the ROSALIND dashboard

The prevalence of the various genotypes detected in wastewater samples over time was presented on the ROSALIND dashboard. The dashboard allows users to select a specific time period, filter results by state or other criteria, and view data summaries by specimens or percentages. Users can explore all lineages and active marker sets with tooltips providing additional information. The USA map features a tooltip showing variant distribution for selected states, with detailed site information such as sewershed ID, sample location, jurisdiction, county, population served, and variant distribution.

In addition to the wastewater surveillance results, the dashboard also presents data on the variants detected in clinical specimens during the same time period. This aspect focuses on genomic surveillance of pathogens isolated from clinical samples, such as those collected from individuals presenting with symptoms or seeking medical care. The same genotyping PCR assays, designed for specific mutations, are used for both wastewater and clinical samples (25,32).

The dashboard also includes prevalence information on SARS-CoV-2 variants over time obtained from sequencing data on clinical isolates which offers a more detailed perspective on SARS-CoV-2 genomic surveillance efforts. The sequence visualization is based on data from [GISAID](https://gisaid.org/). Powered by data from [Covid ActNOW](https://covidactnow.org/?s=50083753), the ROSALIND dashboard offers detailed visualization tools for monitoring COVID-19 metrics, including cases, hospitalizations, deaths, and test positivity rates.

Figure 5 shows a screenshot of the ROSALIND tracker on July 24, 2024. 1,416 wastewater samples from six different states were processed and analyzed using dPCR genotyping approach from April 1, 2023 to May 31, 2024. In comparison, 15,416 clinical samples nationwide were analyzed using this same assay but with qPCR-based genotyping, and 316,602 clinical sequencing results were deposited in the GISAID database. A screenshot of the ROSALIND tracker is shown in figure 5. For more detailed information, visit the [ROSALIND Tracker](https://tracker.rosalind.bio/tracker/dashboard/?date-range=Last+90+Days&selected-date=2024-07-19&graph-display=percentage&metric=cases&graph-controls=cases&graph-controls=hospitalizations&graph-controls=deaths&graph-controls=positivity&graph-daily-weekly=daily&selected-lab=all-labs).

**Figure 5:**
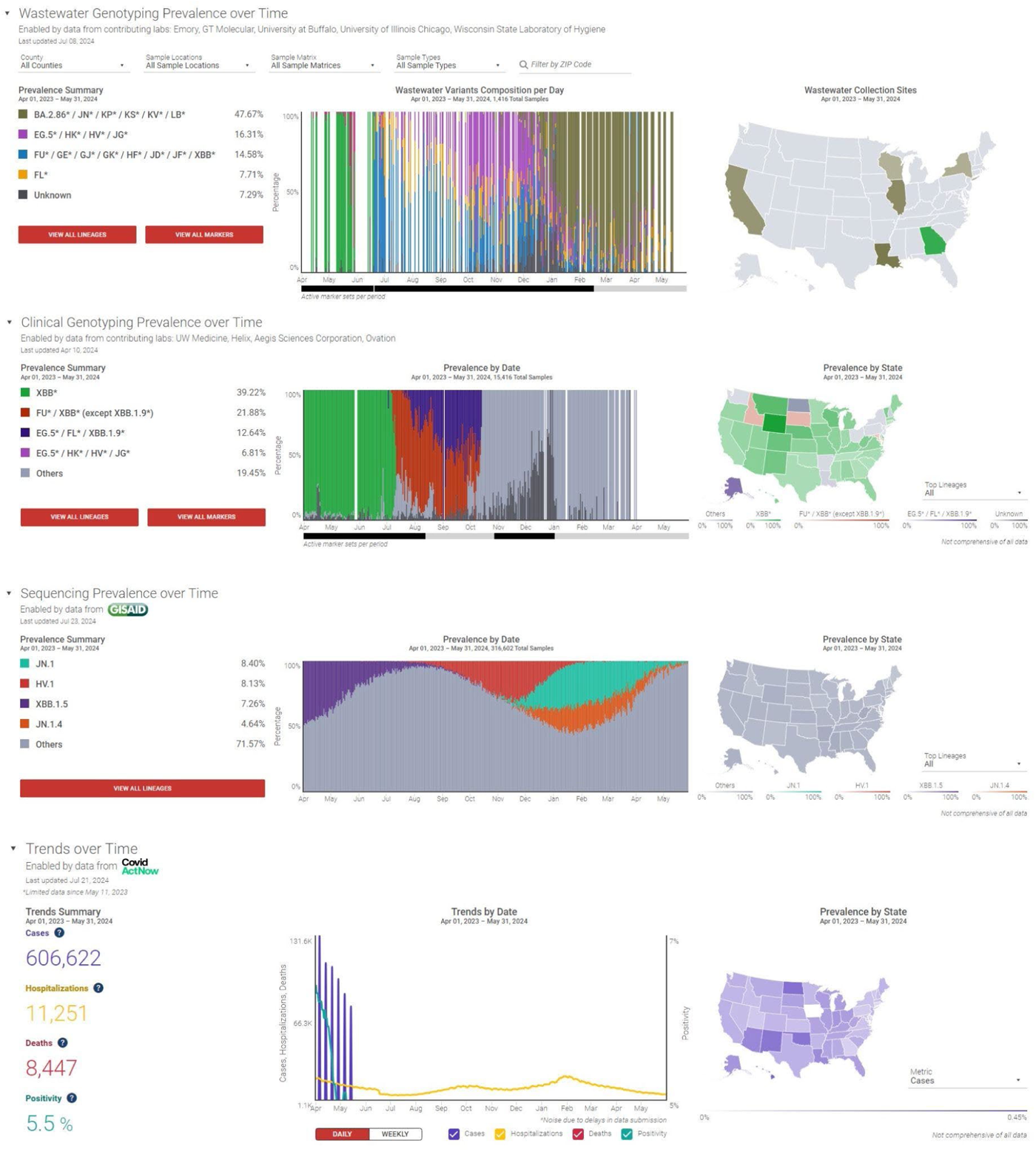
A screenshot of the ROSALIND Tracker for the period April 1, 2023, to May 31, 2024. The tracker displays the results of wastewater genotyping, clinical sample genotyping, and GISAID sequencing data for SARS-CoV-2 variants. The dashboard offers detailed visualization tools for monitoring COVID-19 metrics, including cases, hospitalizations, deaths, and test positivity rates.

### Variant sequencing methods

#### a. Emory University

For wastewater samples processed during the feasibility phase at Emory University, samples were also sequenced using the following protocol: Selected SARS-CoV-2 positive RNA samples were converted into cDNA using the SuperScript™ IV First-Strand Synthesis System and amplified with the ARTIC V4.1 nCOV-2019 Amplicon Panel kit, which uses 98 primers in two pools to detect mutations. Two separate PCR reactions were performed on each cDNA sample to ensure comprehensive amplification, followed by purification and preparation of next-generation sequencing (NGS) libraries. Low-quality libraries (≤20 ng/µL) were excluded. SARS-CoV-2 positive NGS libraries were sequenced using the NovaSeq6000 system, targeting 1 million reads per sample. Sequencing data was aligned to the Wuhan-Hu-1 genome and processed to remove ARTIC primer sequences, with insufficient depth samples excluded. The Freyja pipeline calculated the relative abundance of SARS-CoV-2 lineages, and variants detected at less than 0.01% were not reported. Lineages were categorized into Pangolin variants of concern, with rare variants grouped as “other,” and averaged by collection date and location to determine final relative abundances.

#### b. Wisconsin State Laboratory of Hygiene

WSLH utilized a different library prep method for sequencing using the following protocol. The SARS-CoV-2 libraries were prepared using the QIAseq DIRECT SARS-CoV-2 Enhancer kit using the Booster primers (Qiagen). Briefly, single-stranded viral RNA molecules were reverse transcribed into cDNA using hexaprimers. SARS-CoV-2 genome was then specifically enriched using a SARS-CoV-2 primer panel. The panel consists of approximately 550 primers for creating 425 amplicons, covering the entire SARS-CoV-2 viral genome. Prior to sequencing, library quality was assessed using the QIAxcel Advanced System (Qiagen) and quantified by qPCR using the QIAseq Library Quant System kit (Qiagen). Libraries were sequenced on a MiSeq Illumina platform using MiSeq Reagent v2 (300 cycles) kits targeting a median coverage of at least 500X and at least 80% of the genome covered at 10X.

To assess variant proportions, whole genome sequencing (WGS) data was analyzed using Freyja v.1.3.11, a tool specifically designed to estimate SARS-CoV-2 variant proportions in deep sequence data containing mixed populations (see reference: 10.1038/s41586-022-05049-6). BAM files, generated using viralrecon v2.5 (https://nf-co.re/viralrecon/), were processed through Freyja, utilizing the Wuhan-Hu-1 reference genome (MN908947.3) to produce variant and depth files. The median estimates were obtained through Freyja’s bootstrap boot function (nb = 10). All samples were processed using Freyja’s UShER barcode reference database updated on April 13th, 2024. This ensured the inclusion of all most recent variants detection in samples processed during earlier periods.

Variant proportions derived from Illumina sequencing are accessible through a publicly available dashboard hosted at https://dataportal.slh.wisc.edu/sc2-ww-dashboard. This dashboard showcases the proportions of major variant groups listed on https://covariants.org/, with estimations generated using Freyja’s raw calculations. Additional details on the methodology can be found at https://github.com/wslh-ehd/sc2_wastewater_data_analysis. Raw sequencing data have been deposited into the NCBI repository under the Bioproject PRJNA889839.

## Results

Figure 6 outlines the milestones aimed at advancing the validation and implementation of various genotyping panels for wastewater testing. The project began in January 2023, focusing on adapting previously reported qPCR-based SARS-CoV-2 genotyping assays in clinical samples to wastewater for BA.1 and BA.2. This was followed by validating genotyping assays in the digital PCR system (BQ.1 and XBB). The SOP for Panel 1 (BQ.1 and XBB) was released in April 2023. Validation for new assays for EG, FL, and FD variants began in June 2023. In November 2023, the SOP for Panel 2 (EG.1, EG.5, FL, XBB) was released, and four new testing labs started testing wastewater samples, expanding the testing sites to six states. In December 2023, validation of new assays for HV and JN variants was conducted, culminating in the release of the SOP for Panel 3 (JN, EG.5, FL, XBB) by February 2024. The details, challenges, and outcomes of each phase are described in the next section.

**Figure 6:**
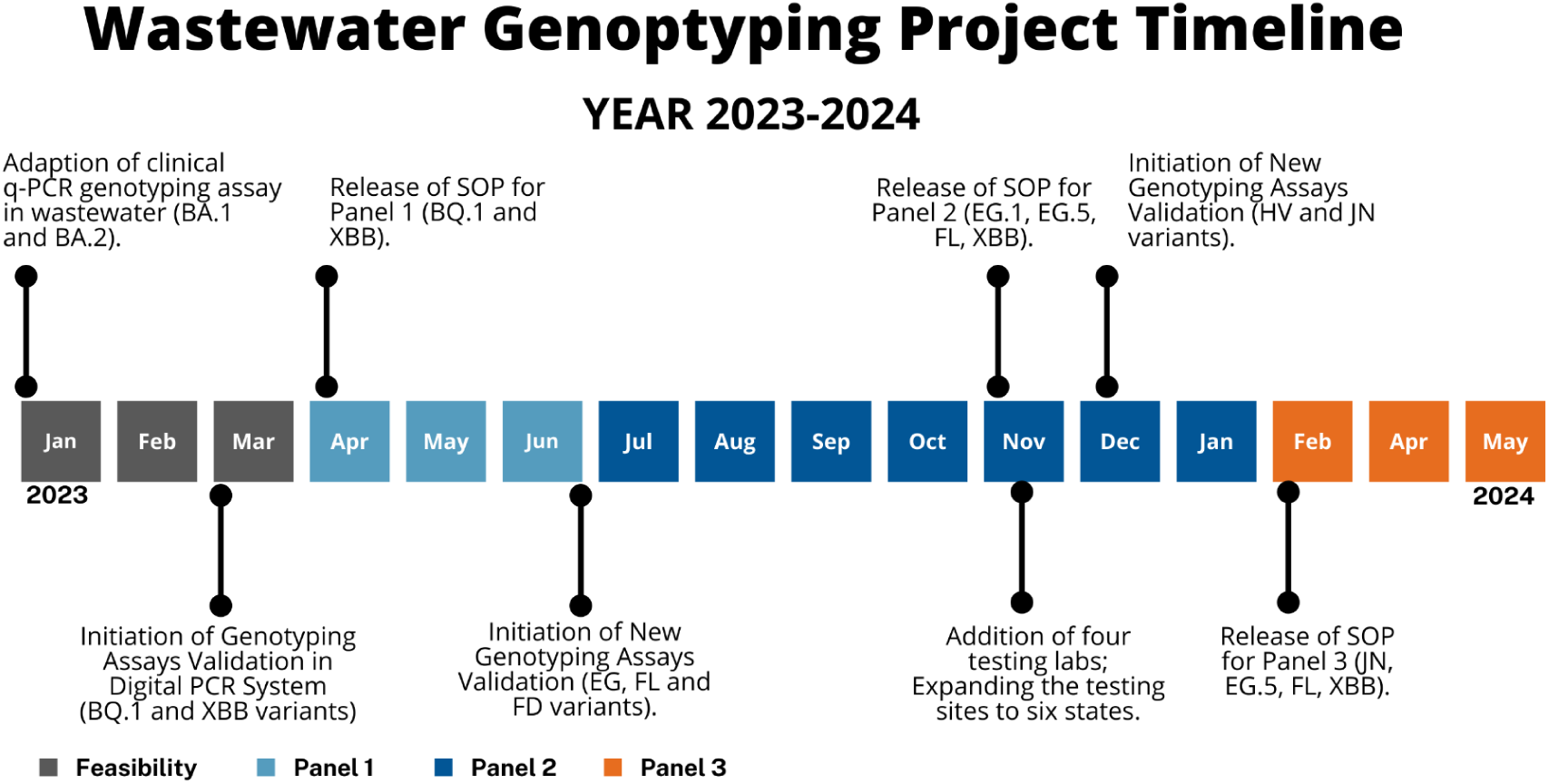
Study Timeline for the dPCR genotyping project outlining key milestones in the development and validation of genotyping assays as well as panels implementation.

### Adapting qPCR-Based SARS-CoV-2 Genotyping from Clinical Samples to Digital PCR for Wastewater Analysis

#### A- BA.1 and BA.2 RT-qPCR genotyping in wastewater vs. whole genome sequencing of viral genomes from wastewater samples

First, we assessed the viability of using the pre-existing ROSALIND qPCR-based genotyping approach (which was being utilized to conduct genotyping for clinical samples) to wastewater samples by analyzing archived nucleic acid extracts from wastewater samples previously processed utilizing the Nanotrap Particle method as well as various variant homogenous and heterogenous controls.

Ten archived nucleic acid extracts from processed wastewater samples that were collected between mid-February 2022 and mid-April 2022 were tested for BA.1 and BA.2 lineages using RT-qPCR genotyping assays (Cat# A49785) on QuantStudio according to the manufacturer’s recommendations.The BA.1 and BA.2 fractions in the wastewater samples were calculated using the ROSALIND automated pipeline originally developed for clinical samples. The same wastewater samples were also analyzed by NGS sequencing. Both methods consistently detected BA.1 and BA.2, with BA.1 prevalence decreasing and BA.2 increasing over time, becoming the dominant variant by early April 2022 (Figure 7). The “Unknown” category in qPCR genotyping and the “Others” category in NGS, present briefly in February and March, respectively, disappear by April, indicating improved detection and classification accuracy or suggesting the presence of other variants or sequencing artifacts that are later resolved. The results from both techniques are comparable, demonstrating the feasibility of qPCR based genotyping for monitoring SARS-CoV-2 variants in wastewater.

**Figure 7:**
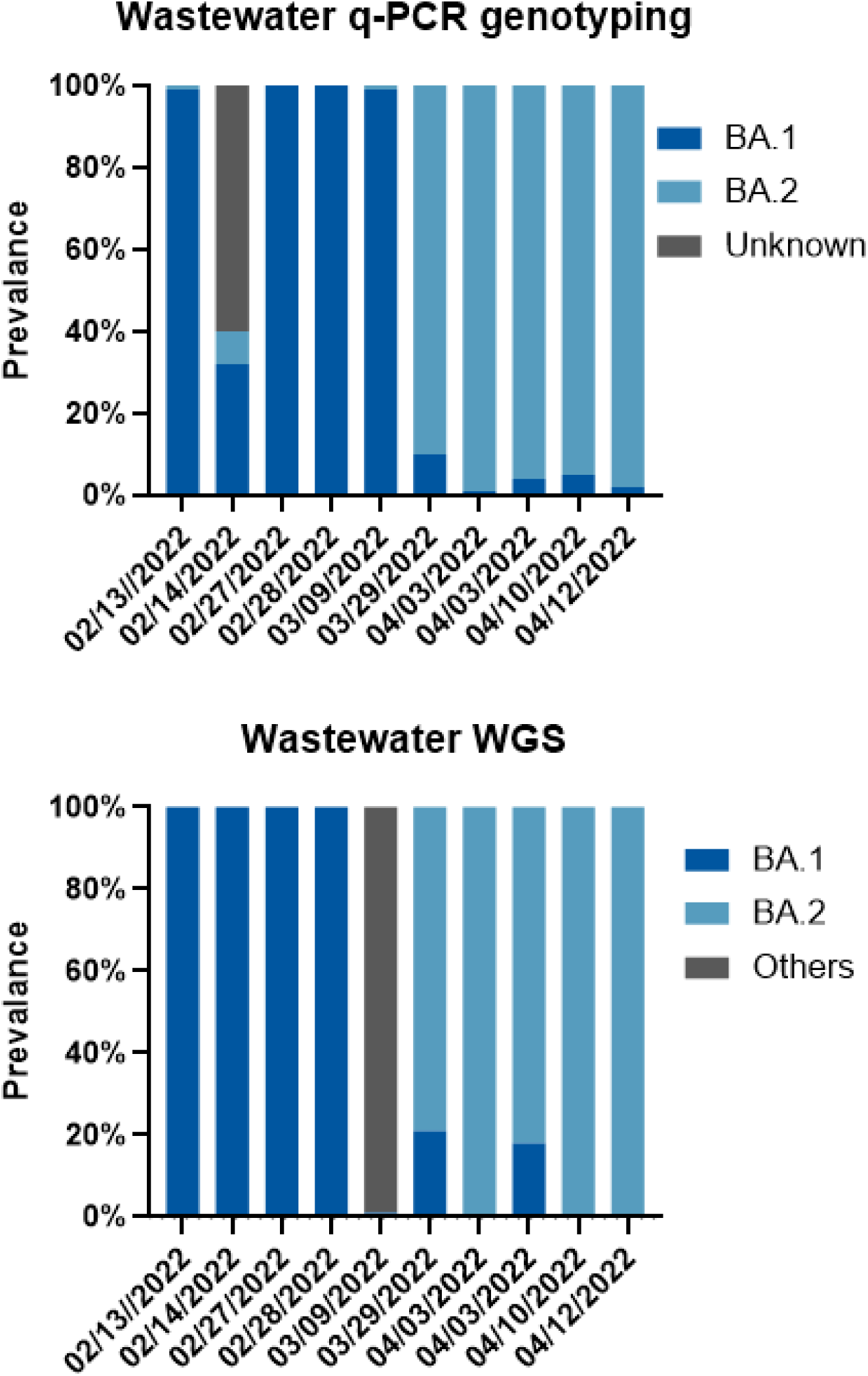
Comparison of SARS-CoV-2 variant prevalence in wastewater samples using qPCR genotyping (top) and WGS (bottom) methods. Both methods consistently detected BA.1 and BA.2 lineages from mid-February to mid-April 2022, showing a decrease in BA.1 and an increase in BA.2 prevalence over time. The results demonstrate the robustness and reliability of both techniques for monitoring SARS-CoV-2 variants in wastewater.

#### B- BQ.1 and XBB dPCR genotyping vs. qPCR genotyping in wastewater samples

Next, we utilized BQ.1 and XBB SARS-CoV-2 variant assays (Cat # CV9HHWW and Cat # CV32Z67, respectively) on the QIACuity Digital PCR platform to evaluate the feasibility of using relative quantitation to determine the presence of variants in wastewater during both unique and transition variant periods. For dPCR, we relied on the proportions calculated by the dPCR software and made necessary adjustments as explained in the “Calculating Variant Percentages’’ section. The ROSALIND team designed a new pipeline to calculate variant prevalence from digital PCR files. The variant fractions in the qPCR method were calculated using the ROSALIND automated pipeline, originally developed for clinical samples and adapted for wastewater use. While clinical samples typically contain one lineage per sample, co-infections were observed regularly (25), allowing us to see the proportion of each variant in patient samples. Based on this, we updated our clinical qPCR pipeline to work with wastewater samples and predict the proportion of the main variants in the mix. To achieve this, in addition to the 100% wild type and 100% mutant controls, we added mixtures of these controls to correctly calibrate our proportion measurements. Figure 8 summarizes the genotyping results for nine wastewater samples collected between mid-January 2023 and early March 2023 using both methods.

**Figure 8:**
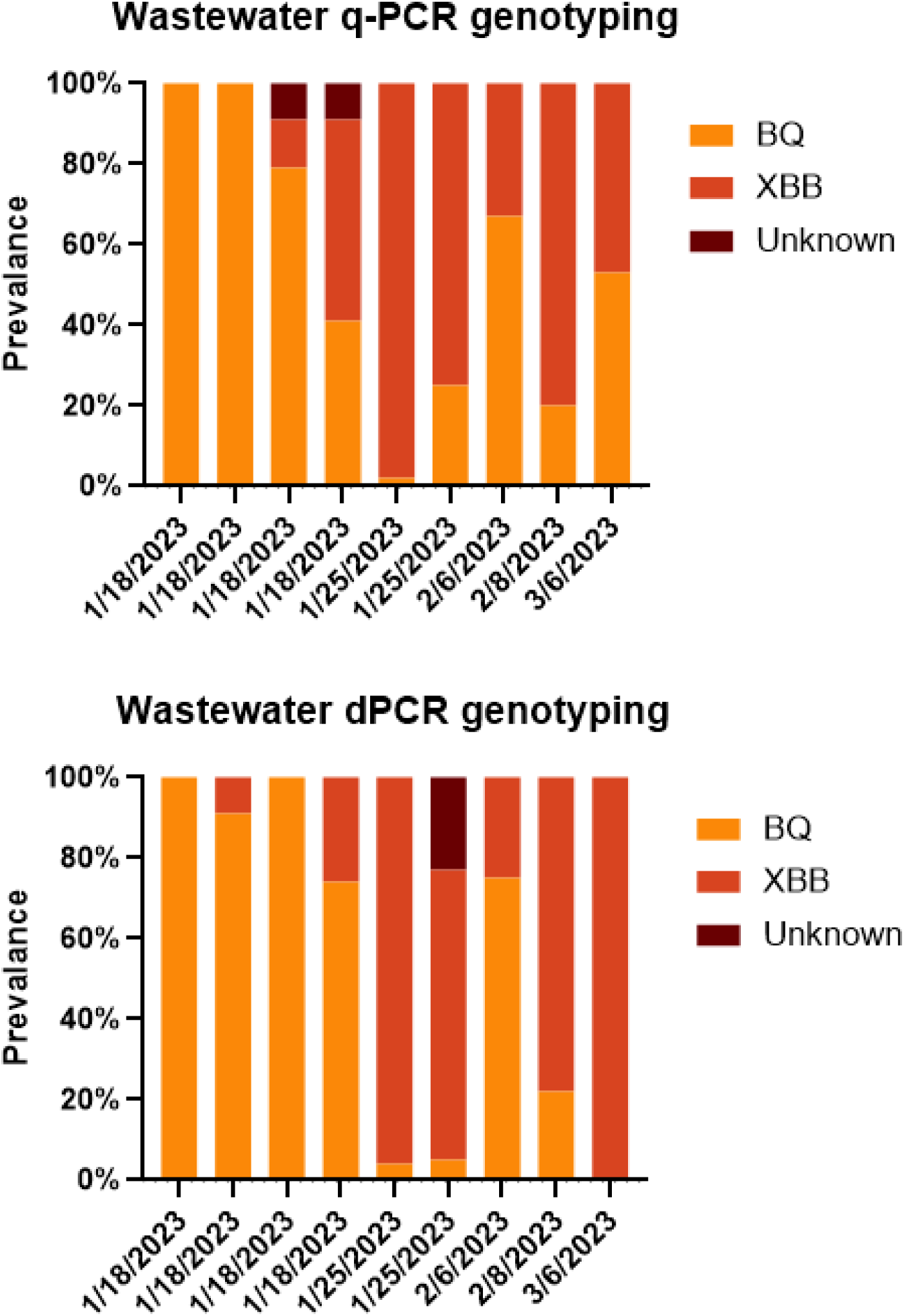
Comparison of SARS-CoV-2 variant prevalence in wastewater samples using qPCR genotyping (top) and dPCR genotyping (bottom) methods for nine samples collected between January to March 2023. Both methods consistently detect BQ (yellow) and XBB (orange) variants, with similar patterns of prevalence across samples. A transition from BQ to XBB variants is observed during this period. The results demonstrate the robustness and reliability of both qPCR and dPCR techniques for monitoring SARS-CoV-2 variants in wastewater.

The data shown in the lower image of Figure 7 depicts the relative amount of XBB and BQ.1 SARS-CoV-2 variants present in each wastewater sample as determined using the dPCR assay. The relative amount of XBB approaches 100% over the time period. In contrast, the relative amount of BQ.1 decreases to nearly 0% over the time period.

Wastewater samples collected between mid-February 2022 and mid-April 2022 showed a rapid displacement of BA.1 with BA.2, and samples collected between mid-December 2022 and early-March 2023 demonstrated a clear transmission from BQ.1 to XBB. These sub-lineage displacements detected by PCR assays were concordant with these sub-lineage dynamics in clinical samples reported in the literature (38–40). These results suggest that dPCR and RT-qPCR based assays can be used for specific and timely detection and monitoring of SARS-CoV-2 variants in wastewater. The slight variations in the proportion of variants detected by each method might be attributable to differences in sensitivity and specificity between qPCR and dPCR.

Data from the qPCR SARS-CoV-2 genotyping feasibility experiments with wastewater samples collected from within the Atlanta metro area demonstrated the utility of the method for the determination of XBB and BQ.1 relative amounts where SARS-CoV-2 N-gene qPCR assay threshold cycles (Ct) ranged from 30 – 37, which is within the typical range for circulating SARS-CoV-2 in wastewater samples (Table 7). A 37 Ct threshold was determined to be the recommended threshold requirement for the genotyping assays. Addition of controls at 0%, 25%, 50%, 75% and 100% were evaluated and it was determined that 0%, 50% and 100% are required for relative variant quantitation.

**Table 7:**
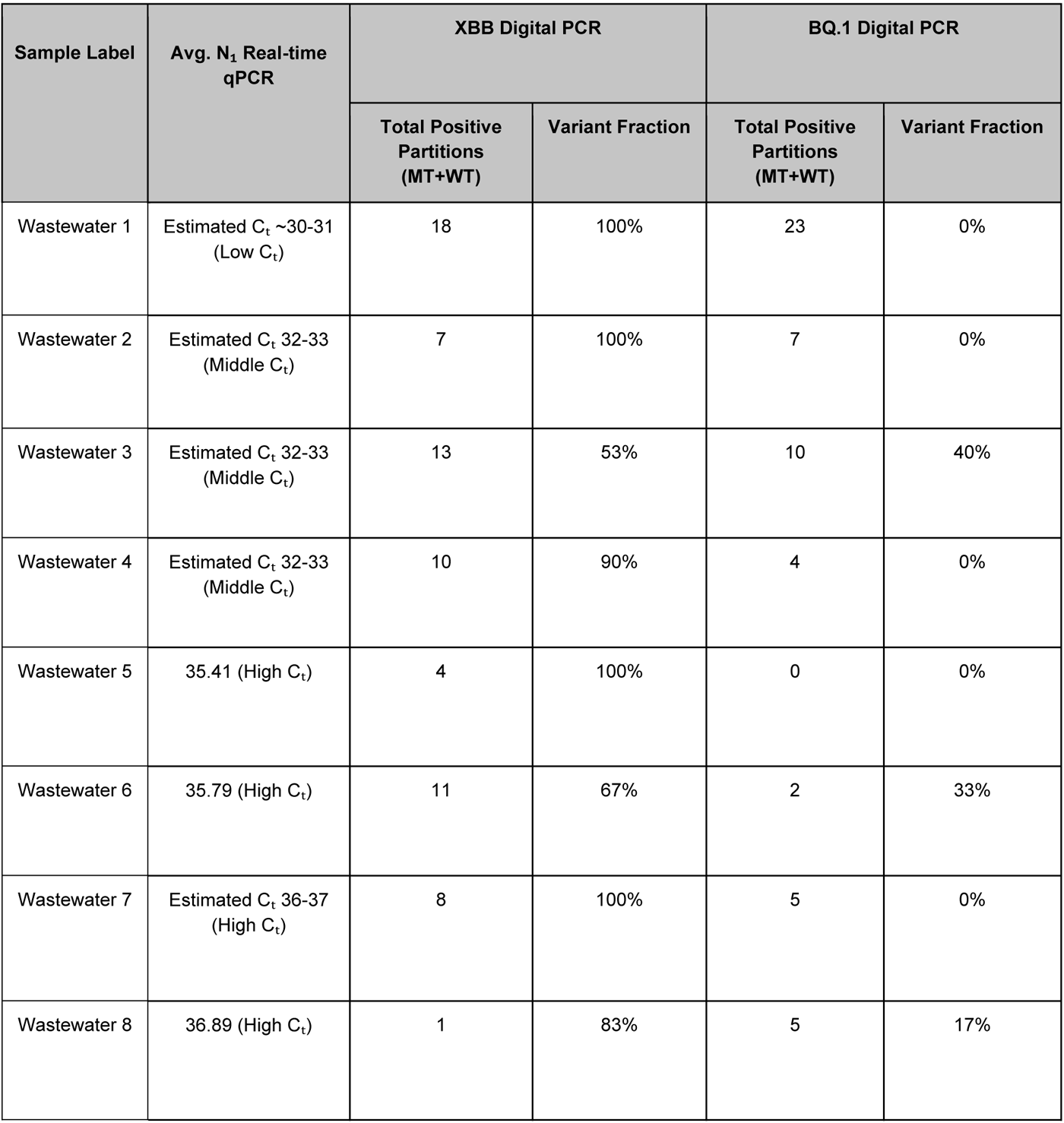
Performance of genotyping assay for Digital PCR at low, middle, and high Ct value. This table presents the average N₁ real-time qPCR estimated Cₜ values alongside the detection results of XBB and BQ.1 variants using digital PCR in eight wastewater samples. The data includes total positive partitions (MT+WT) and prevalence for both variants, highlighting varying detection levels across different Cₜ ranges. XBB variants were consistently detected at varying fractions across all samples, while BQ.1 variants showed much lower or zero detection rates in most samples.

#### C. Assessment of both 96-well and 24-well microwell plate formats on QIACuity dPCR system

The SARS-CoV-2 genotyping assays manufactured by ThermoFisher were validated on QuantStudio qPCR Instruments. Compatibility with Qiagen QIAcuity dPCR instruments was assessed using both 96-well and 24-well nanoplate formats. The 24-well plate format provides 26k partitions per well allowing for increased assay sensitivity, but lower sample throughout and higher cost per sample. Improved sensitivity provided by the 24-well nanoplate format was determined to be the best option for wastewater genotyping where the introduction of new variants can present at low circulating concentration within wastewater samples. Moreover, we determined that a minimum of five positive partitions is required for a reliable determination of variant fraction in the wastewater sample.

### Creating the Wastewater Processing and Genotyping SOP (Revision date April 03, 2023)

Successful implementation of the SARS-CoV-2 genotyping assay in combination with Nanotrap Particle wastewater sample processing was completed at Emory University using samples collected within the Atlanta metro region. The samples utilized for the verification study contained BQ.1, XBB or a combination of the two SARS-CoV-2 variants representing a transition period between BQ.1 and XBB. The Standard Operating Procedure (SOP) was developed based on the criteria established during this phase for both standard qPCR and dPCR methods. The SOP workflow diagram is shown in figure 9. This SOP was implemented for Panel 1 of this study.

**Figure 9:**
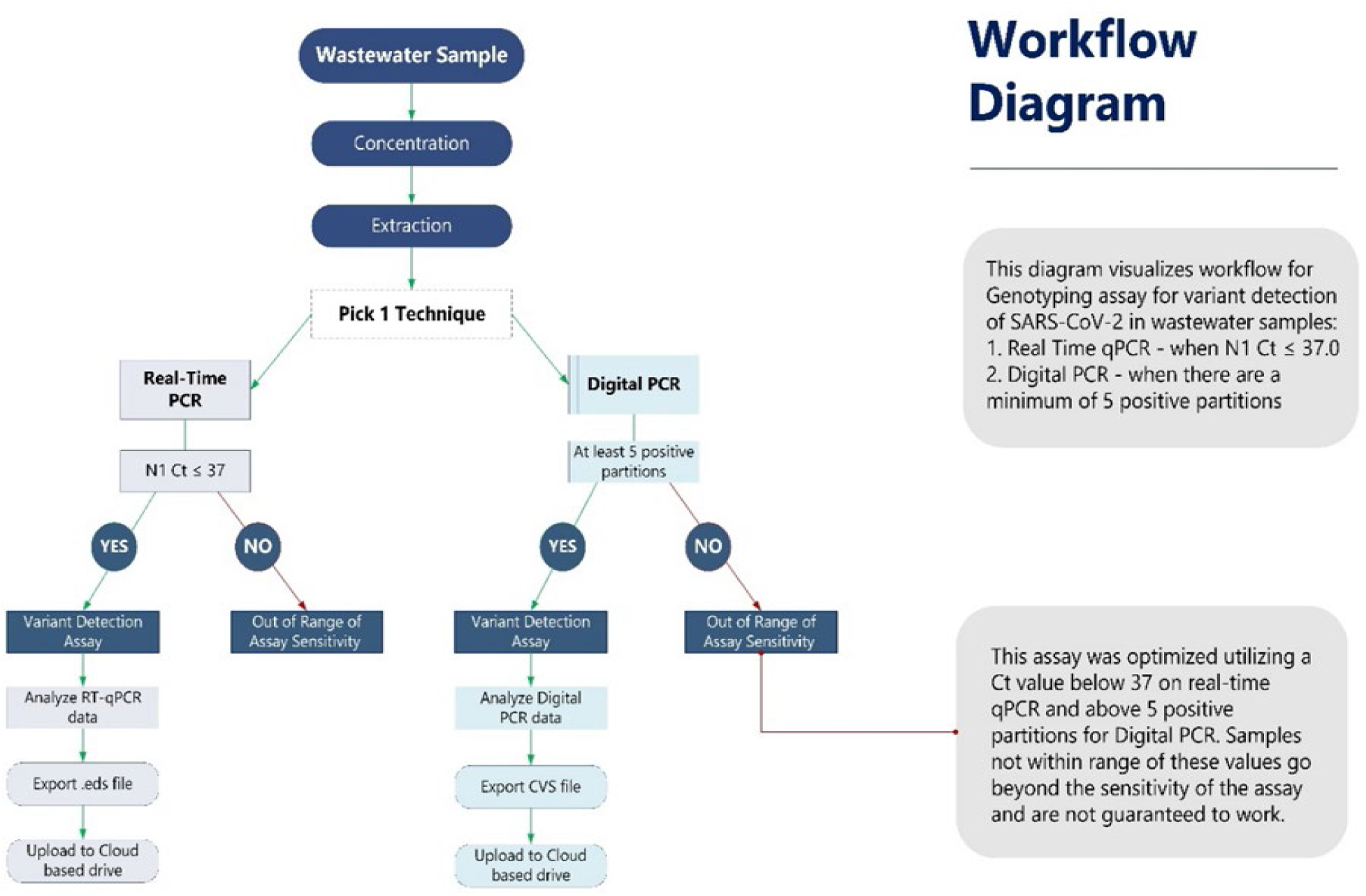
Wastewater Genotyping Project Workflow Diagram. This diagram outlines the genotyping assay process for detecting SARS-CoV-2 variants in wastewater samples, using either Real-Time PCR (N1 Ct ≤ 37) or Digital PCR (≥ 5 positive partitions). Successful assays proceed to variant detection, data analysis, and uploading results to a cloud-based drive. The workflow is optimized for specific Ct values to ensure assay sensitivity.

### Implementation Period 1-Emory University: Georgia State

#### A- Panel 1 (BQ.1 and XBB variants)

Following the SOP Revision 1, dated 04/03/2023, Emory University alpha site ran 86 wastewater samples from multiple sites in the state of Georgia that had been collected between April 1 and June 19. The assays were run on a QIAcuity dPCR system and the data were uploaded to the ROSALIND Tracker. As demonstrated in Figure 10, these data demonstrate that the XBB variant was the most prevalent during that time period and that the BQ variant was the second most common. These data are consistent with the clinical sample genotyping results from that sample time period, though it is important to note that the number of clinical samples from Georgia in that time period is very low – 107 clinical samples were collected and genotyped, representing data from 107 individuals, whereas the 86 wastewater samples represent results from hundreds of thousands of individuals.

**Figure 10:**
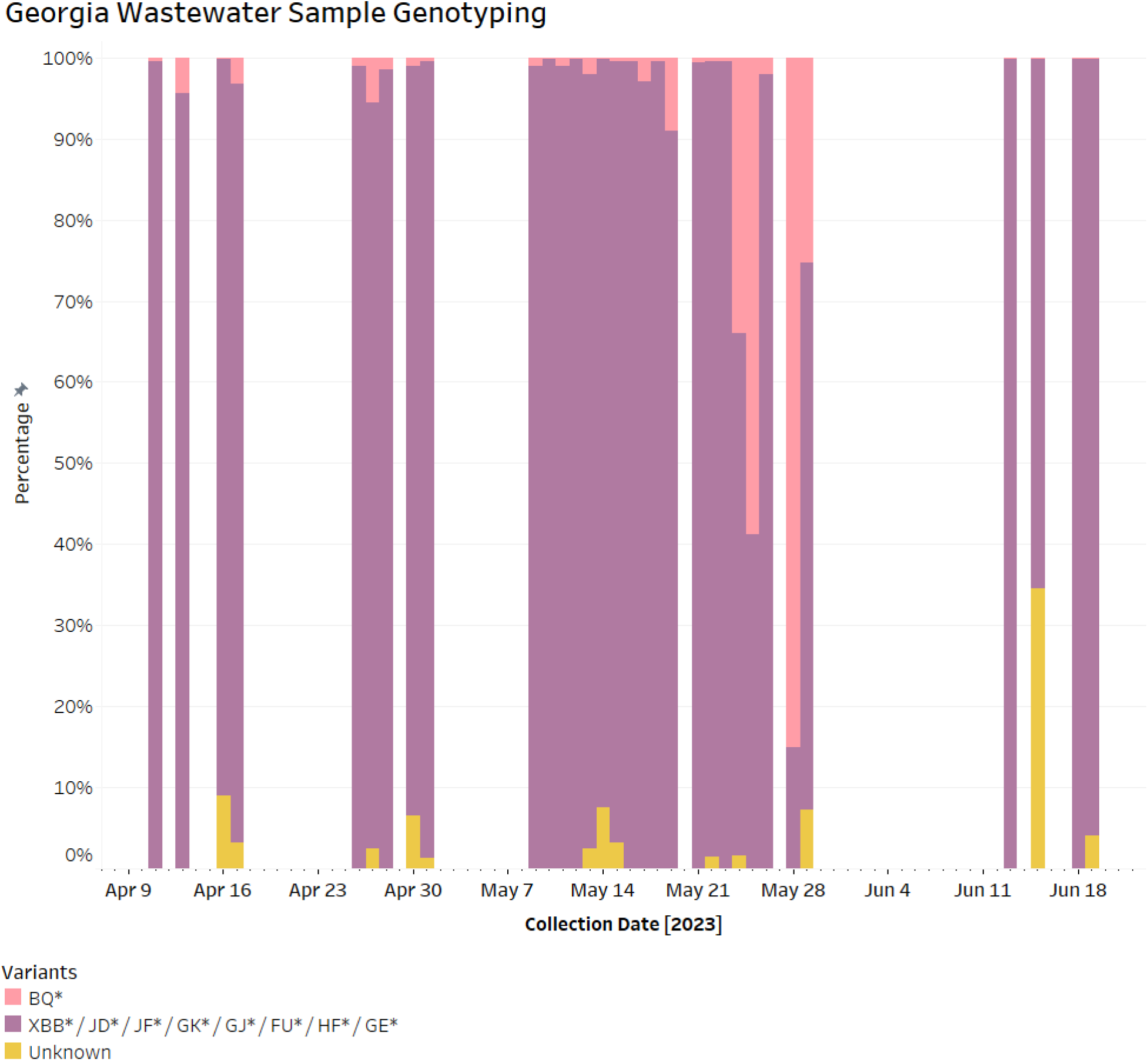
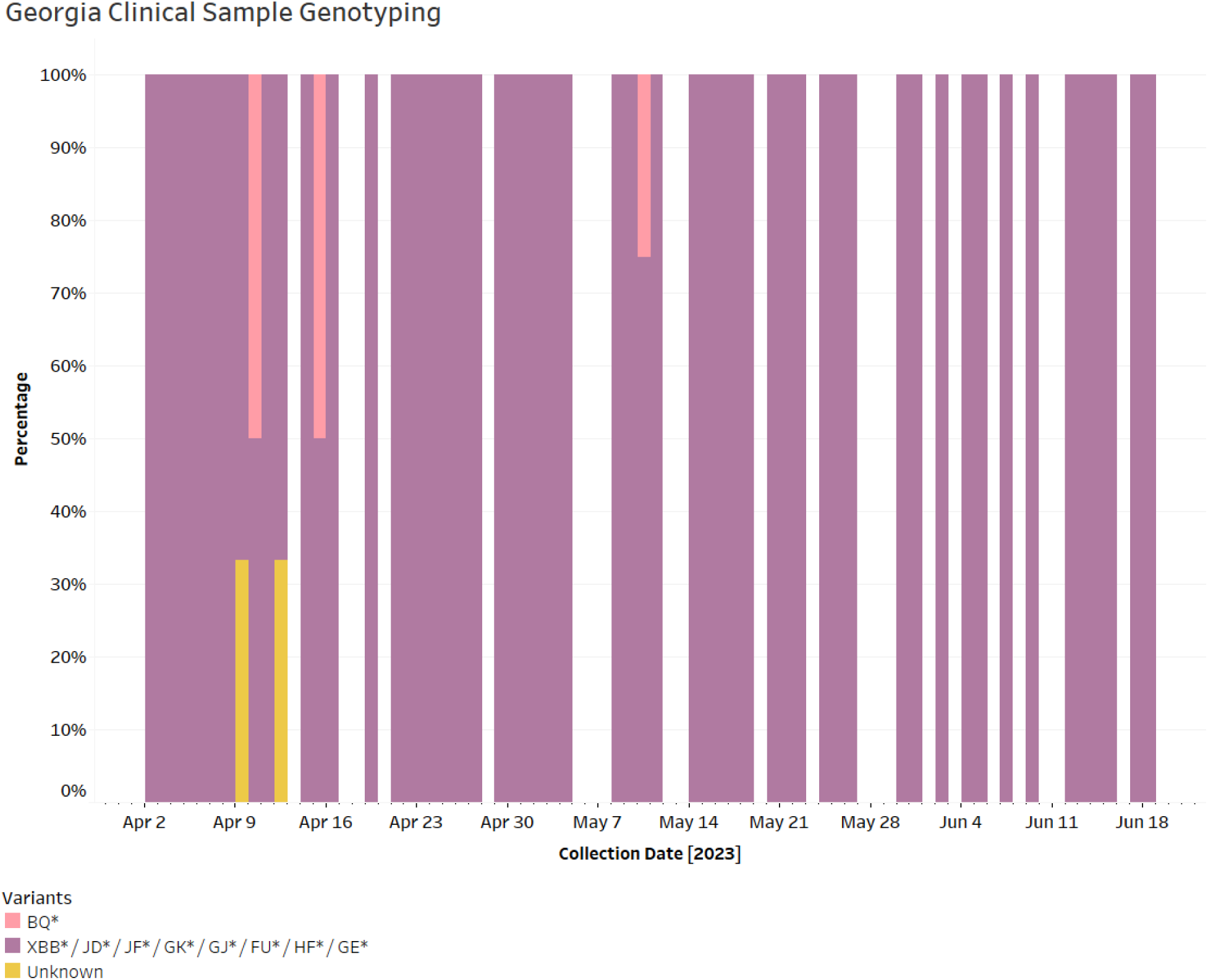
Data from ROSALIND TRACKER, showing variant data from wastewater and clinical samples from the State of Georgia from April 1 to June 19 2023. Despite the fact that the marker sets being used for these two sample types are not exactly the same, the data show that XBB* and BQ* were the predominant variants in Georgia during this time period. Data accessed from https://tracker.rosalind.bio/tracker/dashboard/ on 05/31/2024.

#### B. Panel 2 (XBB, EG.1, EG.5, and FL variants)

At the end of June 2024, the NIH Variant Task Force determined that it was necessary to update the clinical sample and wastewater sample variant genotyping panel to monitor the following variants XBB, EG, FD, and FL. We hoped to use the same assays for wastewater genotyping as were being used for clinical sample genotyping. Because the XBB and BQ assays had worked well for wastewater-based dPCR genotyping, we expected that the new EG, FD, and FL assays would also work. This was true for the EG assay, which provided results for wastewater samples on the dPCR system using the same assay conditions as XBB and BQ. Unfortunately, the FD and FL assays did not work on the dPCR system for wastewater samples under these conditions.

We noticed two problems with the FD assay. It was experiencing double amplification on the dPCR and the wild type control for FD was amplifying. ROSALIND reported that the clinical labs testing the FD assay were also experiencing similar challenges with it. Ultimately, this assay was discarded and not implemented at the panel level.

The FL assay, when run under the dPCR conditions that were optimized for XBB and EG, had very little separation between the negative and positive partitions. Emory, ROSALIND, and Ceres evaluated multiple solutions to this, including longer probe lengths, manual threshold setting, and altering the dPCR assay parameters. The longer probes did not resolve the issue, and manual threshold setting on a plate-by-plate basis was deemed too onerous as a resolution, so we ultimately determined that the best approach assay was to alter the dPCR conditions. The key changes that were made were lengthening the Denaturation and Annealing / Extension times and lowering the Annealing / Extension Temperature. See Figure 11 for an example of how changing the assay conditions improved the results on the dPCR system for this assay.

**Figure 11:**
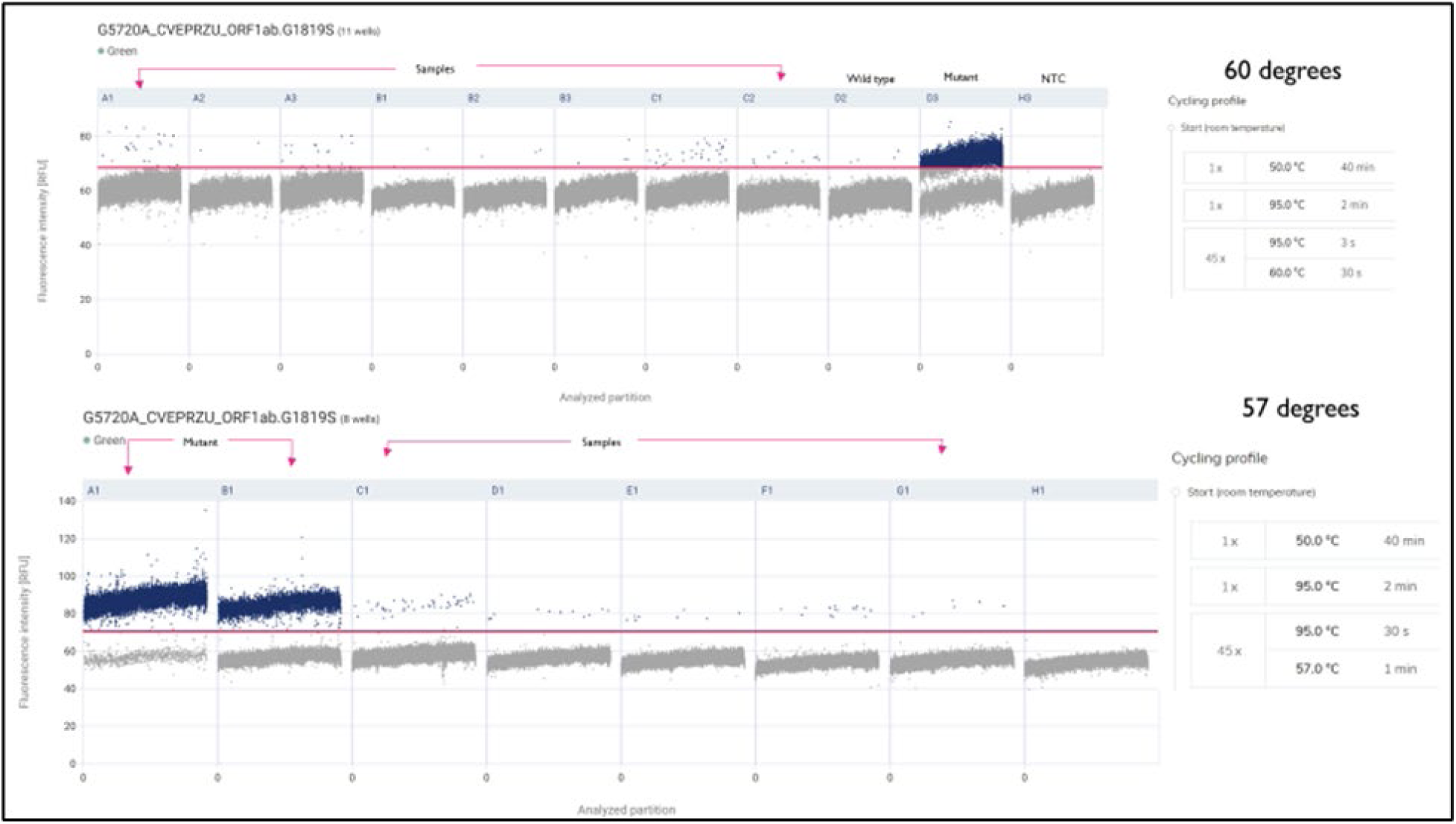
FL assay performance on the QIAcuity dPCR system was dramatically improved when the denaturation and annealing/extension conditions were slightly altered. The top frame is the PCR thermocycling conditions used for XBB and EG assays. The bottom frame shows updated conditions that were selected for the FL assay. Partitioning improved by increasing the denaturation & annealing/extension time and decreasing the annealing/extension temperature.

By the end of September, we had successfully verified that XBB, EG1, and FL (with modified dPCR conditions for FL) could be used in a panel for wastewater samples.

In mid-September 2024, the NIH Variant Task Force again decided to update the variant genotyping panel to monitor the following variants XBB, EG.1, EG.5, and FL. We started by running a validation process for the 0%, 50%, and 100% control samples across all four of these assays using the qPCR QuantStudio platform. These assays performed well in this context. In October, we then ran roughly 60 archived wastewater samples collected from sites in Georgia between mid-June and early September using these assays. The results confirmed that these assays worked for wastewater samples. As demonstrated in Figure 11, these data demonstrate that the XBB variant was the most prevalent in Georgia during that time period and the results were still consistent between the wastewater samples and the clinical samples.

We also noted that despite the fact that there were 3 times fewer wastewater samples than clinical samples tested in Georgia during this time period, that the FL and EG.5 variants were detected 22 days and 31 days earlier in the wastewater samples than in the clinical samples (Figure 12).

**Figure 12:**
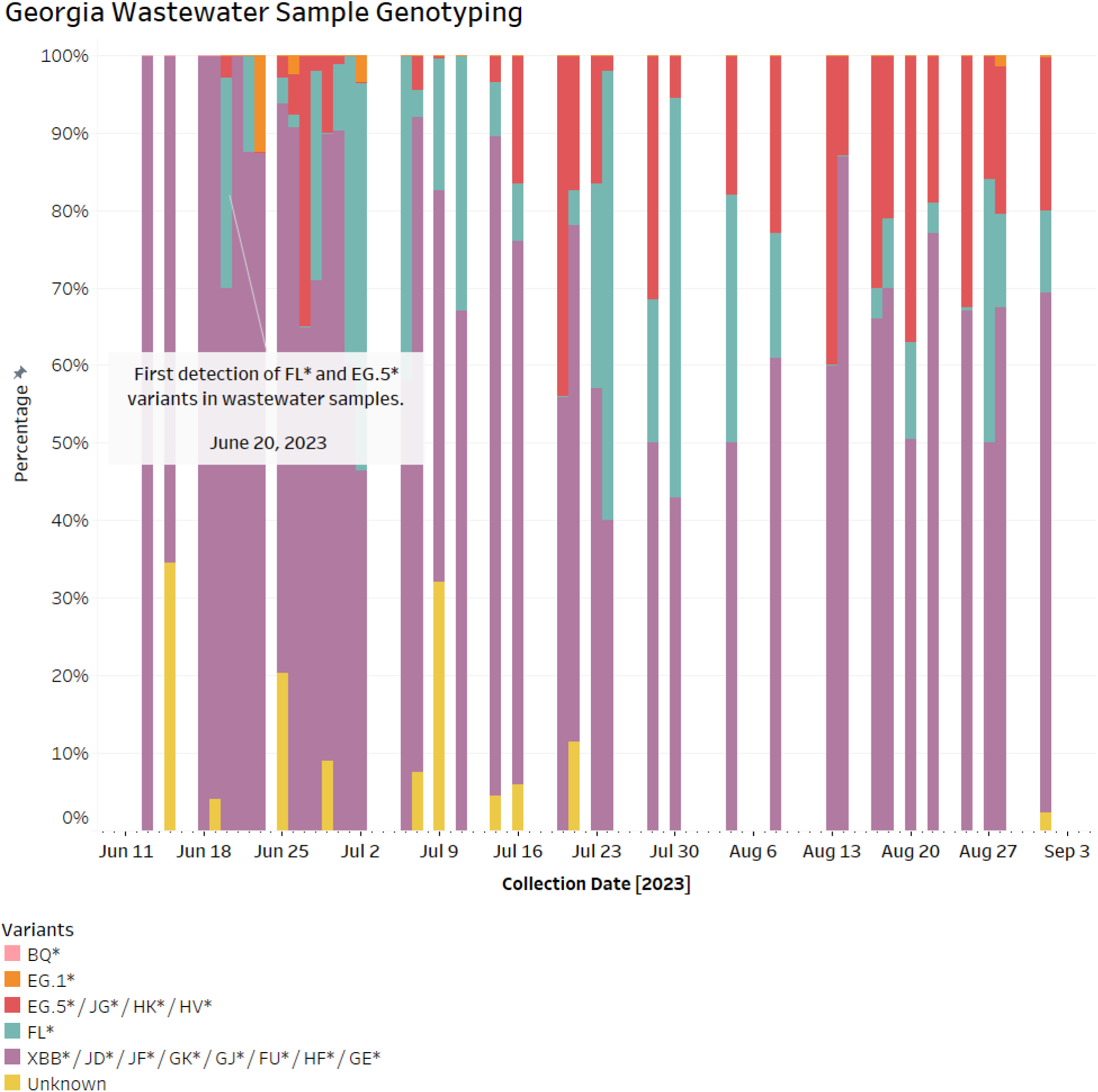
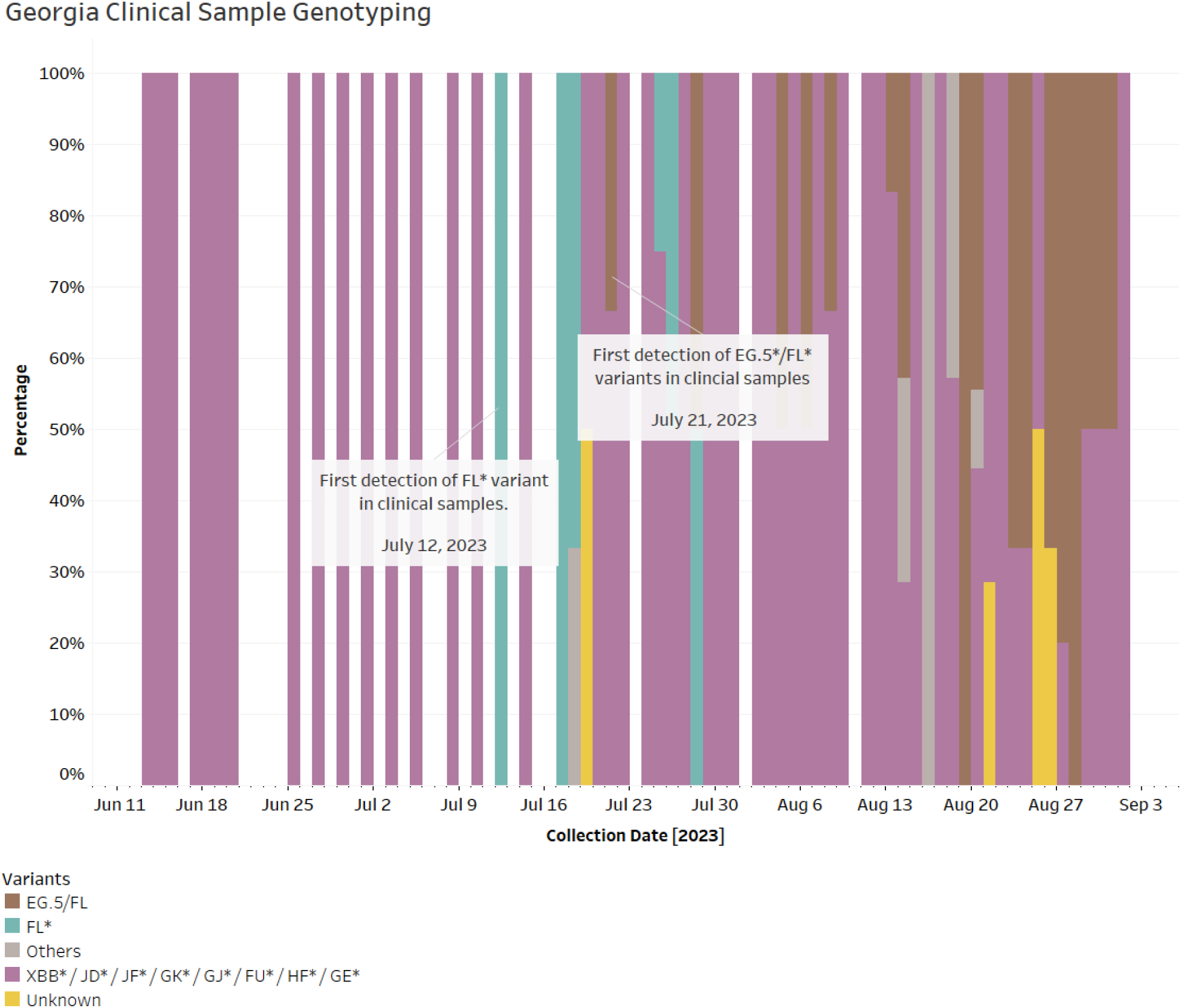
Data from ROSALIND TRACKER, showing variant data from wastewater and clinical samples from the State of Georgia from June to September 2023. June 20, 2023 first detection of FL and EG.5 variants in wastewater samples. July 12, 2023 first detection of FL variant in clinical samples. July 21, 2023 first detection of EG.5/FL variant in clinical samples.

At this point, the SOP was updated to include the details on the new variant panel and assay settings to be used in the expanded pilot phase of this project. This new version was named “Variant detection SOP-Rev.1-1” and is dated 11/14/2023. The SOP is available on Ceres Nanosciences’ website at the following URL: https://www.ceresnano.com/post/study-protocol-for-sars-cov-2-variant-detection-in-wastewater.

### Implementation Period 2-Winter 2023-2024: Expansion to five pilot labs with the XBB, EG.1, EG.5, and FL panel

In addition to Emory University, which continued wastewater genotyping throughout the fall and winter of 2023-2024, four more wastewater testing laboratories began dPCR-based wastewater genotyping for a panel of XBB, EG.1, EG.5, and FL at a cadence of about 17 samples per week: GT Molecular and Wisconsin State Laboratory of Hygiene started in November 2023, the University of Illinois Chicago started in December 2023, and the State University of New York at Buffalo started in January 2024. These laboratories expanded the wastewater genotyping project to cover six states: Georgia, California, Illinois, Wisconsin, New York, and Louisiana.

### Third wastewater panel - XBB, EG.5, FL, JN

Shortly after the five wastewater testing laboratories began the wastewater genotyping using the assays for Panel 2 (XBB, EG.1, EG.5, and FL), a new variant started to rapidly rise in prevalence in the United States – JN. During early 2024, we evaluated two assays for the markers ANDKJKV and ANCFPZX. The ANDKJKV assay, under the dPCR conditions used for XBB, EG.1, and EG.5), exhibited double bands in both WT and MUT channels and false positive partitions (<5) in the WT channel for CTRL-100 (mutant homogeneous control). We evaluated longer PCR assay conditions, which resolved the false positive issue (Figure 13).

**Figure 13:**
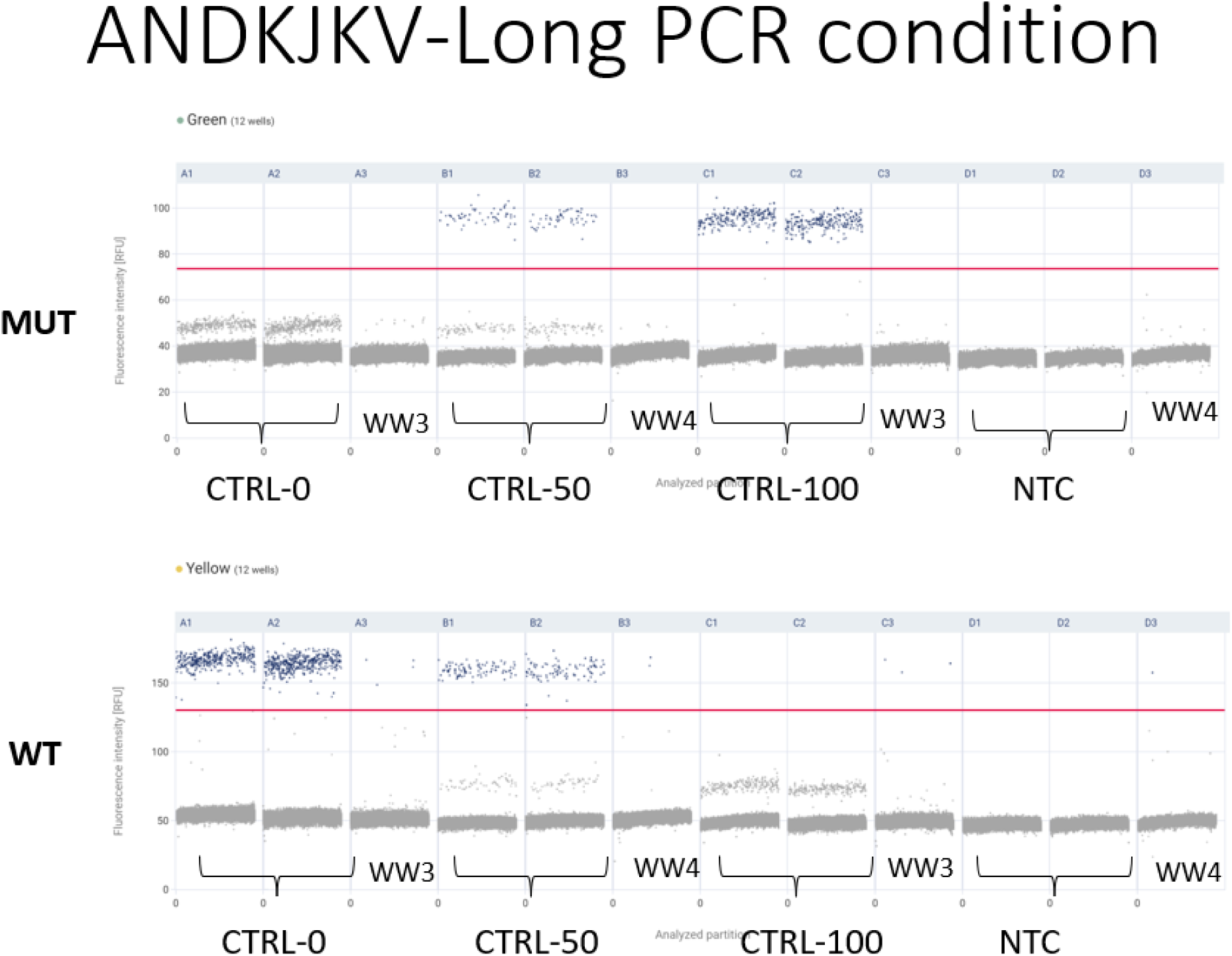
The performance of the JN variant genotyping assay ANDKJKV improved under the longer PCR conditions established for the FL assay in Panel 2. The top graph shows the Mutation Channel, and the bottom graph displays the Wild Type Channel. Double bands were observed in both channels, which can be accurately distinguished by proper thresholding to identify true positive partitions. Controls and samples performed as expected under these conditions. CTRL-0 represents 0% mutation, CTRL-50 represents 50% mutation, and CTRL-100 represents 100% mutation. NTC indicates the No Template Control, and WW represents wastewater samples.

Similarly, the ANCFPZX assay showed double bands in WT and MUT channels, but the right threshold could distinguish between false and true positive partitions. Sporadic false positives (<5 partitions) were observed in CTRL-0 (wild type homozygous control) in the MUT channel. Longer PCR conditions brought the double bands closer together, complicating thresholding. Considering performance and higher result quantity, as well as shorter run time, ANCFPZX was selected for JN variant genotyping (Figure 14).

**Figure 14:**
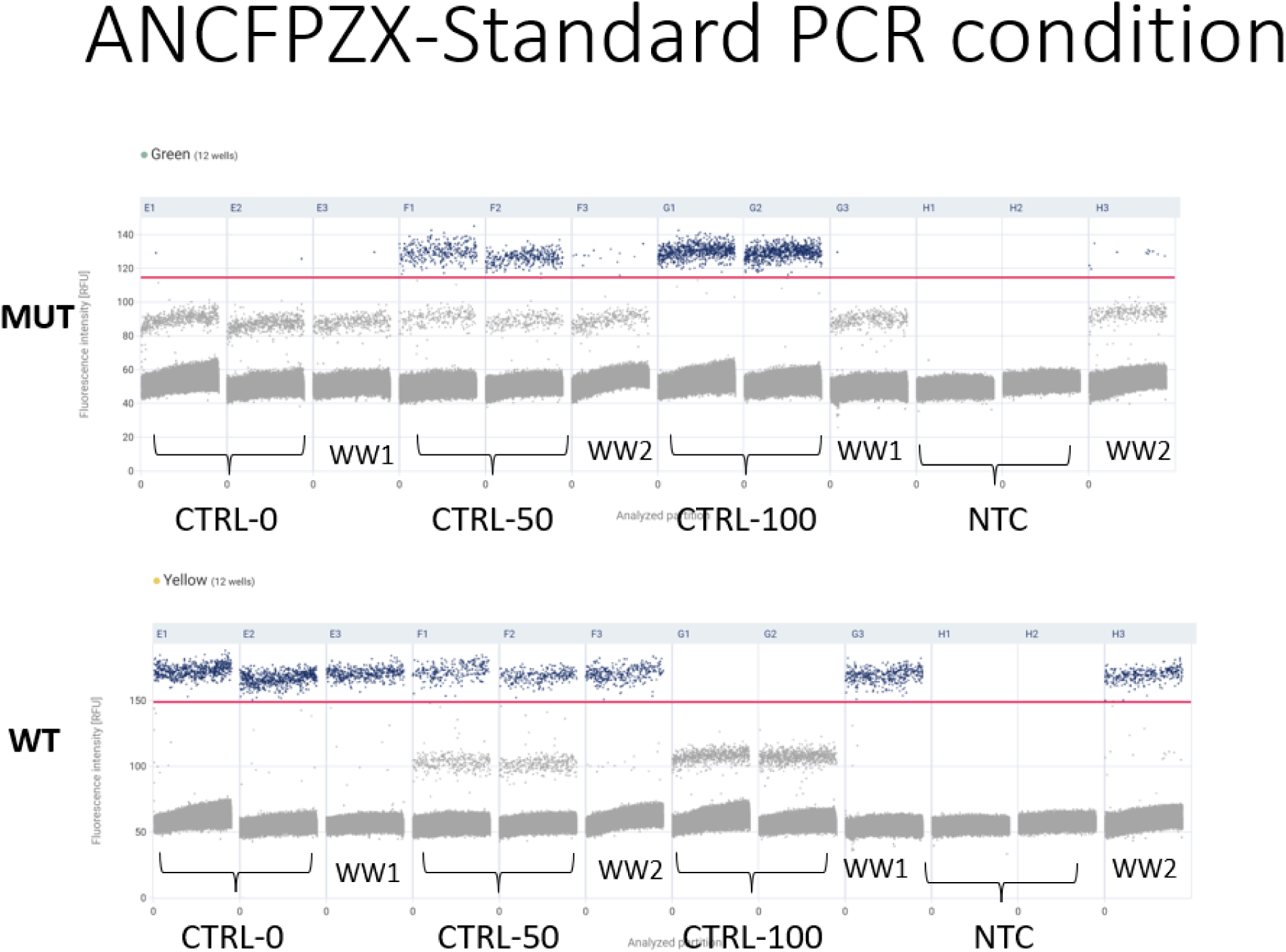
The performance of the JN variant genotyping assay ANCFPZX under the standard PCR conditions established in Panel 2. The top graph shows the Mutation Channel, and the bottom graph displays the Wild Type Channel. Double bands were observed in both channels, which can be accurately distinguished by proper thresholding to identify true positive partitions. Fewer than five false positive partitions were observed in CTRL-0 in the Mutation Channel. Other controls and samples performed as expected in both channels. CTRL-0 represents 0% mutation, CTRL-50 represents 50% mutation, and CTRL-100 represents 100% mutation. NTC indicates the No Template Control, and WW represents wastewater samples.

Validation of JN assay was completed by the end of January 2024. At this point, the SOP was updated to retire EG.1 assay and include the details on the new JN variant assay. This new version was named “Variant detection SOP-Rev.2” and is dated 02/02/2024. Reagent and supplies required for the new panel 3 were shipped to all five labs and instructed to continue wastewater testing using the new panel. Because JN had risen in prevalence more quickly than we could develop and validate the assay, we missed the rise of JN1 in real time (the ROSALIND Tracker dashboard showed a lot of unknown variants in January and February). So, we asked the labs to go back and retest any leftover RNA from January and February using the JN assay. These data were uploaded to the ROSALIND Tracker and were used to retrospectively update the dashboard.

## Discussion

For multiple states, we compare the surveillance data on wastewater genotyping, clinical genotyping, and GISAID clinical sequencing presented on the ROSALIND tracker. It is worth noting that, in contrast to clinical samples, which usually have one dominant variant (e.g., 100% XBB), the genotyping and sequencing results from wastewater typically present a profile of multiple variants (e.g., 70% JN, 20% XBB, 10% EG5) (41). This is because wastewater samples are composites with contributions from multiple individuals, capturing a broader range of circulating variants. Therefore, the variant prevalence in all wastewater samples is aggregated bi-weekly, and the percentage of the total is calculated. In contrast, only individuals with symptoms visit clinics for testing, so the variant prevalence in clinical samples is based on the number of patients observed with the variant of interest during the bi-weekly period, relative to the total number of patients.

We compared the wastewater genotyping data to clinical genotyping data, which was built on the same genotyping assay using the qPCR platform. In response to the emergence of Omicron, a genotyping panel was developed to distinguish Delta and Omicron using four highly specific SNPs from patient samples. The results demonstrate the utility of this condensed panel to rapidly track the growing prevalence of Omicron across the U.S. in December 2021 and January 2022, as previously described (25). As of July 2, 2024, the clinical genotyping dashboard contains the results of 195,549 total samples in the United States (32).

Since January 2020, genomic sequences shared via GISAID, the global data science initiative, have been the primary source of genomic and associated data from SARS-CoV-2 cases (31,42). As of July 2, 2024, more than 16.8 million SARS-CoV-2 sequences have been uploaded to the GISAID EpiCoV database globally, with over 5 million from the United States (32,43). This high-quality, curated data has enabled the rapid development of diagnostic and prophylactic measures against SARS-CoV-2, including the first diagnostic tests and vaccines to combat COVID-19, as well as continuous monitoring of emerging variants in near real-time (31). In this study, we compared the wastewater genotyping data to sequencing data associated with 214,855 sequences available on GISAID from January 1, 2023, to May 20, 2024, for the states of California, Georgia, Illinois, Louisiana, New York, and Wisconsin. Sequencing results were aggregated to match the variant classification in the wastewater genotyping panels, as indicated in tables 3, 4, and 5.

### Georgia State

#### a. Georgia Wastewater Genotyping

Data from wastewater samples in Georgia was collected in January 2023 as part of the feasibility study for the wastewater genotyping project. Figure 15 presents genotyping results of wastewater samples in Georgia from April 11, 2023 to April 5, 2024 (52 weeks), highlighting the percentage of total markers in samples for different groups over a two-week period and the number of samples collected. Initially, the XBB* (purple line) marker dominated with nearly 100% presence in April 2023 but declined steadily to near 0% by December 2023. In contrast, the BA.2.86*/JN* (blue line) marker dramatically increased starting in January 2024, becoming the dominant variant by early 2024. The EG.5* (red line) marker shows fluctuations with a significant presence starting in July 2023, peaking at 50% in November 2023, and gradually declining. The BQ* (pink line) marker exhibits a sharp spike and decline in May 2023, disappearing by June. The FL* (cyan line) marker fluctuated throughout the year, peaking in August 2023, and maintained a consistent low presence until the end of the period. The EG1* (orange line) marker had a brief spike around December 2023 but otherwise maintained a low presence. The black dotted line represents the number of samples collected, averaging 20 samples bi-weekly and remaining steady for a total of 528 wastewater samples during this period.

**Figure 15:**
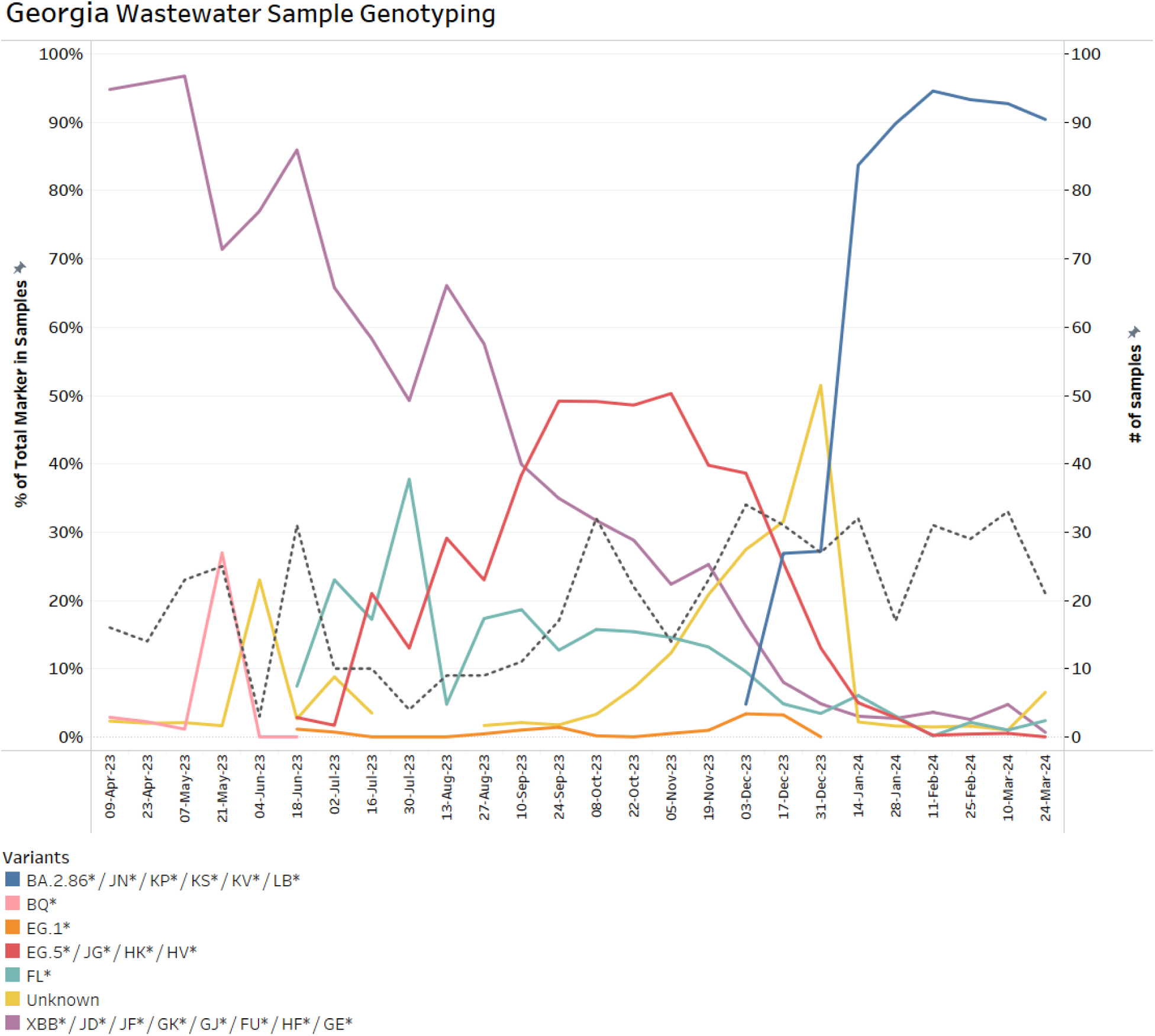
Temporal Distribution of SARS-CoV-2 Genotypes in Georgia Wastewater Samples (April 2023 - April 2024). The graph shows the prevalence of various SARS-CoV-2 genotypes over time. Initially, XBB* (purple) dominated but declined by December 2023. BA.2.86* / JN* (blue) surged to 90% around February 2024. EG.5* (red) peaked at 50% in October 2023. FL* (cyan) showed sporadic peaks, while BQ* (pink) briefly peaked in May 2023. The Unknown marker (yellow) demonstrates an upward trend from October 2023 to January 2024, indicating the introduction of a new variant in the wastewater. The black dotted line represents the number of samples collected, staying steady around 20 samples per two weeks. The data indicate shifts in viral genotypes, suggesting changes in infection patterns or new variant introductions.

The Unknown (yellow line) marker shows periodic spikes, particularly in June and November 2023. Interestingly, the upward trend from October 2023, rising to about 50% in January 2024, experienced a sharp decline after transitioning to the new panel with the JN assay in February 2024. This likely indicates that the unknown marker was indeed the JN variant, and its rising prevalence was a precursor to the later dominance of the BA.2.86*/JN* variant. Retrospective analysis of a small set of samples collected between December 2023 and January 2024 confirmed the Unknown variant as JN variants (Figure 16). This underscores the importance of continuous assay development and implementation in accurately identifying and tracking emerging variants. We suggest studying the trend and setting a threshold of Unknown variant detection in wastewater samples to trigger new assay design and required validation works. This ability to rapidly identify new variants is crucial for timely public health responses and interventions. The emergence and identification of the JN variant highlight the dynamic nature of SARS-CoV-2 evolution and the ongoing need for adaptive surveillance strategies.

**Figure 16:**
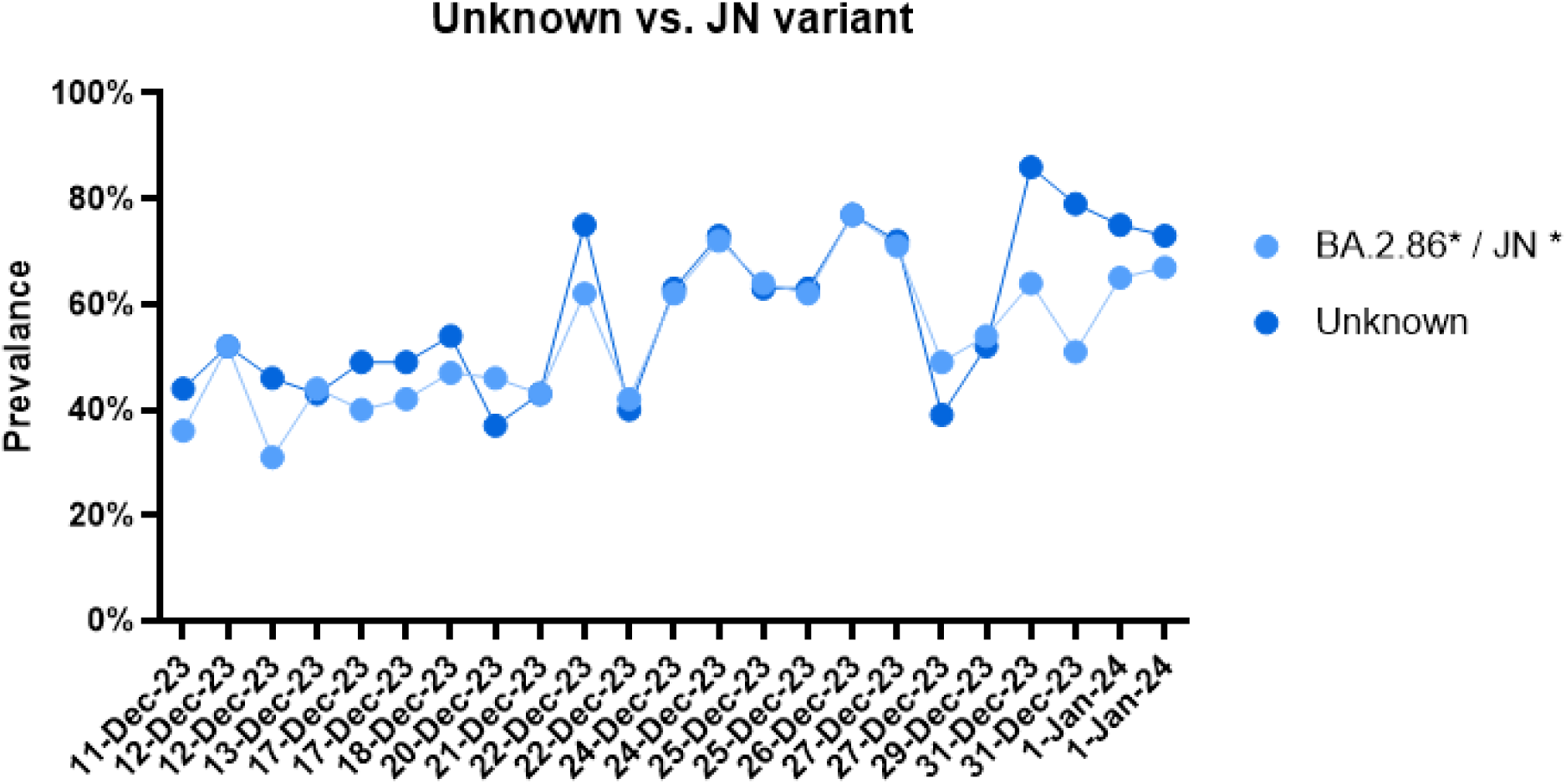
Comparison of Unknown and JN Variant Prevalence in Wastewater Samples (December 11, 2023 - January 1, 2024). The high correlation between the Unknown variant and the BA.2.86*/JN* variant suggests that the Unknown marker from the original Panel 2 analysis is likely the BA.2.86*/JN* variant. Both variants show similar trends and fluctuations, with prevalence ranging from 40% to 80% and notable spikes in mid to late December 2023.

The graph depicting the genotyping results from wastewater samples in Georgia from April 2023 to April 2024 reveals three distinct phases of SARS-CoV-2 variant dynamics. The initial phase, from April to August 2023, is marked by the high prevalence of the XBB* variant, which starts near 100% and gradually declines, indicating a transition away from its dominance. The second phase, spanning August 2023 to December 2024, highlights the emergence and fluctuation of several new variants, including EG.5* and FL*. During this period, the unknown marker, later identified as the JN variant, begins to rise in prevalence from October 2023, underscoring the importance of adaptive assay development for accurate variant identification. The final phase, from December to April 2024, is characterized by the rapid rise and dominance of the BA.2.86*/JN* variant, which becomes the predominant variant by February 2024. This period also marks the replacement of the previously unknown marker by the identified JN variant, reflecting its significant circulation since October 2023. These observations underscore the dynamic nature of SARS-CoV-2 variants and the critical need for continuous monitoring and adaptive public health strategies to manage the evolving pandemic landscape.

The Unknown/BA.2.86*/JN* group’s rapid rise starting in October 2023 suggests a significant event, such as an outbreak or increased transmissibility, making it the dominant variant by February 2024. The BQ* group’s sharp spike in May 2023 and disappearance by June suggest a brief, unsustained outbreak. The EG.5* group’s fluctuations and peak in November 2023 indicate periodic surges. While the FL* group’s fluctuations suggest localized outbreaks or transient competitive advantages, the consistently low prevalence of EG.1 indicates it did not achieve widespread transmission.

#### I. JN vs. Unknown variant

Figure 16 compares the prevalence of the BA.2.86*/JN* variant and the Unknown variant in 23 wastewater samples collected between December 11, 2023, and January 1, 2024. Initially analyzed using Panel 2 (including EG.5, EG.1, FL, XBB), these samples were later re-analyzed with a JN-specific assay to determine the prevalence of the JN variant. The graph reveals a high degree of correlation between the Unknown variant detected in the original Panel 2 results and the BA.2.86*/JN* variant identified using the JN-specific assay, suggesting that the Unknown variant is likely the BA.2.86*/JN* variant. Both variants show similar trends and fluctuations, with prevalence ranging from 40% to 80% and notable spikes around December 17, December 23, and December 28, 2023. This consistency confirms that the Unknown marker detected in the initial analysis corresponds to the BA.2.86*/JN* variant.

#### b. Georgia Clinical Genotyping

The graph in Figure 17 presents genotyping results of SARS-CoV-2 from clinical samples collected from patients who visited clinics in Georgia from April 9, 2023 to February 8, 2024. Initially, the XBB* (purple line) variant dominated with nearly 100% prevalence in April 2023, but it showed a steady decline starting in June 2023, fluctuating at 20% by February 2024. In contrast, the BA.2.86*/JN*-Unknown (blue and Yellow line) variants dramatically increased starting at the end of October 2024, becoming the dominant variant by early 2024. As we discussed in the previous section, the upward trend of the Unknown marker from October 2023 to January 2024, indicating the introduction of a new variant. The Unknown variant was identified as the JN variant after assay introduction

**Figure 17:**
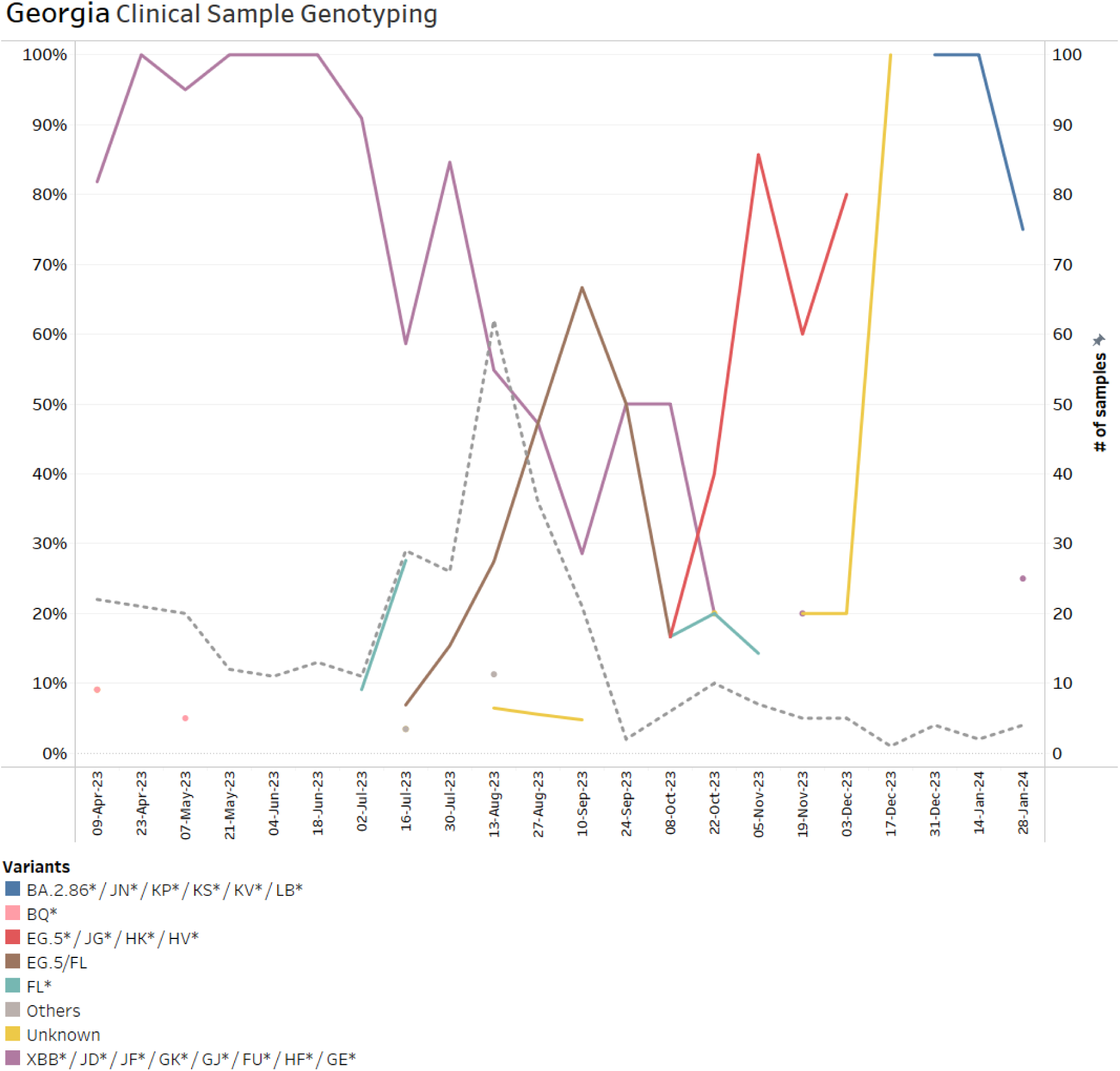
Temporal Distribution of SARS-CoV-2 Genotypes in Georgia Clinical Samples (April 2023 - February 2024). The graph presents the genotyping results of clinical samples in Georgia from April 2023 to February 2024, showing the prevalence of various SARS-CoV-2 genotypes and the number of samples collected. Initially, the XBB* (purple) genotype dominated, nearly 100% in April 2023, but declined steadily to around 50% by October 2023 and further dropped towards the end of the period. The BQ* (pink) genotype had very low presence, briefly peaking in April-May 2023 before disappearing. The EG.5* and FL* (red, cyan, brown) variants appeared in July 2023 and dominated the patient samples from September to December 2023, prior to sudden replacement with an Unknown/ JN* variants. The dotted black line represents the number of samples which experienced a sharp decline at the end of August. No clinical samples have been tested after February 8, 2024.

A small fraction of the patient population (<10%) had the BQ* (pink line) variant in April 2023 which disappears in May 2023. Two new variants (EG.5 and FL) were detected in July 2023. The EG.5+/JG+/HK2+/HV* (red line) variant peaking at about 86% of patients in November 2023 before gradually declining. The FL* (cyan line) variant fluctuated between July to November, but generally maintained 20% of patients. The clinical genotyping dashboard also had a panel for both EG5 and FL which is indicated by Brown color. Considering both EG5 and FL (Red, Cyan, Brown) markers, these two variants dominated the clinical genotyping from September 2023 to December 2023, with a sudden replacement with the Unknown (yellow) variant.

The average number of patient samples collected during this period (black dotted line) was 13 per bi-weekly period, totaling 330 samples. The number of patient samples for genotyping testing sharply dropped from 62 samples in the first half of August 2023 to 2 samples at the end of September 2023. Eventually, no clinical samples were tested after February 8, 2024. Many factors might impact the number of clinical samples, including changes in population immunity, virus transmission rate, healthcare seeking behavior, and policy changes.

It is also worth noting that the low number of clinical samples caused gaps and unsmoothed line graphs, making interpretation challenging, particularly from September 2023 to the end of the study period. However, the clinical sequencing data from GISAID can be used to validate the results of the clinical genotyping dataset since genomic surveillance continued after the expiration of the COVID-19 public emergency declaration to help identify and monitor SARS-CoV-2 variants. (44)

The clinical genotyping data demonstrates similar phases as the wastewater genotyping data. The first phase, from April to August 2023, is marked by the initial dominance of the XBB* variant with nearly 100% prevalence, and the brief emergence of the BQ* variant, which spikes in April but disappears in May. The second phase, spanning August to December 2023, sees the rise and fluctuation of several new variants, most notably the EG.5+/JG+/HK2+/HV* variant, which peaks in November 2023, and the FL* variant, which shows periodic increases. During this period, the Unknown variant begins to display significant spikes, particularly in October. The third phase, from December to April 2024, is characterized by the dramatic rise of the BA.2.86*/JN* variant, becoming the dominant strain by February 2024. Concurrently, the Unknown variant is identified as the JN variant following the introduction of a specific assay, leading to a sharp increase and peak in February 2024.

This genotyping data suggests dynamic shifts in viral genotypes, reflecting changes in infection patterns or the introduction of new variants in the population over the study period.

#### c. Georgia GISAID Sequencing

Figure 18 presents sequencing results of SARS-CoV-2 variants in clinical samples from Georgia, accessed from the GISAID database. The data covers the period from April 9, 2023 to April 5, 2024, presented in two-week intervals. It shows the percentage of total samples for different variant groups over time and the number of samples collected. Initially, the XBB* variant (purple line) shows high prevalence, starting near 100% in April 2023, but steadily declines to around 2% by February 2024. The BA.2.86*/JN* variant (blue line) dramatically rises starting in November 2023, becoming dominant by early 2024 and peaking at nearly 100% by April 2024. The EG.5+/JG+/HK2+/HV* variant (red line) fluctuates, peaking in October 2023 before declining. The BQ* variant (pink line) and EG1* (Orange line) exhibit a low fraction of the clinical samples (<3%) between April to July 2023. The FL* variant (cyan line) maintains low and fluctuating presence throughout the period. The Others category (yellow line) exhibits periodic spikes, before declining towards the end of the period. In this period, 2855 samples were analyzed with an average of 110 samples per bi-weekly interval. The number of samples collected (black dotted line) peaks around August 2023 and January 2024, indicating increased sampling activity, but significantly declines at the end of the study period.

**Figure 18:**
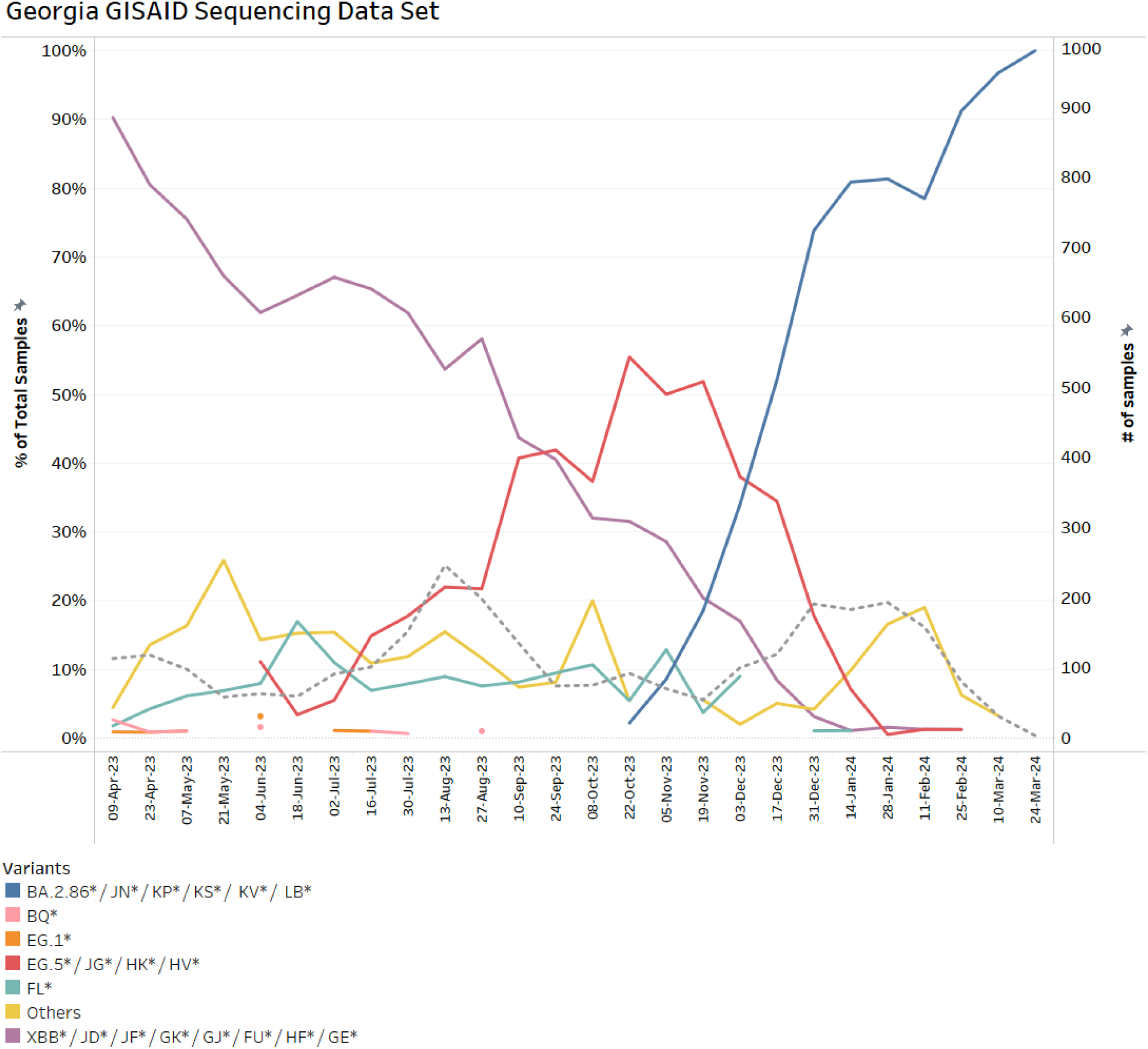
Temporal Distribution of SARS-CoV-2 Genotypes in Georgia Clinical Samples from GISAID Database (April 2023 - April 2024). The graph illustrates the genotyping results of SARS-CoV-2 from clinical samples in Georgia, as recorded in the GISAID database, spanning from April 2023 to April 2024. It shows the prevalence of various SARS-CoV-2 genotypes over time and the number of samples collected. XBB (purple): Initially dominant, comprising nearly 100% of samples in April 2023, but declined steadily to 0% by January 2024. BQ and EG1 (pink, orange): Had very low presence in April and July 2023 before disappearing entirely. FL (cyan): Showed consistently low prevalence from April to December 2023 and disappeared in January 2024. EG.5 (red): Emerged in June 2023, dominated patient samples in October and November 2023, and was suddenly replaced by JN variants. The dotted black line represents the number of samples, averaging 110 samples in bi-weekly increments. The number of samples significantly reduced in February 2024.

The initial dominance and subsequent decline of the XBB* variant, alongside the rapid rise and dominance of the BA.2.86*/JN* variant starting in November 2023, suggest significant shifts in the variant landscape due to factors like increased transmissibility or immune escape. The fluctuations of the EG.5+/JG+/HK2+/HV* variant and periodic spikes in the Others category indicate ongoing viral evolution and the emergence of various other variants. The brief presence of the BQ* variant underscores the potential for transient outbreaks. Increased sampling activity, particularly around August 2023 and January 2024, likely reflects heightened surveillance efforts in response to emerging variants, underscoring the importance of agile and responsive public health measures.

#### D. Georgia Discussion

The data in Figure 19 indicates a significant decline in XBB variants across multiple monitoring sources, reflecting a reduction of these variants within the population. The close correlation between clinical and wastewater data sets suggests that local clinical findings are well-aligned with wastewater surveillance, which captures viral signals from a broader population base, including asymptomatic individuals.

**Figure 19:**
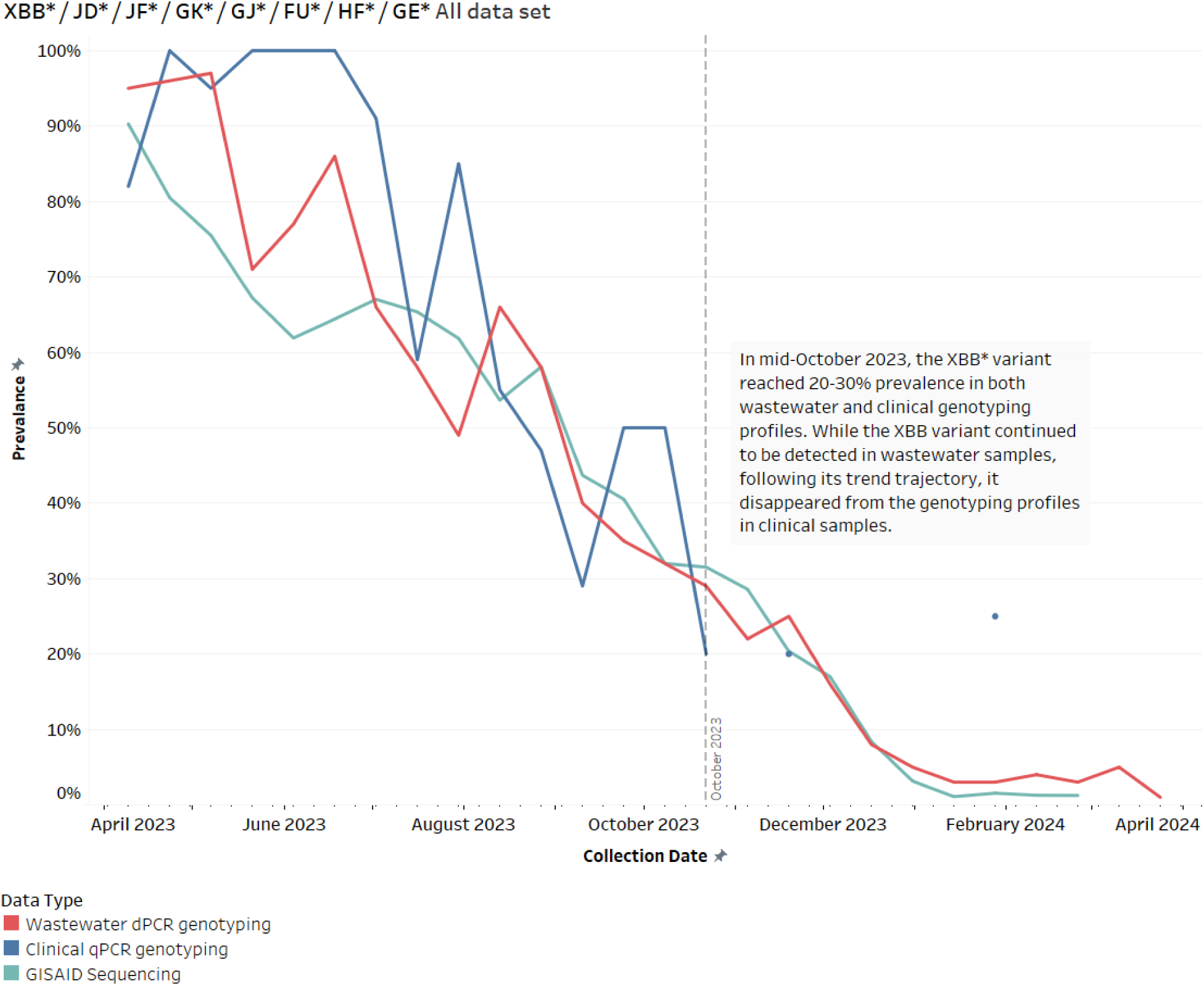
Prevalence of XBB* Variant Across Clinical, GISAID, and Wastewater Data Sets in Georgia (April 2023 - April 2024). The graph shows the prevalence of XBB* variants detected in clinical samples, GISAID sequences, and wastewater samples. A significant decline in XBB* variant is observed across all data sets over the study period. Notably, in October 2023, the XBB* variant signal reached 30% prevalence in both wastewater and clinical sequencing profiles, but while the signal continued to be detected in wastewater, it dropped out from the patient profiles. This highlights the sensitivity of wastewater surveillance in detecting ongoing variant circulation that may not be reflected immediately in clinical data.

The data from October 2023, showing XBB variants detected in wastewater and GISAID but not as prominently in clinical samples, may indicate changes in testing practices, particularly due to a significant decrease in testing observed in September 2023 (Figure 16). Analysis of the COVID ActNow dataset from the ROSALIND Tracker dashboard indicates that the average hospitalization data for Georgia State for the first half of September in 2021, 2022, and 2023 was 12,551, 2,863, and 798, respectively. This shows a downward trend of 94% from 2021 to 2023 and 72% from 2022 to 2023 (32). This aligns with the CDC report indicating that weekly hospital admissions for COVID-19 have decreased by more than 75% and deaths by more than 90% compared to January 2022, the peak of the initial Omicron wave (45). According to the report, more than 98% of the U.S. population now has some protective immunity against COVID-19 from vaccination, prior infection, or both. Additionally, the expiration of the COVID-19 public health emergency declaration on May 11, 2023, and the CDC’s updated respiratory virus guidance in March 2023, which reduced the isolation period to 24 hours instead of 5 days based on symptoms and not testing, impacted the number of people referring to clinics for testing (44,46).

If the number of clinical samples had remained constant, it could indicate that at this level, XBB might not be clinically relevant anymore, or the introduction of another variant might justify the discontinuation of XBB in the clinical trend line. In addition, it shows the wastewater testing provides the sensitivity to continue detecting variant signals, despite the lag in clinical sample testing.

Figure 20 highlights the critical role of wastewater surveillance as an early detection tool for emerging SARS-CoV-2 variants, which might or might not have clinical implications. During the study period, we observed two variants circulating at low levels in Georgia, but they did not appear to impact the clinical variant profile. The sharp spike in the BQ* variant detected in wastewater in May 2023, and the absence of a corresponding spike in clinical and GISAID data, raises questions about the possible reasons for this discrepancy. It may reflect a lag in clinical testing, differences in population sampling, or the variant’s low virulence, leading to fewer clinical cases despite widespread circulation. Since no lag in clinical testing or population sampling was reported during this period, it may be that the BQ* variant had lower virulence and was outcompeted by the XBB* variant.

**Figure 20:**
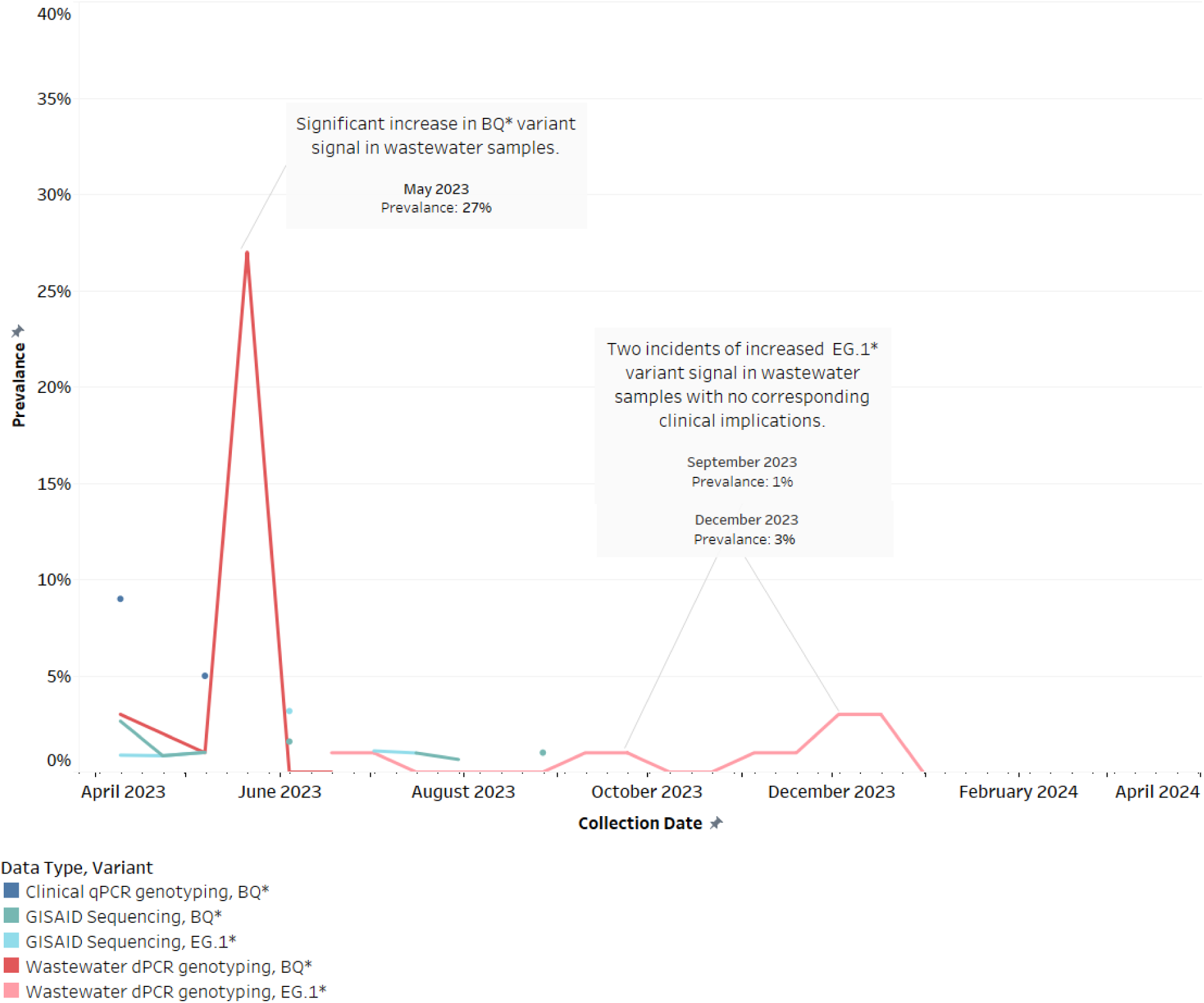
Prevalence of BQ and EG.1 Variants Across Wastewater, Clinical, and GISAID Data Sets in Georgia (April 2023 - April 2024). The graph illustrates the prevalence of BQ* and EG.1* variants detected in wastewater, clinical, and GISAID data sets. A notable spike in the BQ* variant is observed in wastewater samples in May 2023, reaching a prevalence of 27%, which is not reflected in clinical or GISAID data. The EG.1* variant was only detected in the wastewater data set during the fall and winter of 2023, with a prevalence of 1% in September and 3% in December, and showed no signal in the clinical and GISAID data sets. This highlights the sensitivity of wastewater surveillance in capturing emerging variants and underscores the importance of integrating multiple data sources for comprehensive monitoring.

The detection of the EG.1* variant solely in the wastewater data set during the fall and winter of 2023, with no corresponding signals in clinical or GISAID data sets, suggests that this variant did not lead to significant numbers of clinical cases.

The data in Figure 21 emphasizes the importance of wastewater surveillance as a leading indicator for the emergence and spread of SARS-CoV-2 variants. Between July and October 2023, the genotyping panel for clinical samples detected both EG.5 and FL, so we combined the two variants in all datasets to make the comparison more accurate. The graph indicates that EG.5/FL was detected in wastewater samples 22 days earlier than in clinical samples. However, the graph shows that it was detected in the GISAID dataset earlier in April, peaking in June 2023. The EG.5/FL assay was part of panel 2, implemented on June 21, 2023. However, investigating the Unknown marker from panel 1 revealed a spike in June 2023 at 20%, which aligns with the spike in the GISAID dataset (Figure 22). We suspect that the unknown spike in June 2023 is associated with the introduction of a new variant, FL.

**Figure 21:**
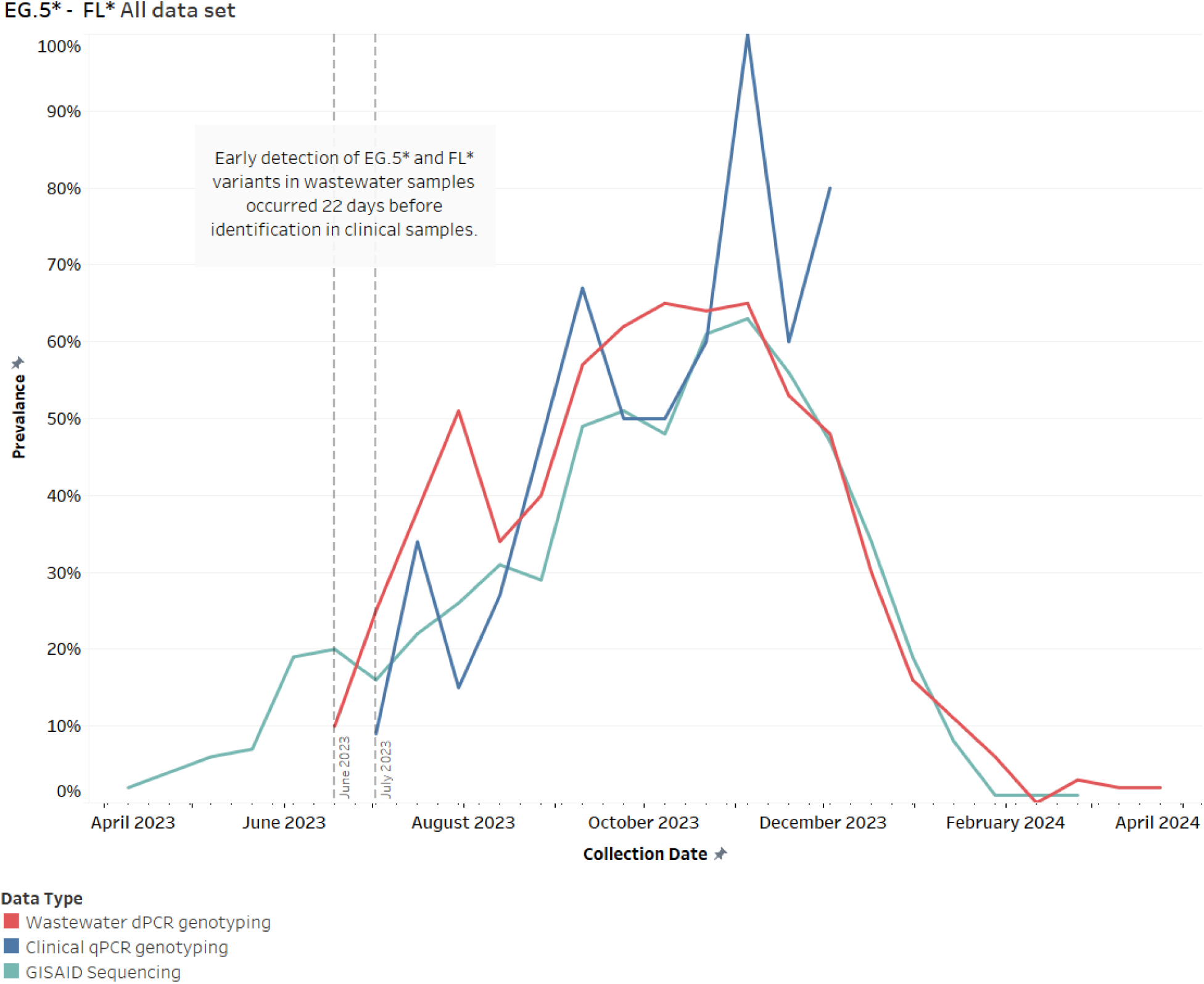
Prevalence of EG.5 and FL Variants Across Wastewater, Clinical, and GISAID Data Sets in Georgia (April 2023 - April 2024). The graph illustrates the prevalence of EG.5* and FL* variants detected in wastewater, clinical, and GISAID data sets. The data shows that EG.5/FL was detected in wastewater samples 22 days earlier than in clinical samples.

**Figure 22:**
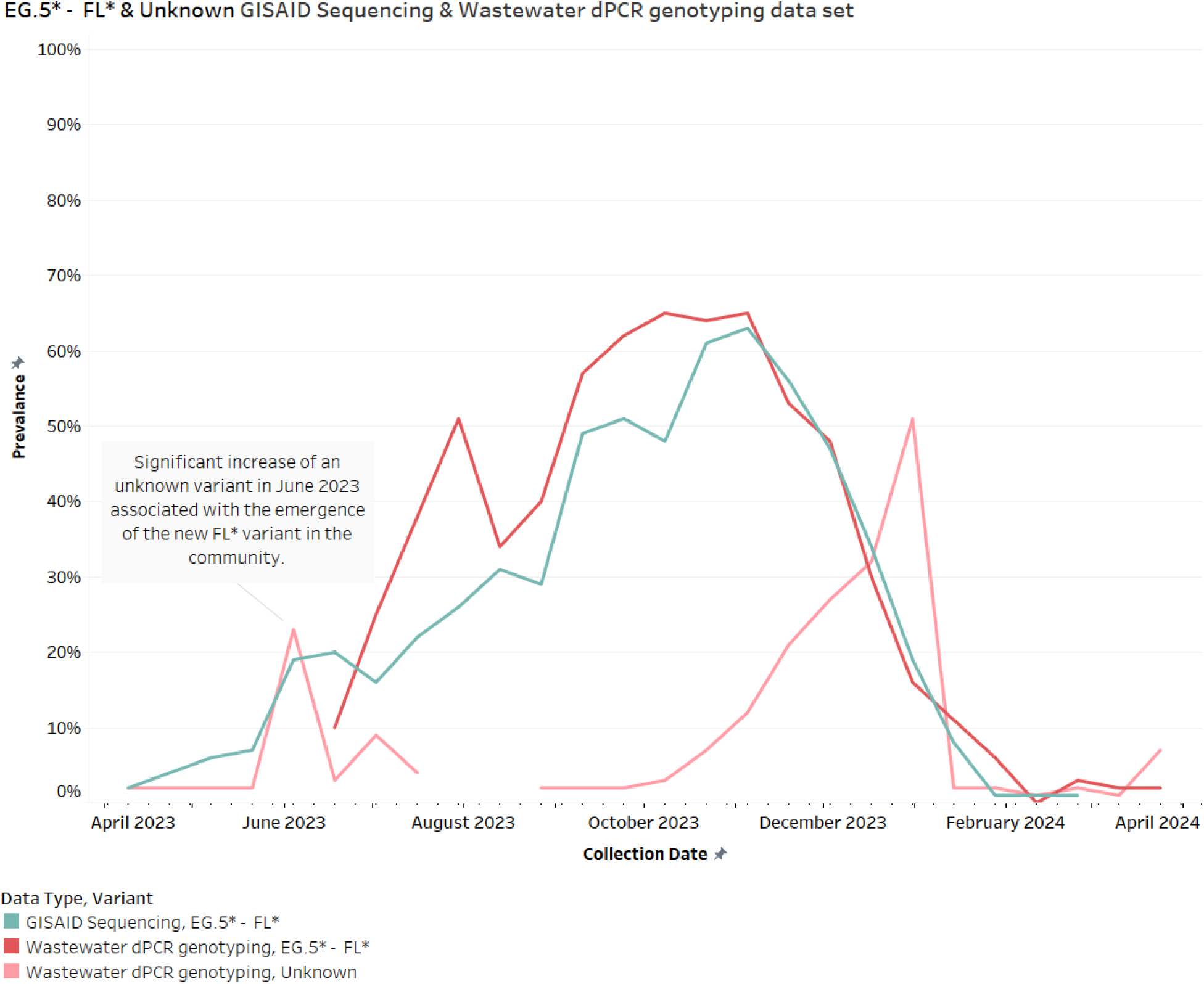
Prevalence of EG.5 and FL, and Unknown Variants in Wastewater and GISAID Data Sets in Georgia (April 2023 - April 2024). The graph illustrates the prevalence of EG.5* and FL* variants detected in wastewater samples and the GISAID data set, along with an unknown variant in the wastewater dataset. A notable spike in the unknown marker in June 2023 aligns with the spike in the GISAID data set, suggesting the introduction of a new variant, FL*.

The sudden disappearance of the trendline in the clinical dataset in December is associated with the reduction in the number of clinical samples tested, which occurred in the fall of 2023 and stopped in early 2024. However, wastewater can be used as an aggregated, non-invasive, and inclusive proxy for community-level infection trends as it closely matches the GISAID clinical dataset with less impact from the change in the number of samples.

The early detection of EG.5* and FL* variants in wastewater samples underscores the utility of wastewater surveillance in providing advance warnings, allowing for timely public health responses. The peaks observed in the datasets around October 2023, reaching approximately 65%, followed by a sharp decline, indicate a significant but short-lived surge in these variants’ prevalence, suggesting effective public health interventions or natural declines in transmission rates.

The second peak of the Unknown marker in the wastewater genotyping dataset was observed from October 2023 to January 2024 (Figure 23). Retrospective testing of wastewater samples collected between December 2023 and January 2024 strongly indicates that this spike in the Unknown marker is associated with the BA.2.86*/JN* variant (Figure 16). However, the JN variant was not detected in samples collected in October 2023. Low levels of JN were identified on November 5 and 6, 2023. Considering the trend observed in the GISAID dataset, we speculate that the JN variant was present in the community during October 2023 but was not detected in the specific sewersheds tested. Additionally, it is possible that the JN variant was present at levels below the detection threshold, given our requirement for a minimum number of positive partitions to classify a sample as positive.

**Figure 23:**
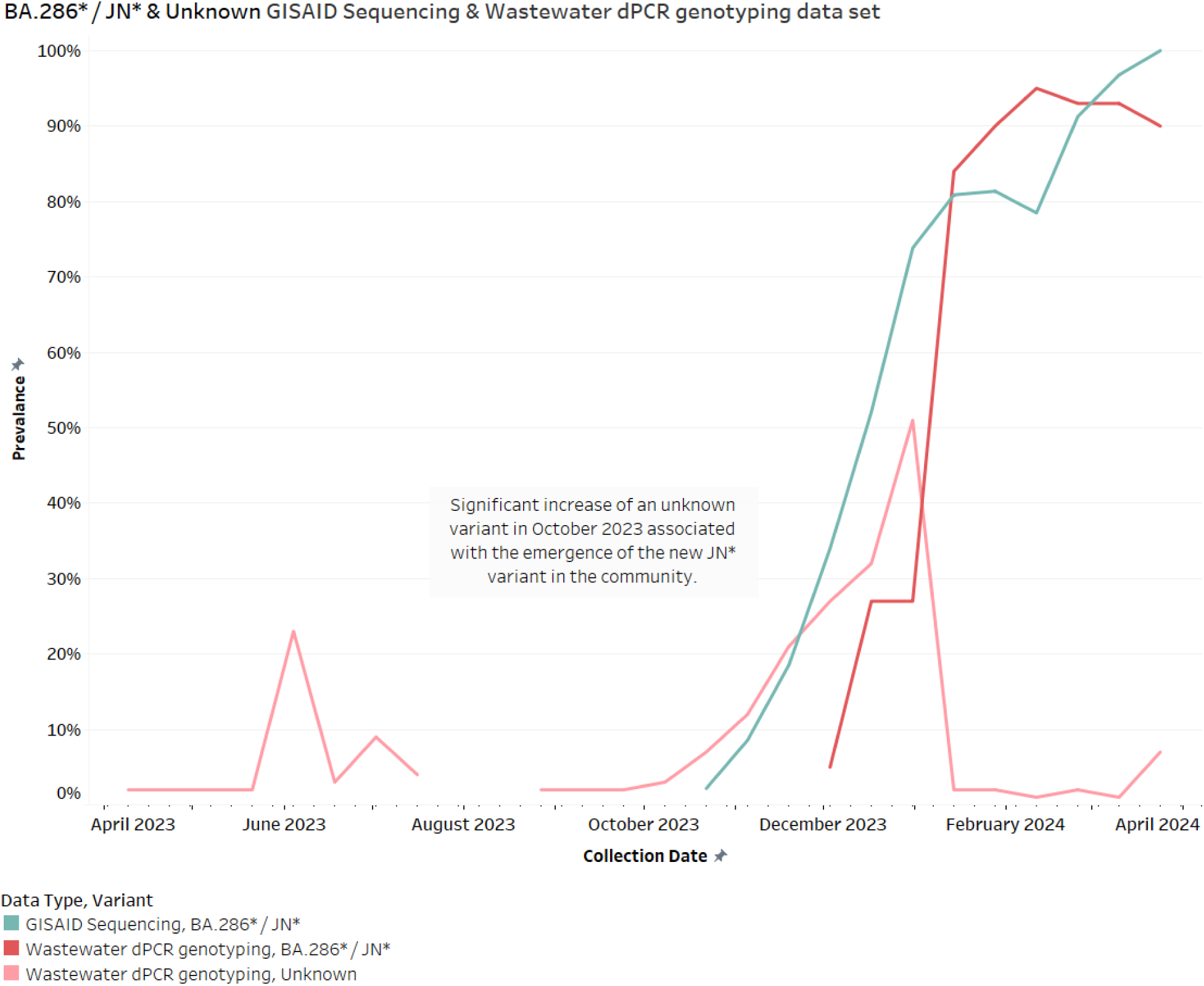
Prevalence of BA.2.86/JN and Unknown Variants in Wastewater and GISAID Data Sets in Georgia (April 2023 - April 2024). The graph illustrates the prevalence of the BA.2.86*/JN* variant and an unknown marker detected in wastewater samples and the GISAID data set. The second peak of the unknown marker in the wastewater dataset from October 2023 to January 2024 is associated with the BA.2.86*/JN* variant. The rapid rise and peak in BA.2.86*/JN* variant prevalence in early 2024 suggests a significant transmission event or enhanced transmissibility.

The combined rise of Unknown variants and JN variants from October 2023 to the end of the study period in April 2024 demonstrates a nearly perfect correlation in the trendlines between the wastewater and GISAID datasets. This alignment underscores the utility of wastewater genotyping surveillance in detecting and monitoring emerging SARS-CoV-2 variants.

The rapid increase and peak in the BA.2.86*/JN* variant prevalence in early 2024 suggests a significant transmission event or enhanced transmissibility of this variant, which is detectable through wastewater testing. However, the virulence of this variant remains unclear without understanding the shift in the number of people referring to clinics and hospitals. Unfortunately, the number of clinical samples in this study was limited during this period. However, the CDC reported that COVID-19-associated hospitalizations did not exceed those of the previous year, indicating no change in the virulence of the JN variant compared to other variants (45).

### Wisconsin State

#### a- Wisconsin Wastewater Genotyping

A total of 158 wastewater samples from Wisconsin state were tested between November 22, 2023, and February 1, 2024. Figure 24 illustrates the prevalence of various SARS-CoV-2 variants detected in these wastewater samples, presented in a bi-weekly cadence. The BA.2.86*/JN* variant (blue line) shows a significant increase, rising from less than 10% on November 19, 2023, to around 85% by January 24, 2024, indicating rapid dominance. The EG.5*/JG*/HK2*/HV* variant (red line) peaks at around 40% in early December 2023 before declining to less than 10% by the end of January 2024. Similarly, the XBB*/JD*/JF*/GK*/GJ*/FU*/HF*/GE* variant (purple line) fluctuates around 20-30% in November and early December, then decreases to less than 10% by late January. The Unknown variant (yellow line) remains relatively low, with a brief peak at 20% in early December before dropping below 10%. The EG.1* variant (orange line) appears only in early December at approximately 1%. The FL* variant (cyan line) starts at 10% in November, fluctuates, and decreases to 2% by January 28. The number of samples collected (black dotted line) is consistent, starting around 20 samples and maintaining an average of 26 samples in each bi-weekly assessment throughout the period. This data underscores the dynamic nature of variant prevalence and the critical role of wastewater surveillance in tracking the spread of SARS-CoV-2 variants in the community.

**Figure 24:**
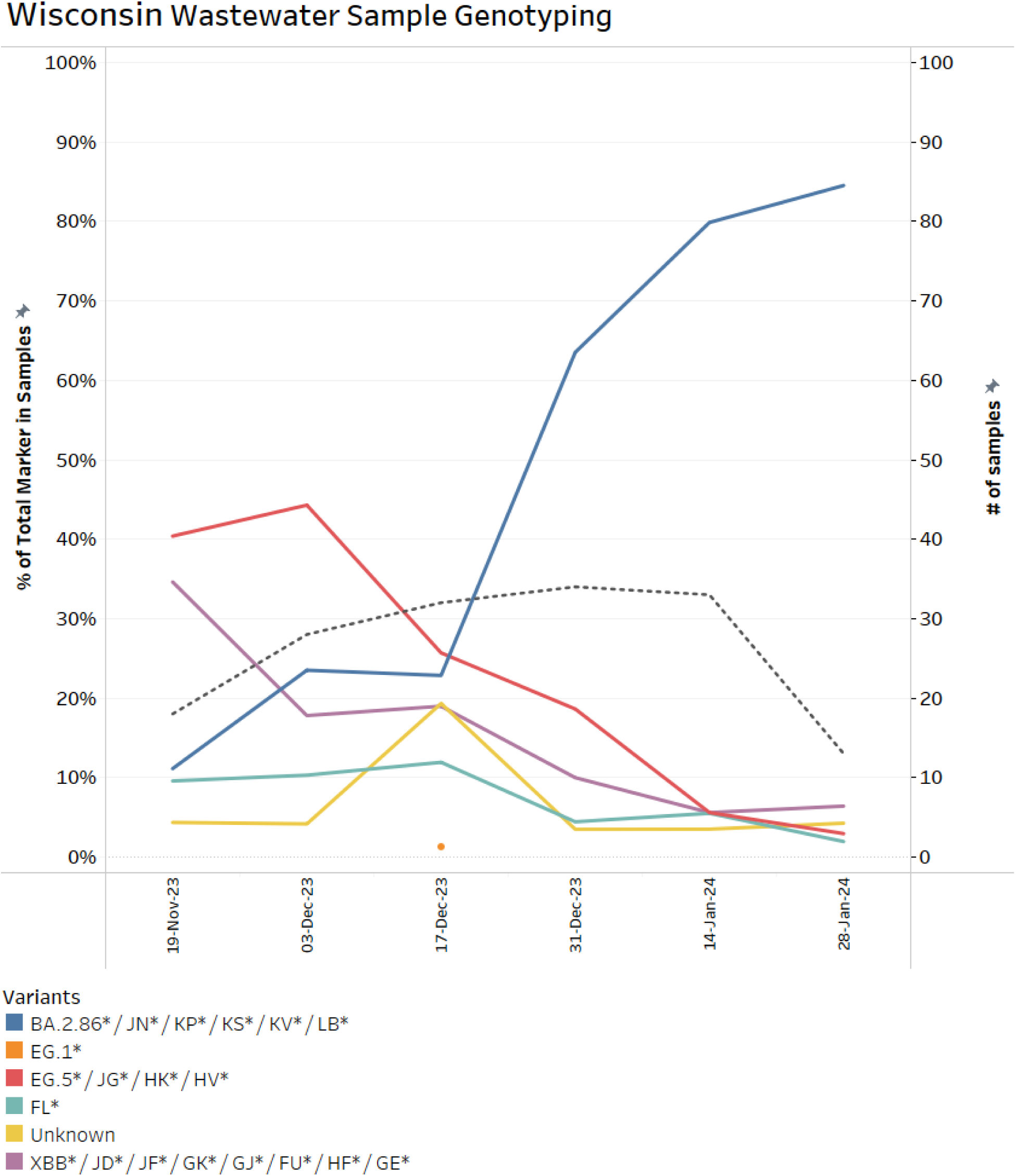
Prevalence of SARS-CoV-2 Variants in Wisconsin Wastewater Samples (November 2023 - February 2024). This graph shows the prevalence of various SARS-CoV-2 variants in 158 wastewater samples from Wisconsin, collected between November 22, 2023, and February 1, 2024. The BA.2.86*/JN* variant (blue line) rises sharply from less than 10% in November to around 85% in January, indicating rapid dominance. The EG.5*/JG*/HK2*/HV* variant (red line) peaks at 40% in early December before declining below 10%. The XBB*/JD*/JF*/GK*/GJ*/FU*/HF*/GE* variant (purple line) fluctuates between 20-30% in November and early December, then drops below 10%. The Unknown variant (yellow line) briefly peaks at 20% in early December, while the EG.1* variant (orange line) appears only in early December at 1%. The FL* variant (cyan line) starts at 10% in November and decreases to 2% by January. The number of samples collected (black dotted line) remains consistent, averaging 26 samples bi-weekly. This data highlights the dynamic nature of variant prevalence and the importance of wastewater surveillance in tracking SARS-CoV-2 spread.

#### **b-** Wisconsin Clinical Genotyping

Figure 25 displays the percentage of total SARS-CoV-2 variant lineages identified in clinical sample genotyping collected in Wisconsin from April 9, 2023, to March 2, 2024 presented in a bi-weekly cadence. Initially, the XBB* / JD* / JF* / GK* / GJ* / FU* / HF* / GE* variant (purple line) dominates nearly 100% in early April 2023 but steadily declines to 50% by mid-July 2023, fluctuating between 30-60% from August to early November before disappearing. The FL* variant (cyan line), EG.5/FL, and EG.5 variant appear in July 2023 and extend to January 2024. The EG.5* / JG* / HK* / HV* variant (red line) shows significant fluctuations, peaking around 75% in early November 2023. The EG1* variant did not appear in the clinical profile during this period. The Unknown variant (yellow line) shows sporadic presence with peaks around 20-60% in November and December 2023. The BA.2.86* / JN* variant (blue line) is almost non-existent until it suddenly dominates at around 100% in the final sample collection in January 2024. The number of samples collected (black dotted line) fluctuated, starting at 59 samples in a bi-weekly cadence in early April 2023 and gradually decreasing to about one sample by the end of February 2024. During this period, a total of 245 samples were genotyped. No samples were reported for March and April 2024.

**Figure 25:**
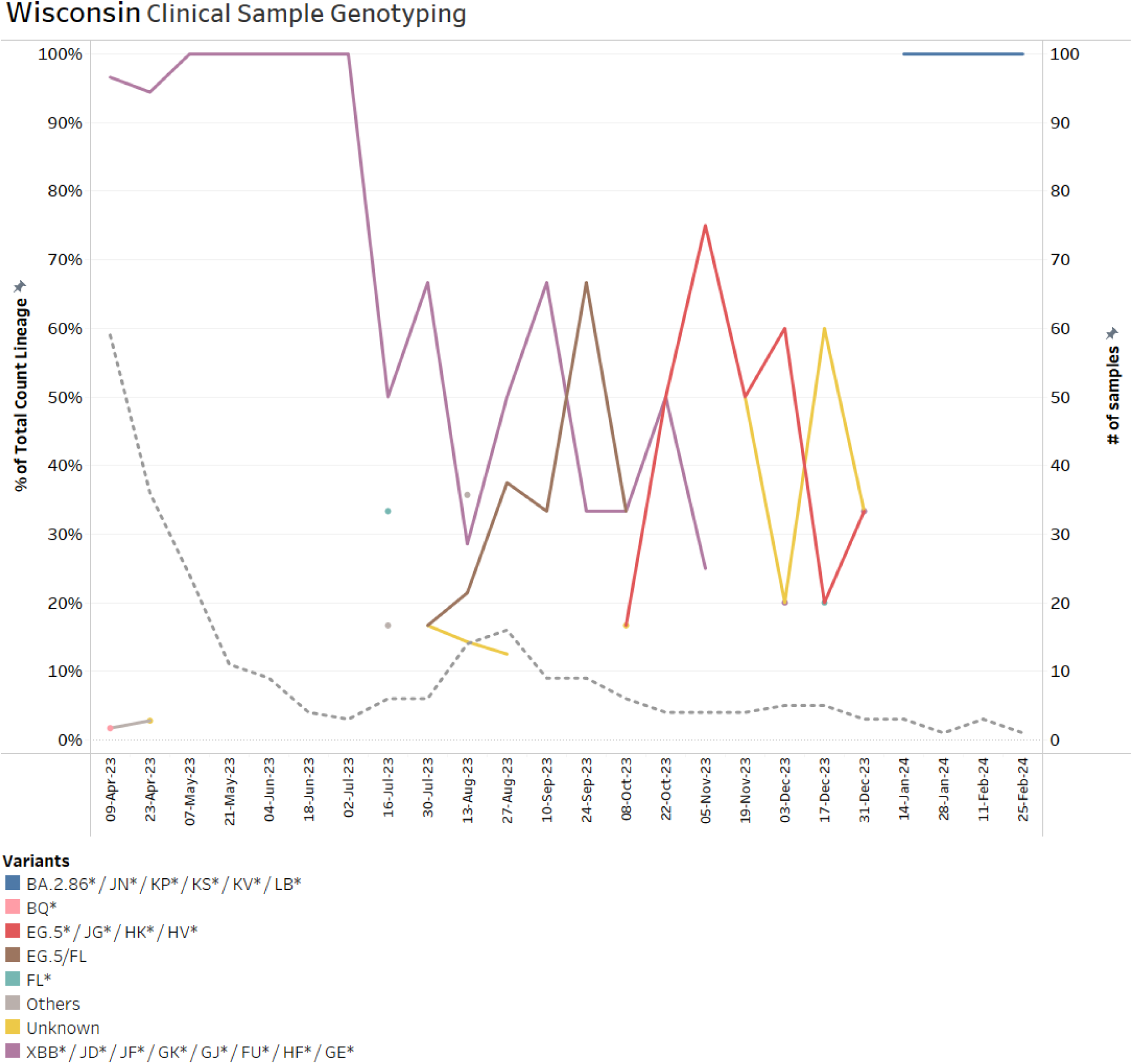
Percentage of SARS-CoV-2 Variant Lineages in Wisconsin Clinical Samples (April 2023 - April 2024). This figure shows the prevalence of SARS-CoV-2 variants in clinical samples from Wisconsin over a year. Initially, the XBB* variant dominates nearly 100% in April 2023 but declines to 50% by mid-July, fluctuating between 30-60% until early November before disappearing. The EG.5* / JG* / HK* / HV* variant peaks at 75% in early November 2023, while the FL* and EG.5 variants appear in July and persist until January 2024. The Unknown variant shows sporadic peaks up to 60% in November and December. The BA.2.86* / JN* variant is nearly absent until it dominates at 100% in January 2024. Sample collection starts at 59 in April 2023 and decreases to 1 by February 2024, with no samples reported in March and April 2024.

#### C. Wisconsin GISAID results

Figure 26 displays the percentage of total SARS-CoV-2 variant lineages identified in clinical samples presented in the GISAID database, collected in Wisconsin from April 9, 2023, to April 5, 2024. Initially, the XBB* group variants (purple line) dominate nearly 100% in early April 2023 but steadily decline below 10% in January 2024, becoming nearly non-existent by March 2024. In contrast, the BA.2.86* / JN* variant (blue line) starts low at the end of September 2023 but shows a sharp increase from November 2023, reaching around 90% by February 2024 and maintaining dominance through March 2024. The EG.5* group variants (red line) fluctuate significantly, peaking at 50% in November 2023 before declining to less than 10% by January 2024. The EG1* variant (orange line) appears in May and has a small peak of about 10% in June before disappearing at the end of July 2023. The FL* variant (cyan line) remains relatively low throughout, with minor fluctuations and peaks below 10%. The “Others” category (yellow line) shows sporadic presence with peaks around 10-20% at different times. This data highlights the dynamic changes in variant prevalence and underscores the importance of continuous genomic surveillance.

**Figure 26:**
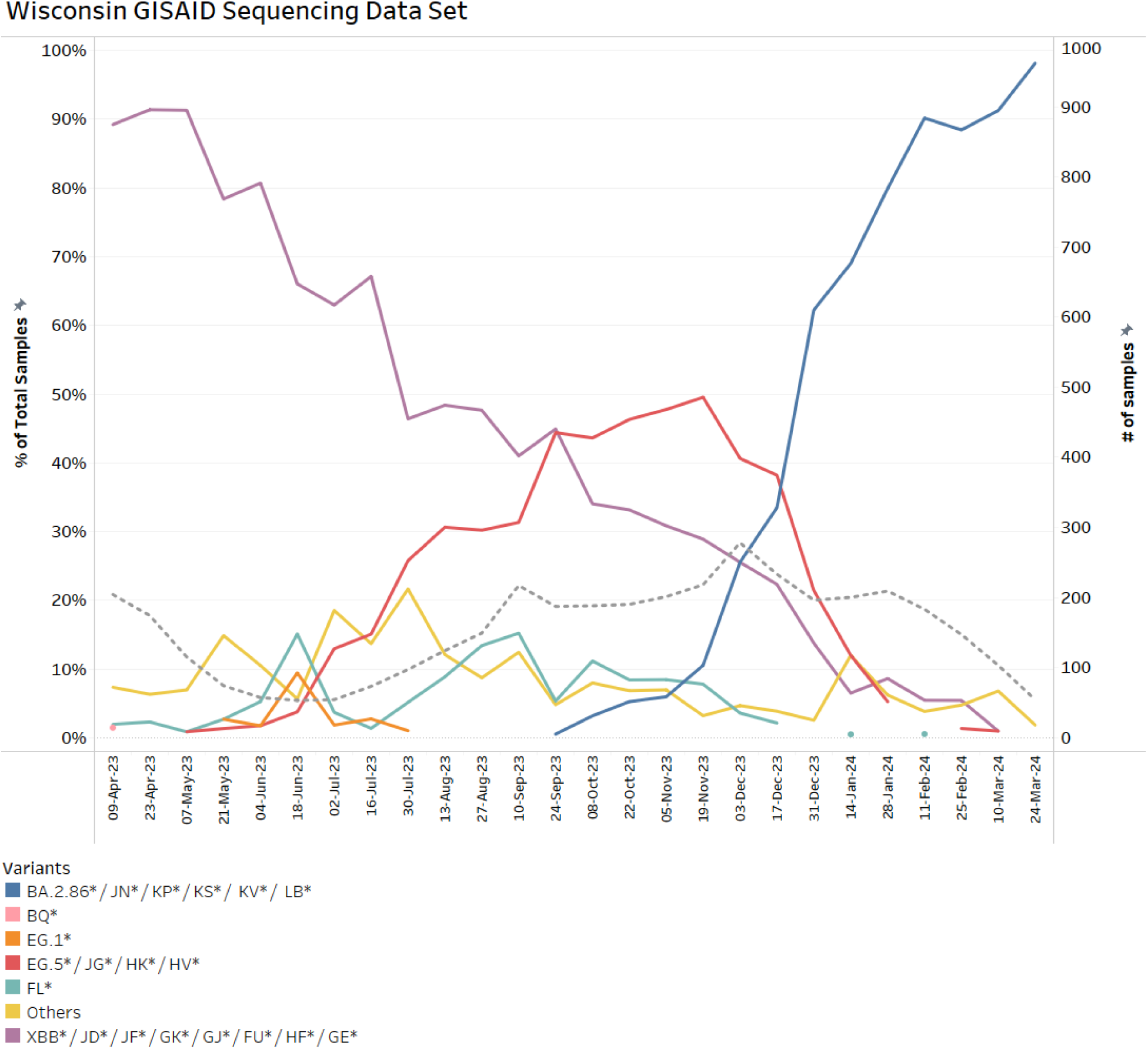
SARS-CoV-2 Variant Lineages in Wisconsin Clinical Samples on GISAID dataset (April 2023 - April 2024). This figure shows the percentage of SARS-CoV-2 variant lineages in clinical samples from Wisconsin, reported in the GISAID database over one year. Initially, the XBB* variants (purple line) dominate but decline below 10% by January 2024. The BA.2.86* / JN* variant (blue line) rises sharply from November 2023, reaching 90% by February 2024. The EG.5* variants (red line) peak at 50% in November 2023 before dropping below 10% by January 2024. The FL* and “Others” variants show low and sporadic presence throughout the period. The number of samples collected (black dotted line) averages 153 bi-weekly, totaling 3,978 samples, with a notable decline at the study’s end.

The number of samples collected (black dotted line) remained steady, with an average of 153 samples tested bi-weekly, showing a significant decline at the end of the study period. A total of 3,978 samples were reported during this one-year period.

#### D- Wisconsin Results Discussion

Figures 27 and 28 illustrate the dynamics of XBB and EG.5/FL variants in the wastewater and clinical genotyping datasets, as well as the GISAID dataset, in Wisconsin from April 2023 to April 2024. The wastewater testing does not cover the entire period, starting in November 2023 and continuing for three months until February 2024. Conversely, there were a limited number of clinical samples tested in the genotyping panel during these three months, averaging 4 samples per two weeks, with no clinical samples tested for genotyping in February and March 2024. Interestingly, the data shows a close resemblance between the trendlines of wastewater samples and the GISAID trendline for XBB and EG.5/FL markers. It appears that the wastewater genotyping data continues the clinical genotyping trend, filling the gap in clinical data despite the limited number of tested samples.

**Figure 27:**
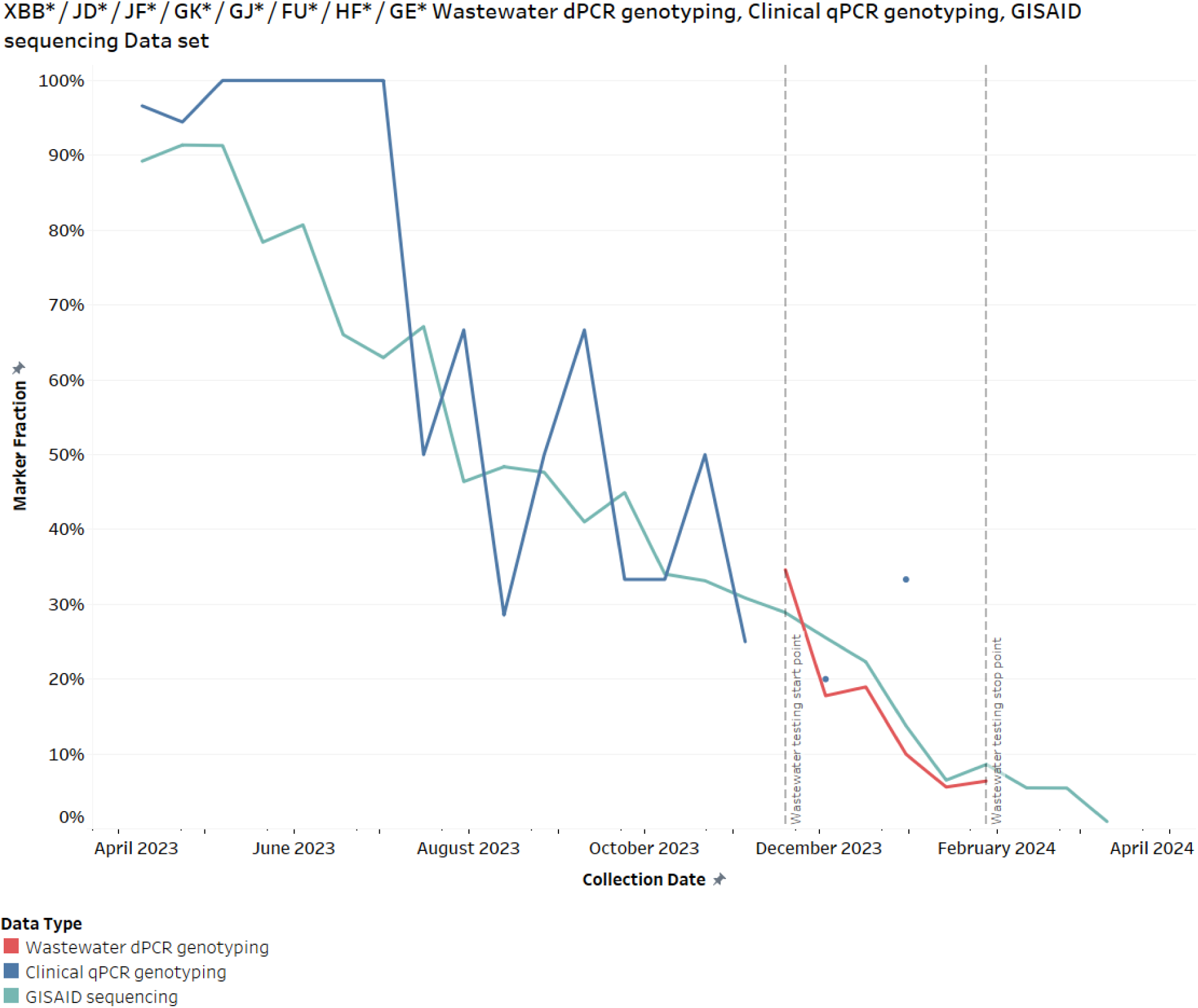
Prevalence of XBB Group Variants in Wisconsin Across Wastewater, Clinical, and GISAID Data Sets (April 2023 - April 2024). This figure shows the percentage of XBB group variant markers detected in Wisconsin’s wastewater (red line), clinical samples (blue line), and GISAID data (green line) from April 2023 to April 2024. All three data sets reveal a consistent decline in the prevalence of XBB group variants, starting near 100% in April 2023 and approaching 0% by April 2024. The strong correlation between local wastewater, clinical, and global GISAID data highlights the effectiveness of wastewater surveillance in tracking variant trends and filling the gap in clinical genotyping data.

**Figure 28:**
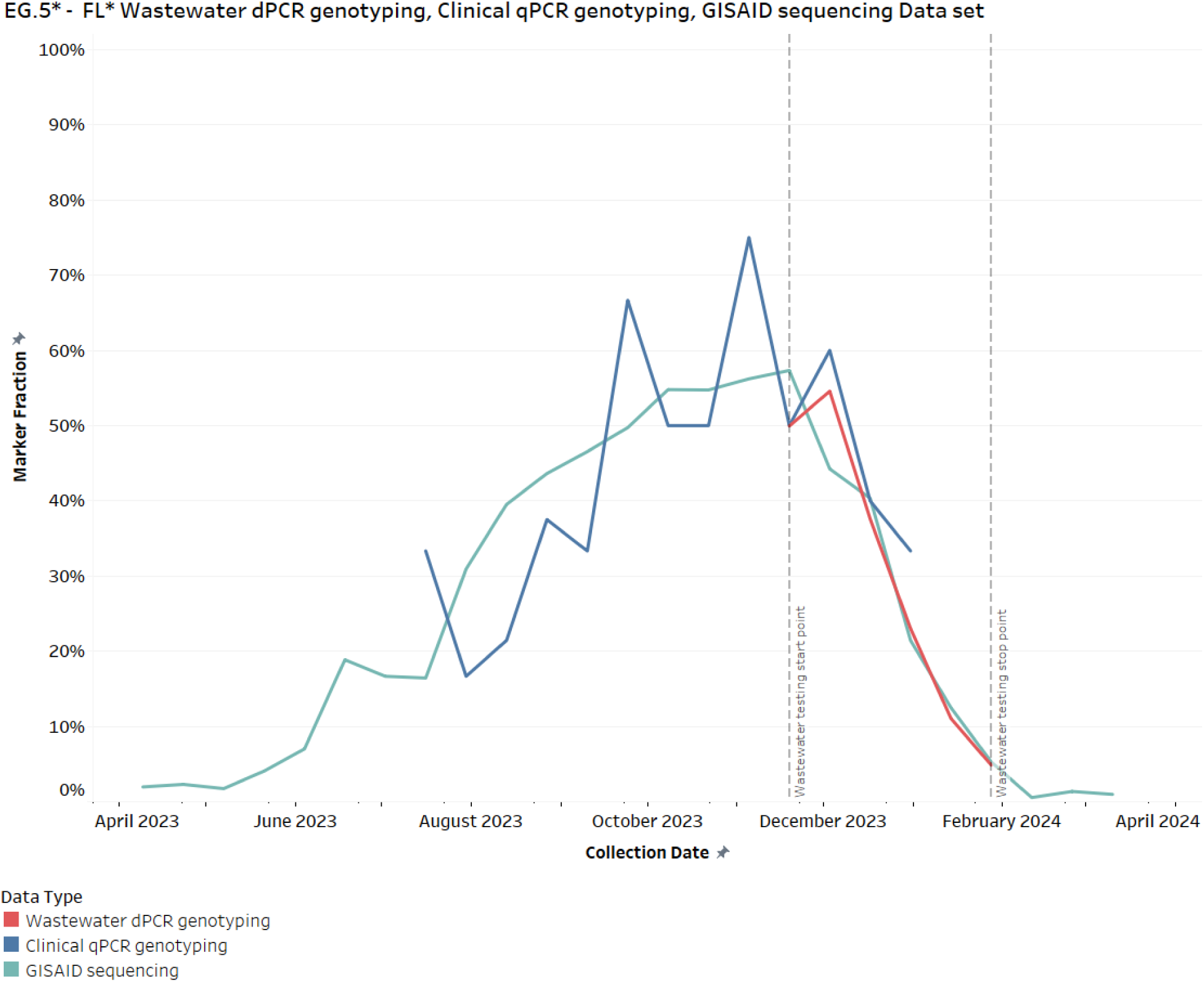
Prevalence of EG.5 - FL Variants in Wisconsin Across Wastewater, Clinical, and GISAID Data Sets (April 2023 - April 2024). This figure shows the percentage of EG.5* and FL* variant markers detected in Wisconsin’s wastewater (red line), clinical samples (blue line), and GISAID data (green line) from April 2023 to April 2024. All three data sets reveal a consistent rise in variant prevalence, peaking around December 2023, followed by a sharp decline to near 0% by February 2024. The strong correlation observed between wastewater, clinical, and GISAID data highlights the effectiveness of wastewater surveillance in tracking variant trends as a proxy for clinical variant genotyping.

Comparing the three datasets for the FL variant shows signal detection in both wastewater and GISAID in November 2023 (Figure 29). However, the GISAID data drops to 0% in December, while the wastewater samples continue to detect the variant. This suggests that the FL variant continued circulating in the community at low levels (<10%), but was not present in clinical samples that were sent for sequencing.

**Figure 29:**
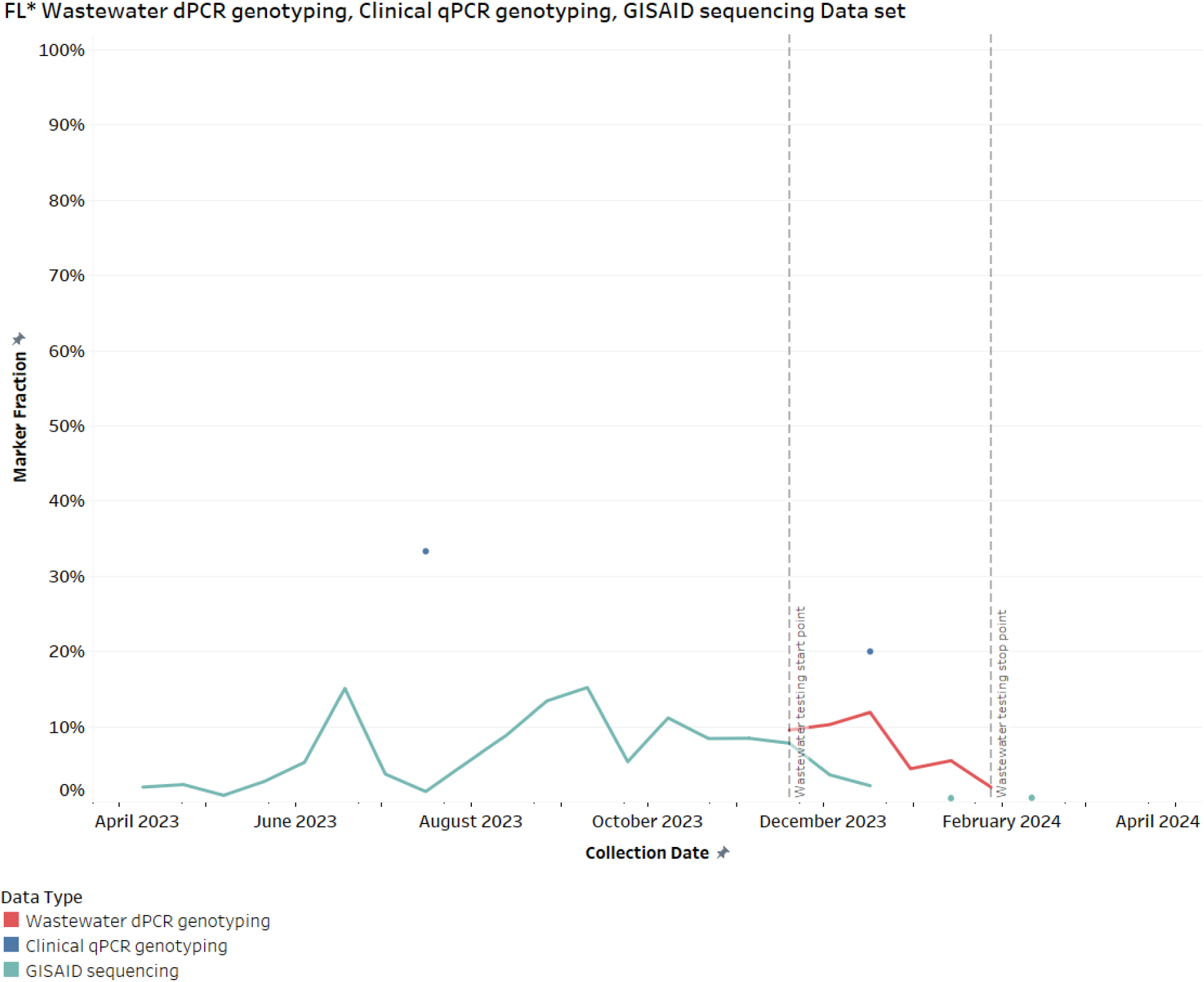
Prevalence of FL Variant in Wisconsin Across Wastewater, Clinical, and GISAID Data Sets (April 2023 - April 2024). This figure shows the percentage of FL* variant markers detected in Wisconsin’s wastewater (red line), clinical samples (blue line), and GISAID data (green line) from April 2023 to April 2024. Detection of the FL* variant in both wastewater and GISAID data occurs in November 2023, but it drops to 0% in GISAID data by December, while continuing to be detected in wastewater samples. This suggests that the FL* variant continued circulating in the community at low levels (<10%), with limited presence in clinical samples and, likely, hospitalizations.

The GISAID dataset indicates that the JN variant was detected at the end of September 2023 in Wisconsin and continued to dominate the variant profile through January 2024 (Figure 30). Retrospective JN analysis of wastewater samples from November to February shows a sharp rise in signal, matching the trend observed in the GISAID dataset. Sixteen wastewater samples tested with Panel 2 showed a spike in the Unknown marker during this period, indicating the presence of the new JN variant in the wastewater samples. Combining the prevalence of the Unknown variant with the retrospective JN variant prevalence indicates a slightly higher percentage of the JN variant circulating in the wastewater, but the trend closely follows the observed trend in the GISAID sequencing data.

**Figure 30:**
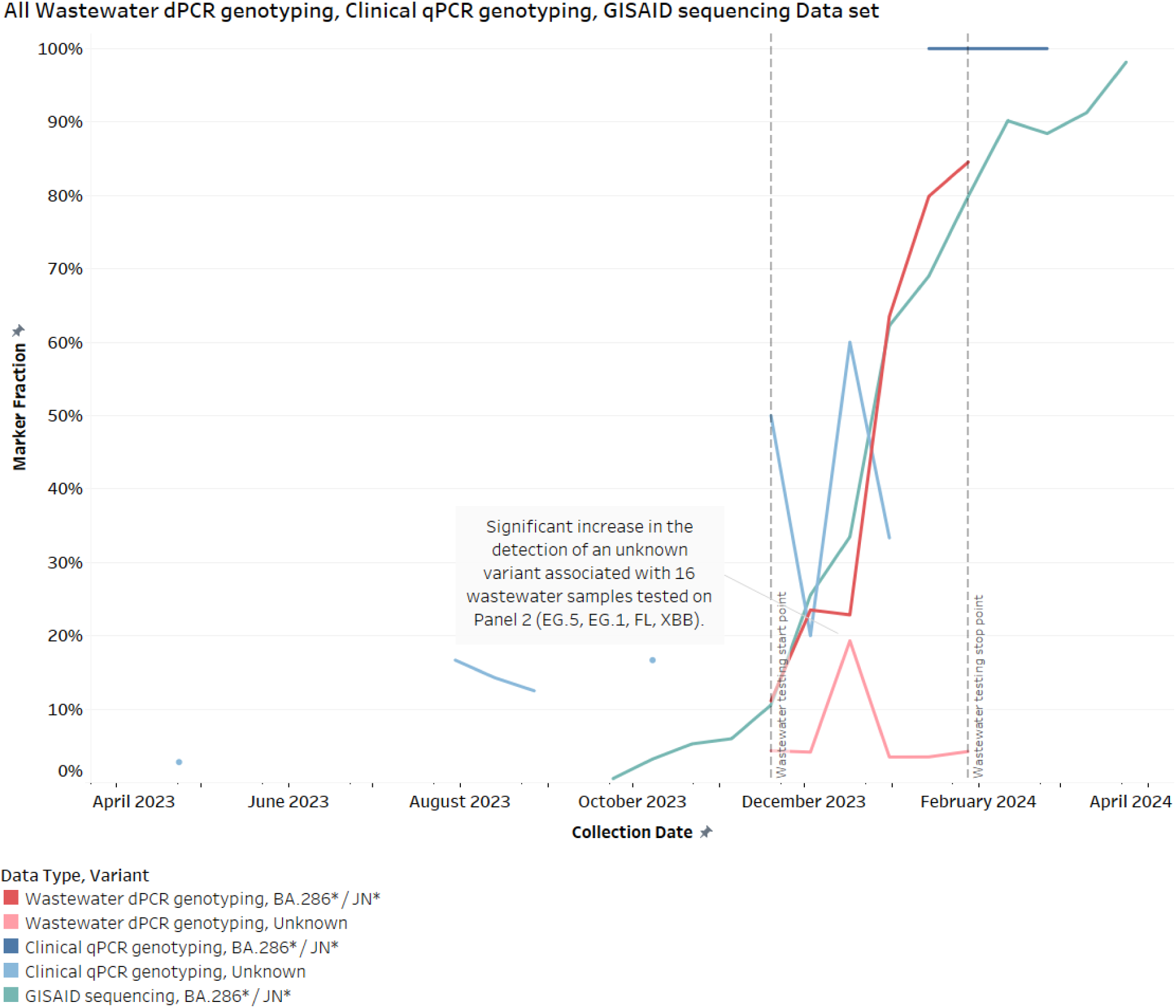
Prevalence of JN and Unknown Variants in Wisconsin Across Wastewater, Clinical, and GISAID Data Sets (April 2023 - April 2024). This figure shows the percentage of JN and BA.2.86* variant markers detected in Wisconsin’s wastewater (red line for JN and pink line for Unknown), clinical samples (dark blue line for JN and light blue line for Unknown), and GISAID data (green line for JN) from April 2023 to April 2024. The Unknown signal was detected during the wastewater testing period. Retrospective analysis of wastewater samples from November to February shows a sharp rise in the JN signal, aligning with the GISAID trend.

#### National Level Comparative Analysis of SARS-CoV-2 Variant Detection Across Wastewater, Clinical Genotyping, and GISAID Sequencing

More than 1,400 wastewater samples from six different states were processed and analyzed using the dPCR genotyping approach from April 2023 to May 2024. In comparison, over 15,000 clinical samples nationwide were analyzed using qPCR-based genotyping, with more than 300,000 clinical sequencing results deposited in the GISAID database. We compared the wastewater genotyping results from six states—New York, Georgia, Illinois, Wisconsin, and California—representing the Northeast, South, Midwest, and West regions of the United States, to the national-level clinical genotyping effort and deposited SARS-CoV-2 sequencing data in the GISAID database for the period of October 1, 2023, to May 1, 2024. During this period, 1,219 wastewater samples, 3,031 clinical samples, and 156,431 sequencing results were analyzed. Despite genotyping fewer wastewater samples, we observed a similar trend in variant dynamics across the three datasets (Figure 31). The rapid rise and dominance of the JN.1 variant in November 2023 were consistently observed in wastewater samples. The comparative analysis of wastewater genotyping, clinical genotyping, and GISAID sequencing data demonstrates the effectiveness of wastewater surveillance as an early warning system for SARS-CoV-2 variants. The integration of these datasets provides a comprehensive view of variant dynamics, supporting informed public health decision-making and response strategies.

**Figure 31:**
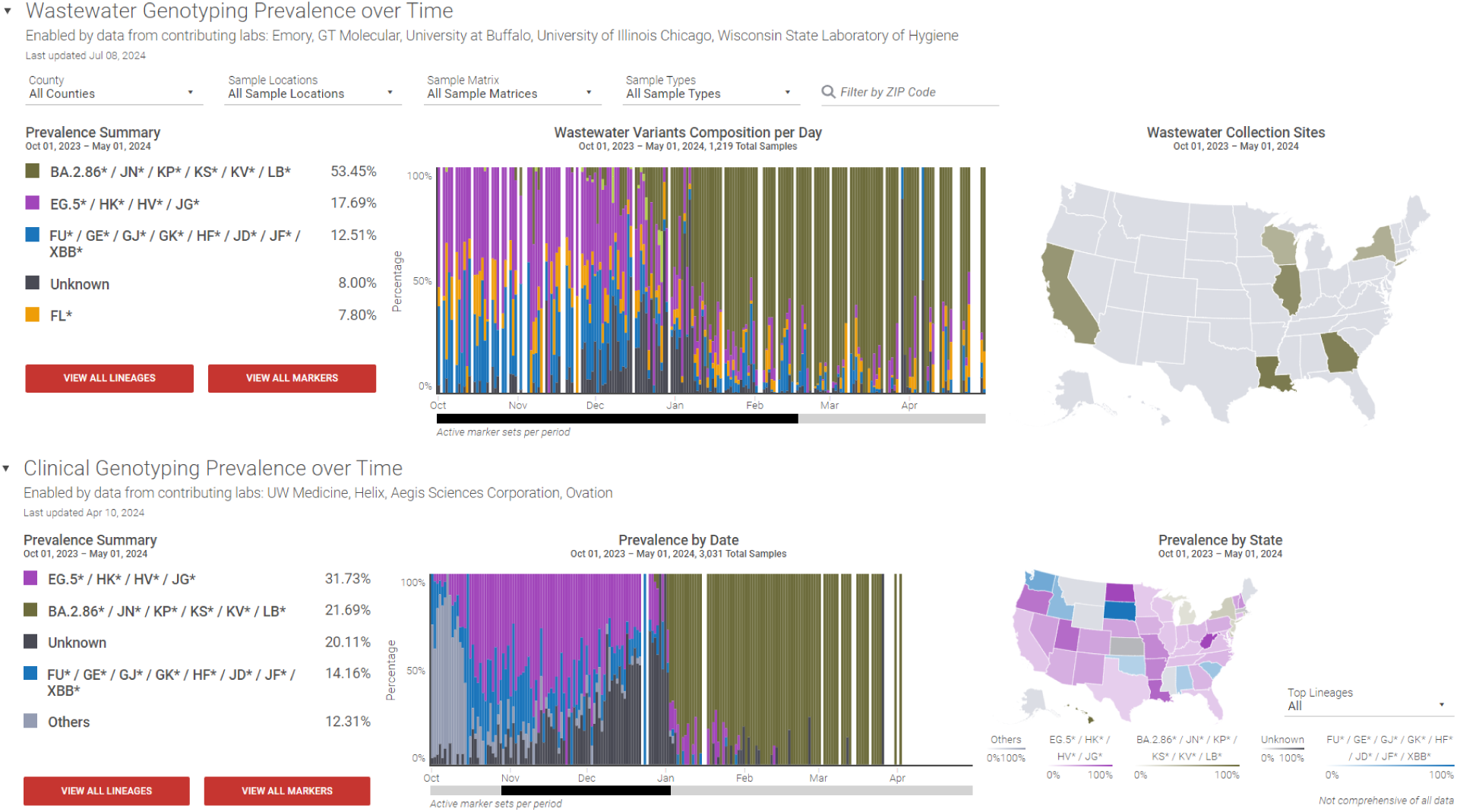
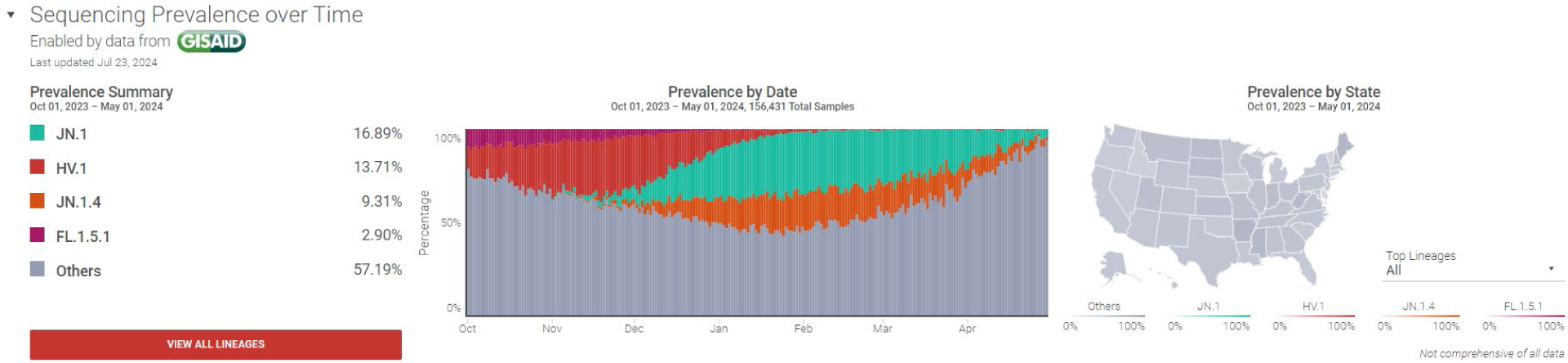
National Comparison of SARS-CoV-2 Variant Prevalence Across Wastewater, Clinical, and Sequencing Data Sets (October 2023 - May 2024). Top: Wastewater Genotyping Dashboard illustrates the prevalence of SARS-CoV-2 variants in wastewater samples collected across six states. Middle: Clinical Genotyping Dashboard presents the variant prevalence from clinical samples across the United States. Bottom: GISAID Sequencing Dashboard displays sequencing data from the GISAID database, covering a wide range of SARS-CoV-2 variants. All three datasets indicate the steady rise of JN.1 and JN* variants starting in November 2023, replacing EG.5*/HV.1 variants and becoming dominant by early 2024. Note that only the top four lineages/variants in this period are listed. Snapshot image from ROSALIND Tracker (32).

#### Performance comparison between Wastewater dPCR genotyping method and Whole Genome Sequencing Approach

We also wanted to compare the performance of the dPCR genotyping method to next generation sequencing. We compared the data from ROSALIND wastewater dashboard to Wisconsin SARS-CoV-2 Wastewater Genomic Dashboard. The Wisconsin State Laboratory of Hygiene WSLH sequences about 20% of the samples that are tested for routine COVID-19 wastewater surveillance (47). Details on the sequencing protocol can be found in the method section, but briefly, Illumina WGS data are processed through the Viralrecon workflow (48). The bioinformatics algorithm Freyja is used to evaluate the relative proportion of the SARS-CoV-2 lineages present in wastewater samples. Data are manually curated to only display the lineages according to WHO and Nextstrain nomenclatures. These data and visualizations are available on a dashboard accessible to the public: https://dataportal.slh.wisc.edu/sc2-ww-dashboard.

For this study, the Wisconsin State Laboratory of Hygiene (WSLH) processed a biological duplicate of 145 wastewater samples; One was conducted based on their routine wastewater genomic surveillance using the following automated workflow: 10 mL of untreated influent was concentrated using Microbiome A Nanotrap particles (Ceres) and extracted using the Maxwell(R) HT Environmental TNA kit (Promega), and Illumnia WGS. The other duplicate was processed following the SOP established for Panel 2 for the dPCR genotyping study. In response to the new circulating variant, 129 of dPCR genotyping elutions were retrospectively assayed to obtain the JN variant fraction. The results of both methods were uploaded on ROSALIND and Wisconsin SARS-CoV-2 Wastewater Genomic Dashboard for visualization. We investigated the results of 129 samples which were processed in both methods. Data from both dashboards extracted and aggregated according to table 8 for correlation analysis purposes.

**Table 8:** Synchronized markers from the Wisconsin State Wastewater Surveillance Dashboard and the ROSALIND Wastewater Genotyping Dashboard for 145 unique wastewater samples.

| Rosalind Wastewater Genotyping Dashboard | Wisconsin Wastewater Sequencing Dashboard |
| --- | --- |
| XBB* / JD* / JF* / GK* / GJ* / FU* / HF* / GE* | XBB.2.3<br>XBB.1.16<br>XBB.1.9<br>XBB.1.5.70<br>XBB.1.5<br>XBB |
| EG.5* / JG* / HK* / HV* | HK.3<br>EG.5.1 |
| BA.2.86* / JN* / KP* / KS* / KV* / LB* | BA.2.86 |
| FL* | - |

At first, we conducted an agreement analysis between two methods for the markers on the Panel 3 (JN, EG.5, FL and XBB). We looked at both positive and negative detection in the wastewater samples. The agreement ranges from 54.3% to 96.9%. observed Table 9 summarizes the results:

**Table 9:** Performance of dPCR genotyping against WGS method for JN, EG.5, XBB and FL variants. Wastewater samples were counted based on the detection or absence of the marker in each method.

|  | Variant | # of agreement | # of disagreement | Percent agreement * |
| --- | --- | --- | --- | --- |
| dPCR genotyping vs. WGS | JN | 125 | 4 | 96.9 % |
|  | EG.5 | 109 | 20 | 84.5 % |
|  | XBB | 89 | 40 | 69 % |
|  | FL | 70 | 59 | 54.3 % |

\* Combined positive and negative percent agreement.

The lower percentage of agreement for the XBB and FL markers likely can be attributed to the specific nature of SNP detection methods in dPCR genotyping and the nomenclature protocols used in sequencing methods. Recombinant variants, such as XBB, often result from the fusion of multiple variants, incorporating mutations from variants like FL. These mutations can be detected in the dPCR SNP assay, yet they are still classified under the broader category of XBB. This dual detection can lead to discrepancies between dPCR and WGS results. Additionally, for variants like XBB, where prevalence calculation depends on multiple markers, the agreement is poorer due to errors from more measurements (e.g., 2-3 measurements instead of 1) and the percentage normalization step.

In contrast, markers like JN likely show higher agreement due to the presence of new mutations not shared with other variants, leading to more consistent identification across both methods. Despite these differences, the overall agreement remains strong when considering the inherent variation between biological samples and the distinct methodologies of dPCR genotyping and WGS. This highlights the robustness of the genotyping assays despite the complexities involved in detecting recombinant and novel variants.

Since the wastewater samples often exhibit a profile of different variants, we performed a correlation analysis on the JN, EG.5 and XBB markers to study the prevalence calculation between two methods: dPCR genotyping approach vs. Illumina WGS. None of the samples were reported positive for FL in the sequencing dashboard therefore is not included in this analysis. Figure 32 illustrates the correlation between the dPCR and WGS for prevalence of three markers in 129 samples:

**Figure 32:**
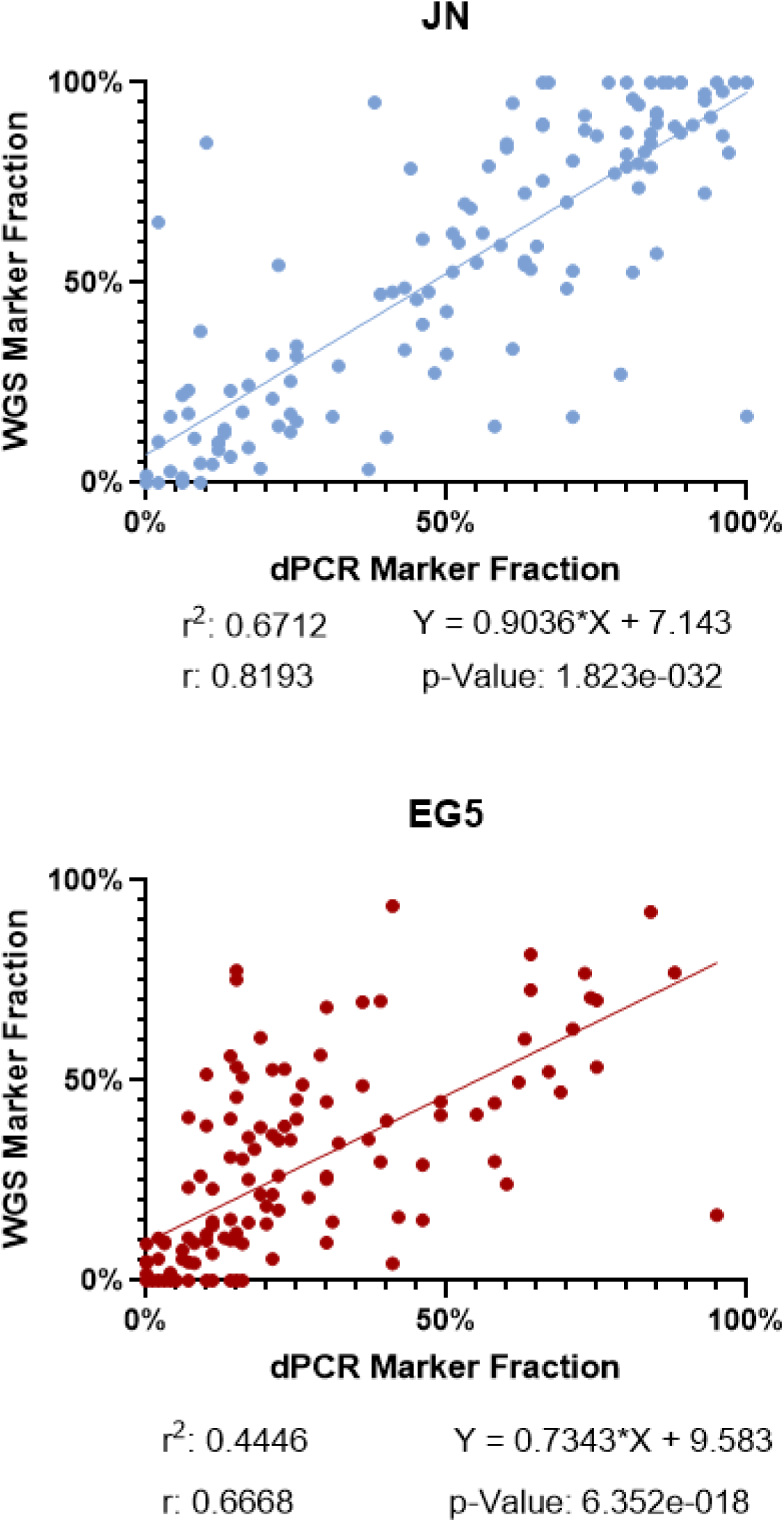
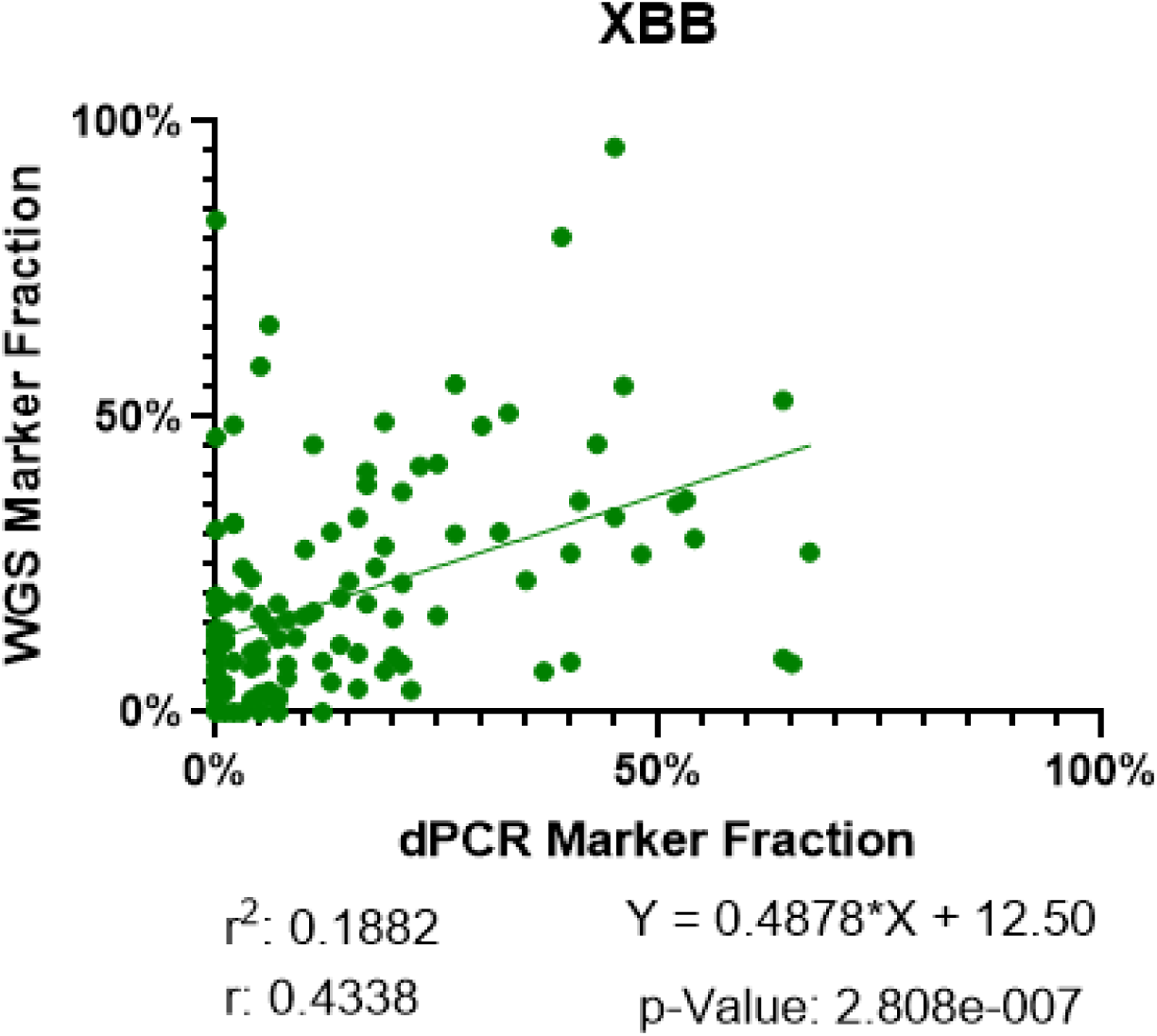
Correlation analysis between dPCR and WGS on 129 wastewater samples. Top: JN marker shows strong correlation and strong consistency, Middle: EG.5 demonstrates moderate correlation, bottom: XBB demonstrates moderate correlation with higher variability.

The top graph demonstrates a strong positive correlation between the fraction of markers detected by digital PCR (dPCR) and Whole Genome Sequencing (WGS) for the JN variant of SARS-CoV-2. The data points show a clear upward trend, with a correlation coefficient (r) of 0.8193 and a coefficient of determination (r²) of 0.67, indicating that 67.12% of the variability in WGS marker fraction can be explained by the dPCR marker fraction. The regression line equation (Y = 0.9036*X + 7.143) suggests that for every 1% increase in the dPCR marker fraction, the WGS marker fraction increases by approximately 0.90%, with an intercept of 7.14%. The extremely low p-value (1.823e-032) confirms the statistical significance of this correlation, making it highly unlikely to be due to random chance. The data points are closely clustered around the regression line, indicating a strong consistency between dPCR and WGS in detecting the JN variant, with WGS generally detecting slightly higher marker fractions.

EG.5 data points exhibit a moderate to strong positive relationship, as indicated by the correlation coefficient (r) of 0.6668 and a coefficient of determination (r²) of 0.4446, meaning that approximately 44.46% of the variability in WGS marker fraction can be explained by the dPCR marker fraction. The regression line equation (Y = 0.7343*X + 9.583) suggests that for every 1% increase in the dPCR marker fraction, the WGS marker fraction increases by about 0.7343%, with an intercept of 9.583%. The extremely low p-value (6.352e-018) confirms the statistical significance of this correlation, indicating that the observed relationship is unlikely to be due to random chance. Despite some variability in the data, the overall trend supports a consistent detection of the EG.5 variant by both dPCR and WGS, with the WGS marker fraction generally being slightly higher than the dPCR marker fraction.

For the XBB variant of SARS-CoV-2, the correlation coefficient (r) of 0.4338 and the coefficient of determination (r²) of 0.1882 indicate that only 18.82% of the variability in the WGS marker fraction can be explained by the dPCR marker fraction. The regression line equation (Y = 0.4878*X + 12.50) suggests that for every 1% increase in the dPCR marker fraction, the WGS marker fraction increases by approximately 0.4878%, with an intercept of 12.50%. The p-value of 2.808e-007 confirms the statistical significance of this correlation, making it highly unlikely to be due to random chance. Despite the moderate strength of the correlation and significant variability in the data, the overall trend indicates that both dPCR and WGS are consistent in detecting the XBB variant, with WGS generally detecting higher marker fractions compared to dPCR.

The analysis of the three graphs for the JN, XBB, and EG.5 variants of SARS-CoV-2 reveals varying degrees of correlation between the fractions of markers detected by dPCR and WGS. For the JN variant, there is a strong positive correlation, indicating a high level of agreement between dPCR and WGS. The EG.5 variant also shows a statistically significant positive correlation, though slightly weaker, demonstrating moderate agreement between the two methods. In contrast, the XBB variant exhibits a moderate positive correlation with a lower r of 0.4338 and an r² of 0.1882, indicating substantial variability and less agreement between dPCR and WGS. As explained in previous analyses, besides differences in methodology, detection sensitivity, and baseline detection levels, the labeling of recombinant variants such as XBB, which combine mutations from different lineages, can lead to inconsistent results compared to SNP-based methods like the dPCR genotyping approach. Overall, while dPCR and WGS show strong consistency in detecting the JN variant and moderate consistency for EG.5, the XBB variant’s detection reveals more discrepancies, reflecting the inherent variability and challenges in consistently detecting certain recombinant SARS-CoV-2 variants.

### dPCR genotyping vs. NGS sequencing: Cost-Time-Labor analysis

Due to the complex library preparation protocols and the time-intensive analysis inherent in next-generation sequencing (NGS), it is often more expensive and has a longer turnaround time compared to RT-qPCR (49–51). We performed a side-by-side comparison between the cost of the dPCR genotyping method used in this study to the whole-genome sequencing (WGS) amplicon-tiled sequencing approach on the Illumina platform by NGS technology, which often is considered the gold standard for SARS-CoV-2 variant detection in wastewater (24). Our calculation is based on 17 wastewater samples at a time, which is the throughput of the wastewater genotyping method we developed. Table 10 summarizes the findings.

**Table 10:**
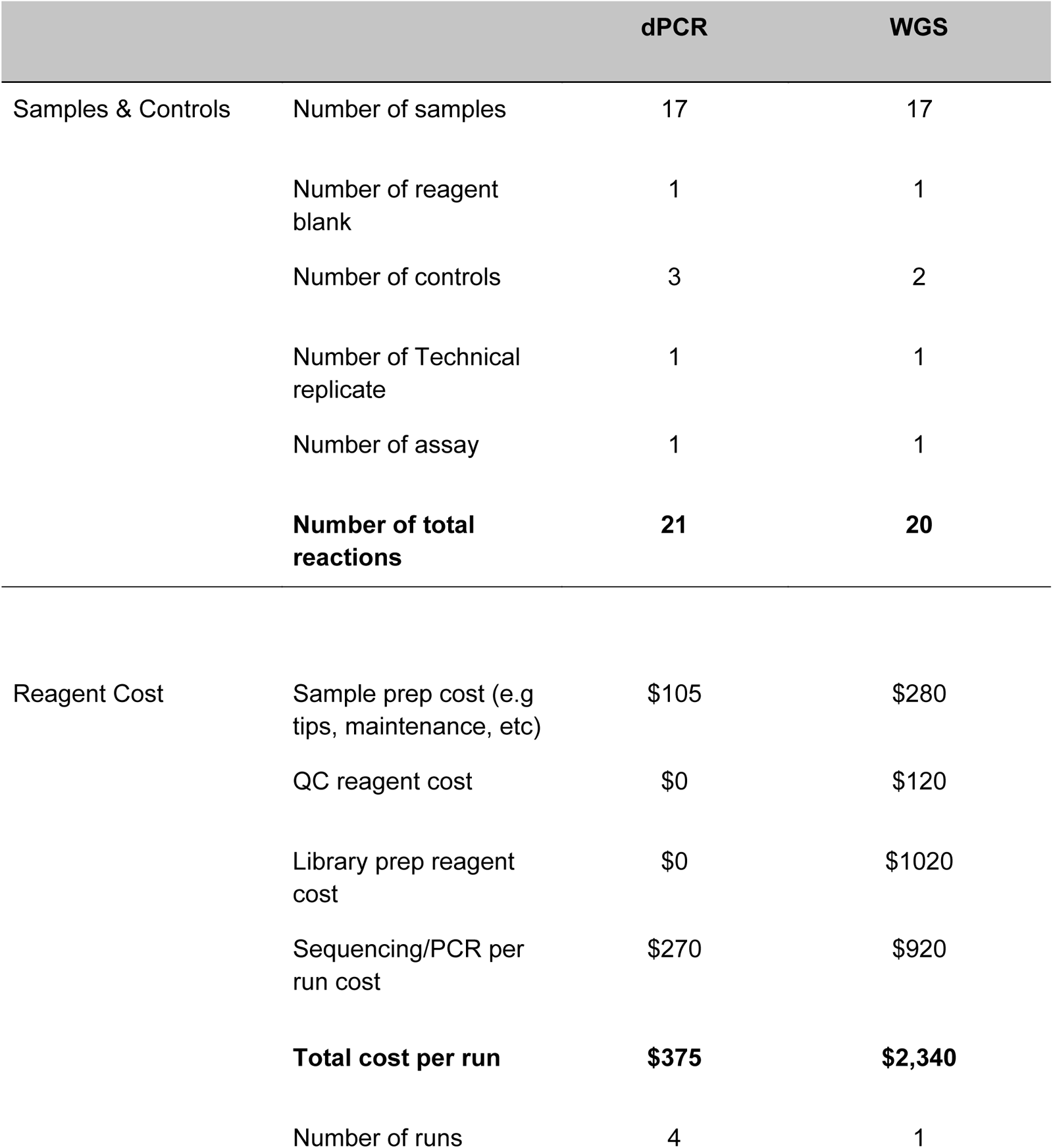
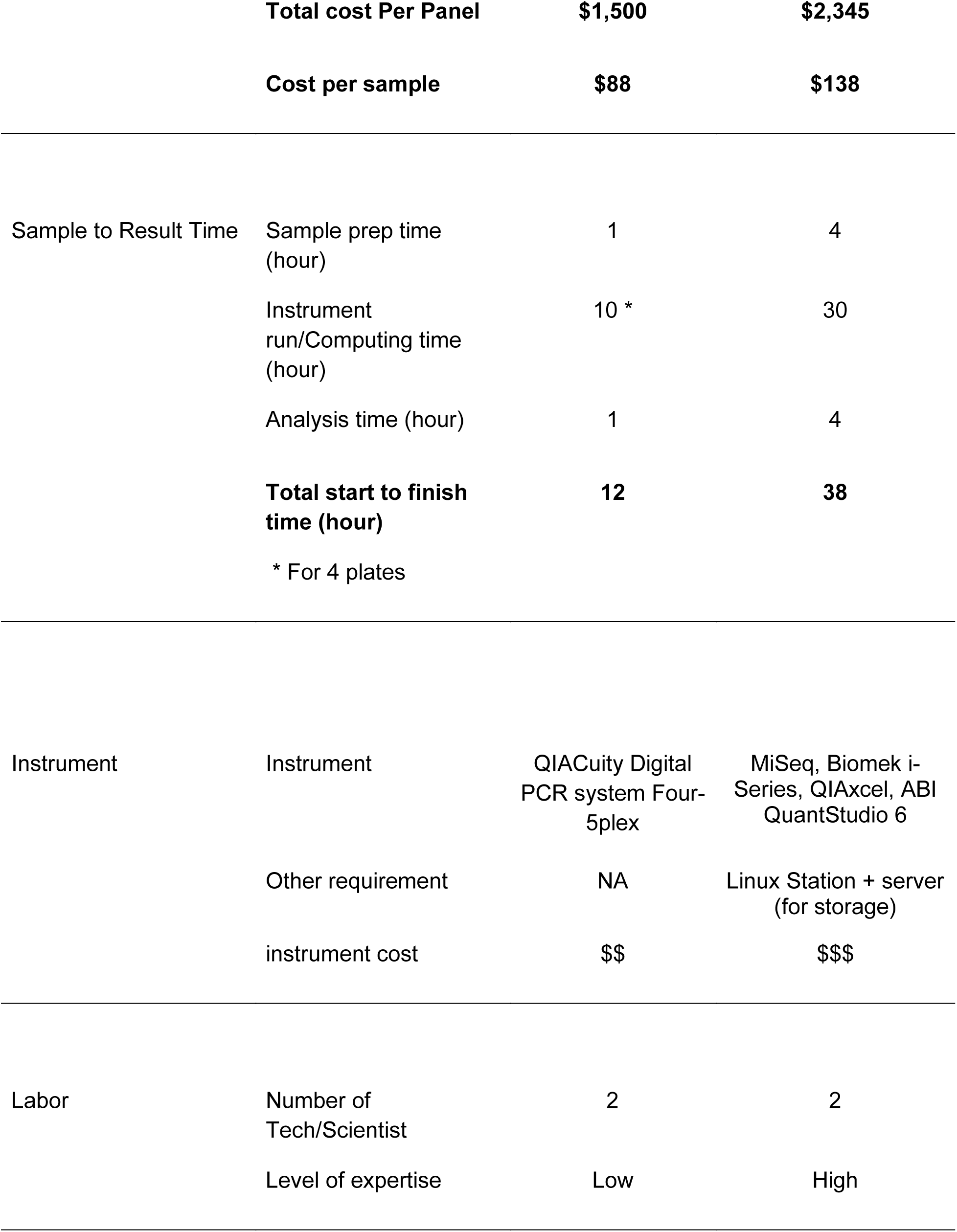
Comparison of dPCR Genotyping Method and WGS Amplicon-Tiled Sequencing Approach. This table compares the costs, sample processing requirements, and time to results for the dPCR genotyping method and the whole-genome sequencing (WGS) amplicon-tiled sequencing approach on the Illumina platform, based on 17 wastewater samples. The current singleplex dPCR method shows 36% lower reagent costs and a 68% shorter turnaround time compared to WGS. Multiplexing can further reduce reagent costs by up to 84% and the start-to-finish time by 90%.

The dPCR genotyping assay requires two positive controls for SNP mutation and the wild-type allele (CTRL-100, CTRL-0). Additionally, one NTC is required to control the PCR reagent. In comparison, sequencing requires one positive and one negative control. This increases the number of samples for dPCR to 21 and sequencing to 20 samples.

Our calculations show that each dPCR run costs $375. Since we used singleplex assays in this study, each panel required four plates, increasing the total cost to $1500 and $88 per wastewater sample. In contrast, sequencing requires only one run but at a higher cost of $2345 per run, which is $138 per wastewater sample. Considering the current singleplex setup, the cost of reagents for the dPCR genotyping assay is 36% lower than the standard sequencing workflow. A significant advantage of the singleplex method is the ability to quickly switch out individual markers without the need to revalidate the entire multiplex assay, providing flexibility and reducing validation efforts. Additionally, with multiplexing, which is an available approach to save cost and time in PCR, the cost per run can be reduced by up to 84%, bringing it down to approximately $22 per sample, representing a significant cost advantage.

The dPCR workflow involves fewer steps compared to the multi-step sequencing workflow (which includes QC, library preparation, and cleaning). A dPCR run usually takes between 2-3 hours. The analysis requires thresholding and exploring the CSV file, which does not require extensive technical skills. The results provide the target variant quantity in the samples, which can be manually or automatically converted into copies in wastewater or prevalence in the wastewater. In our current setup, we ran four dPCR assays to generate data on four variants of interest, with a total sample-to-result time of about 12 hours. By multiplexing the dPCR assay, we believe the run time can be shortened to 4 hours. The singleplex method’s flexibility in switching out markers quickly without extensive revalidation is particularly beneficial for adapting to emerging variants and rapidly updating surveillance protocols.

On the other hand, sequencing runs require significant computing time to generate reads, and the output needs bioinformatic tools to convert it into actionable data on variant prevalence in wastewater, typically done through an automated pipeline. The sequencing method requires a higher degree of expertise to generate valid and reliable results compared to the dPCR method. The sample-to-result turnaround time for the sequencing method is 38 hours. Despite using singleplex assays, the dPCR method used in this study is 68% faster. We believe that using a multiplex assay can reduce the turnaround time by 90%, to less than a working day. This calculation does not consider limitations in sample shipment, logistics, and scheduling. In reality, a typical sequencing sample-to-result process might take weeks to be reported back to the customer. Consequently, the GISAID sequencing database is very useful for retrospective analysis, but for real-time monitoring, there is a delay of at least four weeks after sample collection. Therefore, wastewater genotyping is currently a practical solution to quantify lineages and identify new trends early.

### Lessons Learned and Opportunities for Improvement

We encountered numerous challenges during this project, resulting in valuable lessons for future deployment of dPCR based genotyping for wastewater:

1. Use Manufacturer-Specific Design Tools:

Utilize primer and probe design tools developed by the manufacturer of the dPCR instrument. In this project, ROSALIND used the Thermo Fisher qPCR primer and probe design tools.

2. Customized Assay Concentrations:

Work with an assay manufacturer that provides customized concentrations of primers and probes, allowing for optimization specific to wastewater samples.

3. Rapid Response to Emerging Variants:

New variants can arise quickly and are expected to appear in wastewater samples earlier than in clinical samples. Having verified assays ready for deployment as soon as possible is desirable, which might require designing, ordering, and testing assays weekly on a more frequent cadence.

4. Multiplex Assays:

Instead of singleplex assays, use multiplex assays to reduce the burden on the testing laboratories. Running four separate dPCR plates each week was necessary for validation but is likely to be viewed as onerous and costly in the long run. Based on our estimated costs for assays, controls, reagents, and plates, this amounts to approximately $88 per sample. Despite the cost, the faster turnaround time of dPCR (1 day vs. 1 week or longer for sequencing) can be beneficial.

5. QIAcuity Software Update Issues:

Instrument manufacturers regularly update the instrument software. We experienced an update to the QIAcuity dPCR instrument software, which slightly modified the data output format, causing issues during data upload to the dashboard. Implementing a QC check on data format can eliminate such interruptions.

6. Including SARS-CoV-2 Quantity in Panels:

Including SARS-CoV-2 levels in the panel can be a useful metric for monitoring community transmission. Understanding virus levels alongside variant prevalence provides a better picture of transmission dynamics. Although this study did not measure SARS-CoV-2 levels in wastewater, it is possible to do so by analyzing mutant and wild-type quantities in each assay. Further study is needed to assess the accuracy of SARS-CoV-2 levels based on these results.

## Conclusion

This study highlights the utility of wastewater-based genotyping as an effective way to monitor SARS-CoV-2 variants. By integrating genotyping and sequencing methods, we compared the prevalence of variants in wastewater samples with clinical and GISAID data sets, demonstrating a strong correlation and emphasizing the reliability of wastewater testing for community-level tracking of virus transmission.

The consistency in the rise and fall of variants across wastewater genotyping, clinical genotyping, and GISAID sequencing data strengthens the reliability of these trends and highlights the value of multiple surveillance methods. Although the accuracy of wastewater testing can be affected by sewershed-specific characteristics—hence, the results of virus levels in the wastewater must be interpreted with caution (52–54)—our one-year study shows that the prevalence of variants in the wastewater closely matches the variant prevalence in the clinical setting in the state of Georgia.

Understanding the dynamics of variant prevalence helps in predicting future trends and preparing appropriate public health responses. The data indicate that while some variants like BQ* and EG.1* may have short-lived impacts, others can rapidly dominate, necessitating swift action. The rapid emergence of the BA.2.86*/JN* variant and its quick dominance underscores the importance of agile monitoring systems and adaptive public health strategies to manage new variants effectively. The periodic spikes in the Unknown/Others category suggest ongoing viral evolution and the potential for new variants to emerge. Continuous genomic surveillance is essential for early detection and response to these changes.

The detection of the unknown marker and its subsequent identification as a new variant (BA.2.86*/JN*) also demonstrates the value of continuously updating and validating genotyping panels to accurately identify and track emerging variants. The sudden rise of unknown variants to 10% can indicate the introduction of a new variant. However, we found that the validation process can be challenging with limited access to the design of the assay and formulation. In this one-year study, one variant typically exceeds 50% prevalence each quarter, with a maximum of three variants comprising the profile. This approach could be improved if custom, multiplexed wastewater panels were developed and validated on a more regular basis.

The detailed analysis of multiple data sets from Georgia and Wisconsin revealed several key findings. First, the dominance and transition of SARS-CoV-2 variants, such as the shift from XBB to BA.2.86*/JN* in early 2024, were effectively captured in both wastewater and clinical samples. This underscores the capability of wastewater-based surveillance to provide early detection and extended monitoring of circulating variants, often preceding clinical detections by several weeks.

Comparisons between digital PCR and next-generation sequencing methods indicated that while NGS remains the gold standard for comprehensive variant analysis, dPCR is a cost-effective and faster alternative for routine surveillance, though the turnaround time is dependent on having functioning assays ready when new variants arise. Our cost analysis showed that dPCR genotyping is 36% cheaper in reagent costs and 68% faster in turnaround time compared to whole-genome sequencing (WGS). The potential for multiplexing in dPCR could further enhance its efficiency, reducing reagent costs by up to 84% and start-to-finish times by 90%.

The study also highlighted the challenges and lessons learned in development, validation, and implementation of a dPCR-based wastewater genotyping approach. The need for more consistent updating and validation of genotyping panels was evident, especially with the emergence of new variants like JN. The integration of SARS-CoV-2 quantification into genotyping panels can provide a more comprehensive understanding of variant transmission dynamics in the community.

Overall, our findings support the use of wastewater-based SARS-CoV-2 genotyping as a cost-effective and complementary approach to clinical sample genotyping and sequencing, offering a broader and more inclusive picture of variant prevalence and transmission. This approach is particularly valuable in times when clinical testing is limited, as observed during the study period.

## Data Availability

All data produced in the present study are available upon reasonable request to the authors

https://tracker.rosalind.bio/

## Acknowledgements

We gratefully acknowledge all data contributors, i.e., the Authors and their Originating laboratories responsible for obtaining the specimens, and their Submitting laboratories for generating the genetic sequence and metadata and sharing via the GISAID Initiative, on which this research is based.

This project has been funded in part by the NIH Rapid Acceleration of Diagnostics (RADxSM) initiative with federal funds from the National Institute of Biomedical Imaging and Bioengineering, National Institutes of Health. The current contract is funded from the Public Health and Social Services Emergency Fund through the Biomedical Advanced Research and Development Authority, HHS Office of the Assistant Secretary for Preparedness and Response, Department of Health and Human Services, under Contract No. 75N92021C00012.

## Notes

### Clinical Protocols

https://download-files.wixmp.com/ugd/f7710c_4dfc75069dca4c1598815705fed9059d.pdf?token=eyJhbGciOiJIUzI1NiIsInR5cCI6IkpXVCJ9.eyJpc3MiOiJ1cm46YXBwOmU2NjYzMGU3MTRmMDQ5MGFhZWExZjE0OWIzYjY5ZTMyIiwic3ViIjoidXJuOmFwcDplNjY2MzBlNzE0ZjA0OTBhYWVhMWYxNDliM2I2OWUzMiIsImF1ZCI6WyJ1cm46c2VydmljZTpmaWxlLmRvd25sb2FkIl0sImlhdCI6MTcyNDI1NTM4NCwiZXhwIjoxNzI0MjkxMzk0LCJqdGkiOiJmYjM5OWZlOC1jY2Q1LTQ3MGUtOGQyOS05YmQ2YjkxNzM3NDAiLCJvYmoiOltbeyJwYXRoIjoiL3VnZC9mNzcxMGNfNGRmYzc1MDY5ZGNhNGMxNTk4ODE1NzA1ZmVkOTA1OWQucGRmIn1dXSwiZGlzIjp7ImZpbGVuYW1lIjoiVmFyaWFudCBkZXRlY3Rpb24gU09QLSBSZXYuMS0xLnBkZiIsInR5cGUiOiJhdHRhY2htZW50In19.ykLKq53wW4JuvmK2DEOm14DR8XOrRKAwjOEqu1uMH9A

### Author Declarations

All clinical samples, retrospective and prospective, were collected predominantly in the states of Washington, Florida, and California. Aegis Sciences Corporation, Helix OpCo, and UW are Clinical Laboratory Improvement Amendments (CLIA) certified labs participating in the CDC National SARSCoV2 Strain Surveillance sequencing program to monitor variant distribution in the US. Ovation.io, Inc. is a human genomics company that operates the Ovation Research Network of CLIA certified labs that contributed variant data from 45 states, predominantly Utah and Michigan. The Pearl independent institutional review board gave ethical approval for the use of Aegis Sciences Corporation de-identified remnants of clinical testing. Western Institutional Review Board-Copernicus Group, the institutional review board of record for the Helix Respiratory Registry, gave ethical approval for the use of Helix OpCo de-identified remnants of clinical testing. Use of the UW de-identified excess clinical specimens was approved with a consent waiver by the UW IRB. Use of de-identified clinical remnant specimens was approved by individual labs, through their participation in the Ovation Research Network, and by the Ovation Ethics Committee.

### Summary of Updates

Updated IRB / oversight body statement. Corrected Saswata Sahoo's name in the author list.

